# Planning robust clinical trials for *Shigella* vaccines: A simulation-based evaluation of the impact of naturally-acquired immunity on vaccine performance

**DOI:** 10.64898/2025.12.11.25341848

**Authors:** Avnika B. Amin, Maria Garcia Quesada, Natalie E. Dean, Benjamin A. Lopman, James A. Platts-Mills, Elizabeth T. Rogawski McQuade

## Abstract

**Background:** Several *Shigella* vaccine candidates are in late stages of development, and the design of large Phase 3 trials in target populations is underway. Immunologic catch up by unvaccinated infants to vaccinated infants, which is determined by the trial site-specific force of infection, may modify the vaccine efficacy (VE) estimates observed in such trials. To set expectations and support optimal planning of future *Shigella* vaccine trials, we aimed to quantify the potential bias of VE estimates from the force of infection under different surveillance strategies and assumptions about *Shigella* immunity.

**Methods:** We simulated *Shigella* vaccine trials in low-, medium-, and high-burden settings under different assumptions about vaccine- and infection-conferred immunity. We evaluated six trial designs: active surveillance for *Shigella* infection or symptom-based reporting paired with single-outcome, stratified recurrent outcome, or crude recurrent-outcome Cox proportional hazards models. In each scenario, we evaluated bias, 95% confidence interval coverage, and the false negative rate.

**Findings:** Only the trial design using active surveillance for infection and single outcome models was consistently unbiased across all examined scenarios. The other five designs underestimated VE by up to an absolute 20%, with a single outcome model paired with symptom-based reporting performing least poorly. Bias generally was worse at higher-burden sites. For realistically sized trials, the 95% CI coverage and false negative rate were also worse at higher-burden sites.

**Interpretation:** The observed VE in *Shigella* vaccine trials is likely to underestimate true VE. Careful trial design is needed to minimize this underestimation.

## Introduction

*Shigella* is the leading bacterial cause of diarrheal disease globally.(1,2) Children aged <5 years experience the most *Shigella*-related morbidity and mortality,(3) and children in low-resource settings, particularly in south Asia and sub-Saharan Africa, are most affected.(1,4) Beyond the immediate morbidity and mortality from disease, *Shigella* can also have longer-term effects like linear growth faltering,(5,6) which has been associated with increased susceptibility to infection, delays in development, and worsening of undernutition.(7,8) While *Shigella* is treatable with antibiotics, diagnostic testing is largely unavailable in low-resource settings,(9) and antimicrobial resistance is increasing.(10) Effective *Shigella* vaccines are poised to prevent poor outcomes from shigellosis and potentially reduce further antibiotic resistance. Several *Shigella* vaccine candidates are currently being evaluated, with Phase 3 studies in target populations in the planning phase.(9,11)

Robust planning of Phase 3 *Shigella* trials requires both accurate expectations for vaccine efficacy (VE) and an understanding of how immunity mechanisms affect appropriate trial design and analysis. Lessons from rotavirus vaccines provide a cautionary tale. Rotavirus vaccines have lower VE in high-burden compared to low-burden settings.(12,13) Explanations like nutritional deficiencies, co-administered oral poliovirus vaccine, and passively-acquired antibody interference were initially proposed, but interventions to address these factors only modestly improved rotavirus vaccine immunogenicity.(14) At least some of the discrepancy in VE between high- and low-resource settings may be due to how quickly unvaccinated infants accrue infection-acquired immunity.(15) The faster they immunologically catch up to vaccinated infants due to higher force of infection in high-burden settings, the more that VE estimated in a trial underestimates VE in a fully susceptible population.(16) This phenomenon will also likely affect the evaluation of *Shigella* vaccines and pose a key challenge for planning clinical trials and interpreting trial results.

To support the optimal evaluation of *Shigella* vaccines, we conducted a simulation study to quantify the potential impact of accrued infection-acquired immunity on VE estimates from a hypothetical phase III efficacy trial. We evaluated the impact of varying the force of infection, the strength of infection-acquired immunity compared to vaccine-conferred immunity, post-vaccination immunity gains, and vaccine effects on infection risk. We then evaluated whether different trial designs and analytical strategies may improve the ability of the trial to estimate a VE that approximates the targeted VE in a fully susceptible population. These findings can inform trial site selection, priority gaps in our understanding of immunity, and protocols for data collection and analysis.

## Methods

### Simulation structure

We simulated a Phase 3 vaccine trial where healthy, immune-naïve infants were enrolled and 1:1 randomized to two doses of either a placebo or a hypothetical *Shigella* vaccine. Doses were administered at 6.5 and 9.5 months of age to be compatible with the EPI schedule. After a two-week immune ramp-up period, follow-up to ascertain primary outcomes began at 10 months of age. To characterize the broader relationships between the parameters varied in each simulation scenario while minimizing variability due to small sample size, we first simulated an unrealistically large trial cohort. We assumed 5,000 infants were enrolled in each trial arm (i.e., more than double the size of several rotavirus vaccine trials in LMICs)(17,18) and that all of them remained infection-naïve between enrollment and the start of follow-up. We simulated recurrent *Shigella* infections between 10 and 36 months of age, when follow-up ended, and assumed no loss to follow-up. The recurrent infection process is conceptually represented and described in Supplemental Figure 1.

To simulate recurrent *Shigella* infections, we modelled (Supplemental Equation) (i) the hazard of infection over time, (ii) the conditional probability of progressing to diarrhea given infection, and (iii) the conditional probability of experiencing severe diarrhea given symptoms. We model each component as a function of age group, vaccination status, and infection-acquired immunity. *Shigella* natural histories that were identified in the MAL-ED study, a longitudinal birth cohort study of the first two years of life, informed age trends in hazards and severity probabilities for the 10-12, 13-15, 16-18, 19-21, and 22-24 month age groups.(19) Based on studies reporting declines in *Shigella* incidence and severity after the second year of life,(20) we assumed that hazards and severity probabilities peaked for the 22-24 month age groups and subsequently declined for the 25-30 and 31-36 month age groups. Modifications by vaccination status and infection-acquired immunity depended on the specific simulation scenario, described below.

### Simulation scenarios

In line with the target product characteristics,(20) the vaccine was assumed to have 40% efficacy against Shigella diarrhea and 60% efficacy against severe Shigella diarrhea. For simplicity, we assumed that efficacy did not wane during follow-up, there was no loss to follow-up, and the population was homogeneous with respect to covariates.

We examined 24 different scenarios (summarized in Supplemental Figure 2) where four factors were varied: 1) the vaccine effect on infection hazard, 2) the strength of infection-acquired immunity, 3) hybrid immunity (i.e., whether natural infection provided additional protection beyond vaccination), and 4) the force of infection (i.e., hazard of infection among susceptible individuals in the population). In all scenarios, unvaccinated infants could accrue immunity until they reached the maximum possible immunity for a vaccinated infant. VE against infection (factor 1) was either 0% (i.e., no effect on the infection hazard) or 20%. The degree of protection after an infection, or infection-acquired immunity (factor 2) was equivalent whether the infection was asymptomatic or symptomatic and was either comparable or inferior to vaccination. If inferior, infection reduced the probability of subsequent diarrhea by 10% and the conditional probability of a severe outcome by 20%. For hybrid immunity (factor 3), vaccinated infants either did not acquire additional immunity from natural infections or acquired additional immunity from, at most, one infection.

We evaluated three forces of infection (factor 4) to reflect *Shigella* burdens at different trial sites in low-resource settings. One represented a relatively low-burden site (based on incidence in the MAL-ED Brazil site), another represented a relatively high-burden site (based on incidence in the MAL-ED Bangladesh site),(19) and the third represented a relatively medium-burden site based on the midpoint between the low- and high-burden sites. Infection incidence rates, as well as progressions to expected diarrhea and severe diarrhea incidence rates, are in Supplemental Table 1.

In a sensitivity analysis, we evaluated how VE estimates might be affected by the underlying true VE by varying true VE against any *Shigella* diarrhea in the scenario corresponding to no vaccine-conferred protection against infection, comparable infection- and vaccine-conferred immunity, hybrid immunity does not occur, and a medium-burden site.

### Trial structure for analysis

We simulated 500 *Shigella* vaccine trials for each scenario. Before data analysis, we selected data from the simulations to be consistent with two types of data collection protocols: active infection surveillance and passive symptom-based reporting. For the (unrealistic) trial setting of active infection surveillance, we assumed all diarrhea episodes and subclinical infections would be identified in the trial, which allows for subclinical infections that elicit immunity to be incorporated into analysis. In the symptom-based reporting setting, we assumed all diarrhea episodes would be identified in the trial, but not subclinical infections. This is more consistent with common and feasible approaches to outcome ascertainment that are based on participants presenting to care when ill or by continuously monitoring participants for symptoms. In this setting, subclinical infections are not observed and cannot be considered in analysis.

### Data analysis

All analyses used a Cox proportional hazard model with time since 10 days after second dose as the underlying time scale and trial arm (placebo or vaccine) as the only covariate. We estimated VE against both any and severe *Shigella* diarrhea. VE was calculated as (1-hazard ratio) * 100% over the full follow-up period (10 days after second dose to 36 months) and stratified by the first (10-22 months) and second years (23-34 months) of follow-up. 95% confidence intervals (CIs) for the VE estimates were calculated similarly with the CI bounds for the hazard ratio.

We evaluated three methods to estimate VE: a single outcome model, a stratified recurrent outcome model, and a crude recurrent outcome model. The single outcome model censored individuals after the first observed *Shigella* episode, which was either *Shigella* infection (regardless of symptoms) when active infection surveillance was used or *Shigella* diarrhea when symptom-based reporting was used. The recurrent outcome models did not censor individuals and included multiple endpoints for individuals if they occurred. The stratified model allowed the baseline hazard to differ for individuals with different observed *Shigella* episode histories. The crude model assumed a constant baseline hazard unaffected by event history. For recurrent outcome models, robust variances were used to account for correlated outcomes from the same individuals and the first two days after a *Shigella* episode were excluded from analysis time to reflect that a new episode could not be defined until after at least two diarrhea-free days.

We evaluated a total of six approaches based on the structure for data collection and analytic approach: active and passive surveillance paired with all three models (summarized in Supplemental Table 2).

To compare VE results, we examined bias and mean squared error (MSE). Bias was calculated as the average VE estimate for a set of simulations minus the true VE in a fully susceptible population, while MSE was calculated as the average of the square of the estimate minus the true VE parameter.(21) We evaluated the percent of 95% CIs that contained the true VE parameter (coverage) and the percent that incorrectly indicated a non-significant effect (false negative rate).

To evaluate results that might be observed in realistically sized *Shigella* vaccine trials, we determined the minimum size *n* required in each trial arm for 80% power to detect 60% VE against severe *Shigella* diarrhea. For each simulation run, we used data from the last 200 infant IDs in each trial arm to represent pilot data that informed the minimum *n*. We calculated *n* with the powerSurvEpi R package based on the two statistical models (recurrent and single outcome models). We then restricted data to the first *n* infant IDs in each trial arm and repeated the six previously-described analytic approaches.

Finally, to understand how bias would change with different true VEs in a fully susceptible population, we varied VE against any *Shigella* diarrhea between 10% and 90%. We set VE against severe *Shigella* diarrhea to the same value as VE against any *Shigella* diarrhea to avoid a scenario where vaccination would make severe *Shigella* diarrhea more likely, given symptomatic *Shigella* infection. For simplicity, we only evaluated one scenario: one where vaccination did not affect the infection hazard, infection-acquired immunity was comparable to vaccination, hybrid immunity does not occur, and the trial was conducted at a medium-burden site. All simulations and analyses were conducted in R, with code available on GitHub.

Given the number of scenarios and performance metrics examined, we parse out key findings that are consistent when only one aspect of a scenario is changed. We focus the results on VE against severe *Shigella* diarrhea, as that is the expected primary endpoint for candidate *Shigella* vaccine trials. The full results for VE estimates against any and severe *Shigella* diarrhea across all simulation scenarios and analytic approaches are presented in Supplemental Tables 3-26.

## Results

### Comparison of approaches in large trials

When trials were highly powered (i.e., 5,000 infants in each trial arm) and true VE against severe *Shigella* diarrhea in a fully susceptible population was 60%, only the trial design using active surveillance and a single outcome model yielded unbiased results (Figure 1, Supplemental Table 3). Under this trial design, the mean estimate of VE against severe *Shigella* diarrhea was 59.5% from trials conducted at a medium-burden site when infection- and vaccine-conferred immunity were comparable, vaccination did not reduce the infection hazard, and hybrid immunity does not occur (Supplemental Table 3). However, mean VE estimates ranged from 47.3% to 51.7% (bias of an absolute -8.3% to -12.7%) when using the other five trial designs in the same scenario (Supplemental Tables 4-8).

**Figure 1.**
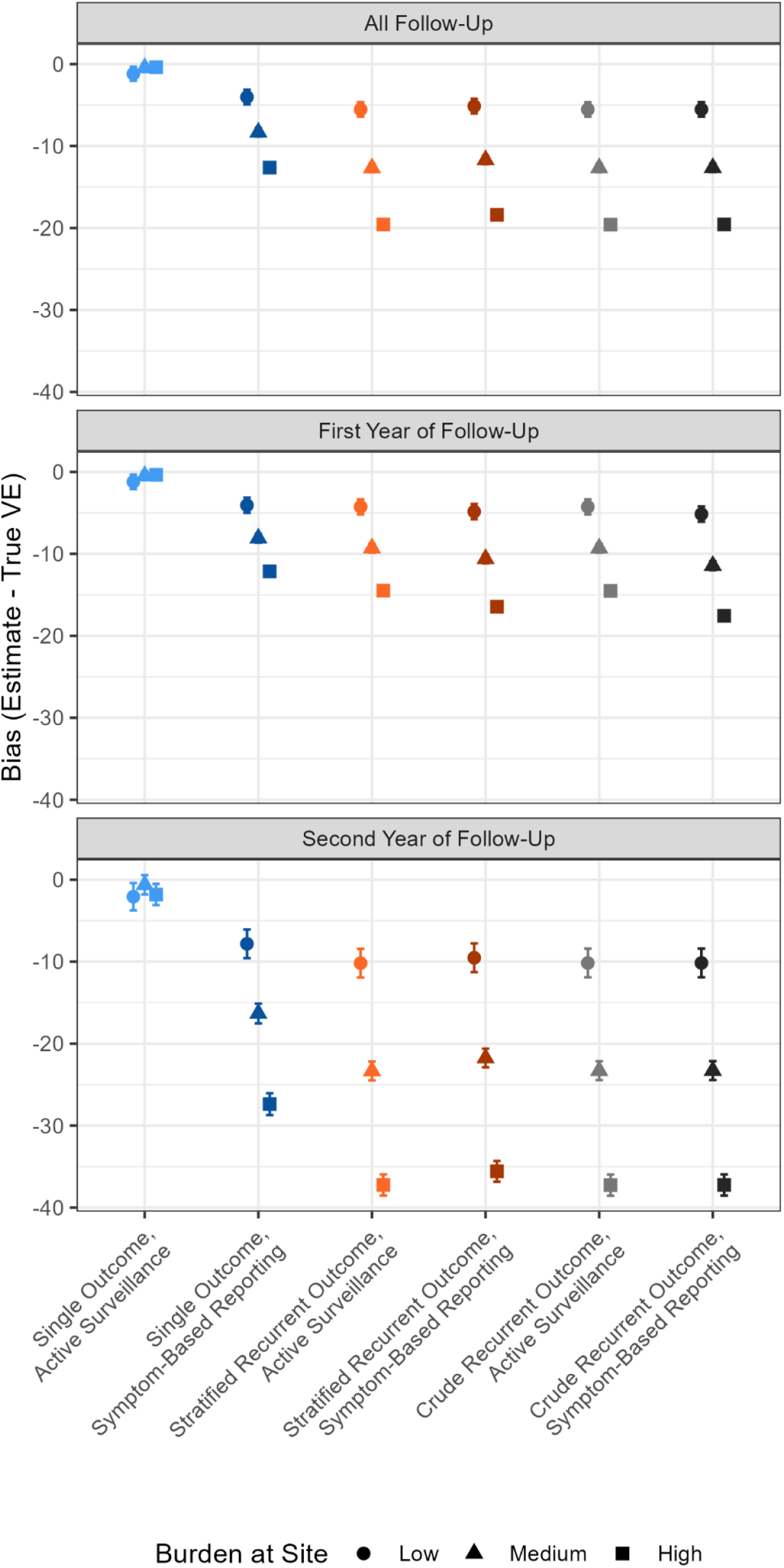
Bias of the estimated vaccine efficacy (VE) against severe, *Shigella*-attributable disease and 95% confidence intervals for well-powered trials with 5,000 infants in each trial arm when different approaches to data collection and analysis are used. A: VE is estimated for the full follow-up period (10-36 months); B: VE is estimated for the first year of follow-up (10-22 months); C: VE is estimated for the second year of follow-up (23-34 months). True VE in a fully susceptible population against severe *Shigella* diarrhea is 60%. These results derive from scenarios when infection- and vaccine-conferred immunity are comparable, vaccination does not reduce the infection hazard, and no hybrid immunity is possible.

For most trial designs (excluding single outcome active surveillance), the underestimation of VE in a fully susceptible population worsened as the force of infection increased (Figure 1, Supplemental Tables 4-8). At low-burden sites, bias did not substantially differ between the biased analysis approaches, with mean estimates of VE against severe *Shigella* diarrhea ranging from 54.5% to 56% (approximately -5% bias; Supplemental Tables 4-8). However, as the force of infection increased, results from symptom-based reporting paired with a single outcome model were less biased than approaches using recurrent outcome models. Mean VE estimates from symptom-based reporting with a single outcome model were 51.7% and 47.4% at medium- and high-burden sites, respectively (corresponding to bias of -8.3% and - 12.6%; Supplemental Table 4). In contrast, bias from the recurrent outcome model approaches neared 20% in the high-burden site.

The magnitudes of bias for the VE estimates from the first year of follow-up were similar to those from the full follow-up time (Figure 1, Supplemental Tables 3 and 9). However, bias was substantially larger when estimating VE during the second year of follow-up, nearing -40% at the high-burden site. As expected for highly powered trials, VE results were very precise, resulting in low coverage of 95% CIs and rare false negatives (Supplemental Tables 3-8).

### Comparison of approaches in realistic trial sizes

When more realistic sample sizes were used for trials (Supplemental Tables 9-14), bias was comparable to that seen with well-powered trials for each approach. 95% CI coverage was >90% at low-burden sites regardless of the trial design approach used. However, the 95% CI coverage and the false negative rate worsened as the force of infection increased for all approaches except when active surveillance was paired with a single outcome model (Figure 2, Supplemental Tables 9-14). A single outcome model with symptom-based reporting was the next best option, as it had a lower false negative rate and better 95% CI coverage than designs using recurrent outcome models. For example, at a high-burden site, pairing a single outcome model with symptom-based reporting meant that 82.2% of 95% CIs contained the true VE and a false conclusion of no vaccine effect was drawn in 42.2% of simulation runs (Figure 2, Supplemental Table 10).

**Figure 2.**
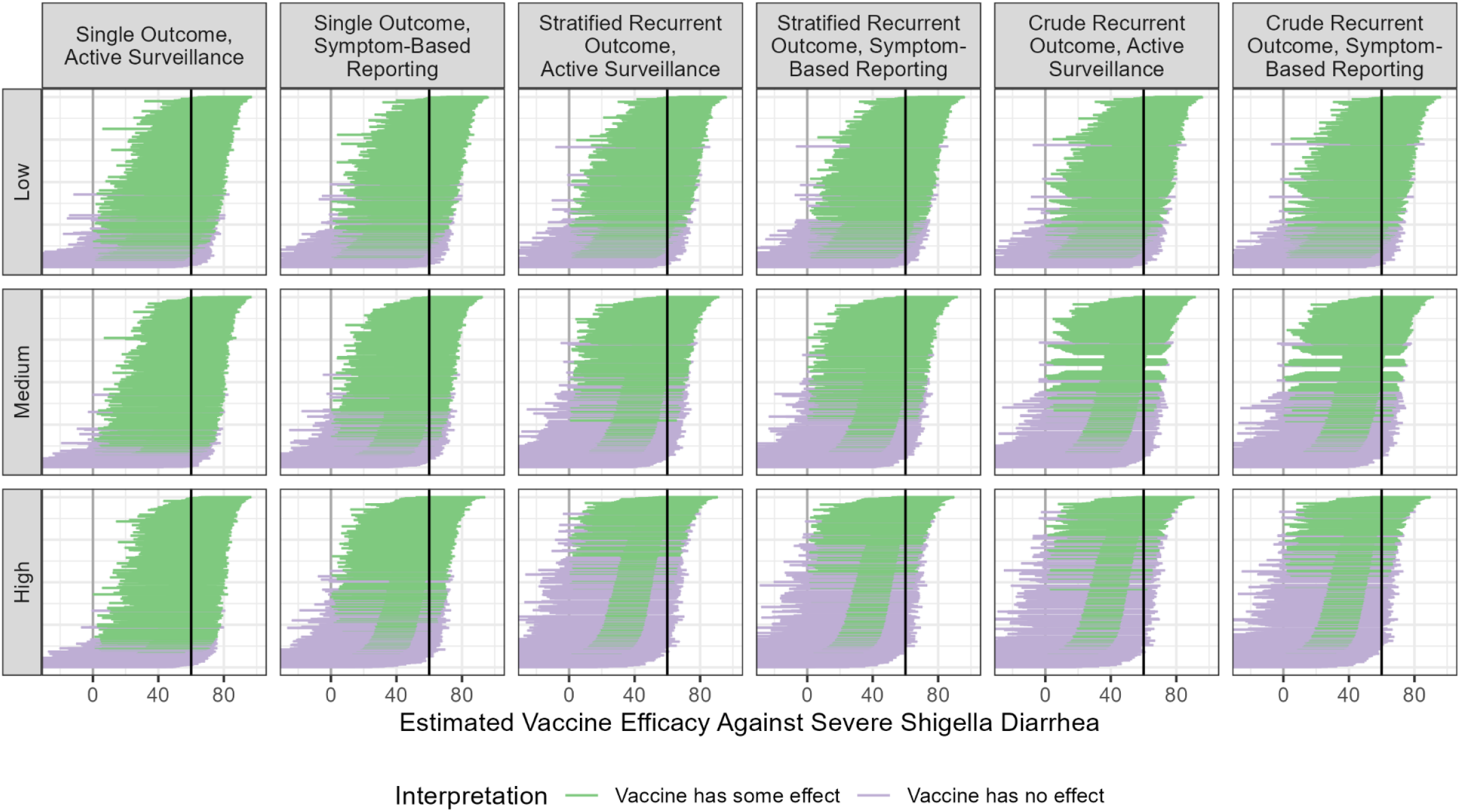
Estimated vaccine efficacy against severe *Shigella* diarrhea and 95% confidence intervals for trials with realistic sample sizes under different *Shigella* forces of infection and trial designs. The thick grey vertical rule indicates the null (0% efficacy), while the thick black vertical rule indicates the truth (60% efficacy). Purple indicates false negatives and green indicates correct conclusions about the vaccine effect. These results reflect scenarios when infection- and vaccine-conferred immunity are comparable, vaccination does not reduce the infection hazard, and hybrid immunity does not occur.

### Alternative assumptions about immunity

The bias of VE estimates differed depending on the assumptions made about infection- and vaccine-conferred immunity. Pairing active surveillance with a single outcome model continued to yield unbiased results regardless of the immunity scenario (Supplemental Tables 3 and 9). For the remainder of this section, we will focus on describing results for the five designs with the most bias (symptom-based reporting with a single outcome model and active surveillance and symptom-based reporting paired with stratified and crude recurrent outcome models) in the large trial (5,000 infants per arm).

Assumptions around hybrid immunity appeared most influential. Estimates from scenarios where hybrid immunity was possible were only slightly biased regardless of the other scenario conditions or trial design used (Supplemental Tables 4-8). Scenarios where hybrid immunity was not possible resulted in more extreme underestimation of VE in a fully susceptible population. For example, at a medium-burden site, mean bias ranged from -4.1% to -12.9% when hybrid immunity was not possible but only from -0.3% to -3.4% when hybrid immunity was possible, regardless of the assumptions around infection-versus vaccine-conferred immunity or the effect of the vaccine on infection hazard (Figure 3, Supplemental Tables 4-8).

**Figure 3.**
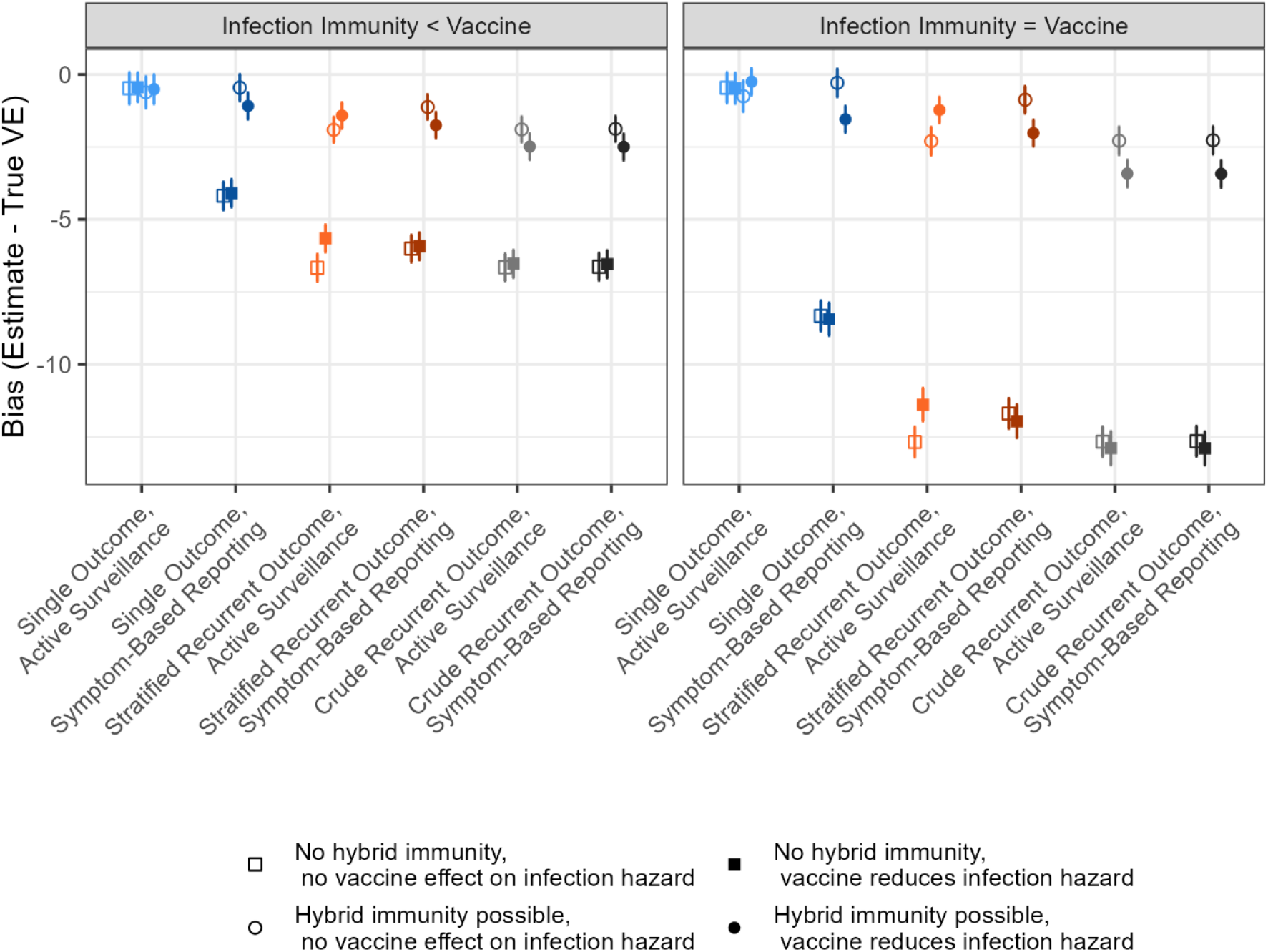
Bias of the estimated vaccine efficacy (VE) against severe, *Shigella*-attributable disease and 95% confidence intervals for well-powered trials under different scenarios of vaccine- and infection-conferred immunity. True VE in a fully susceptible population is 60%. These results reflect trials conducted at a medium-burden site.

The similarity between infection- and vaccine-conferred immunity mattered more when hybrid immunity was not possible, with worse bias when infection- and vaccine-conferred immunity were comparable. When hybrid immunity was not possible and a medium-burden site was used, mean bias ranged from -4.1% to -6.7% if infection-conferred immunity was less protective than vaccination (Figure 3, Supplemental Tables 4-8). However, when infection- and vaccine-conferred immunity were similar, mean bias ranged from -9.3% to -12.9% (Figure 3, Supplemental Tables 4-8). No clear trends emerged when comparing scenarios where vaccination did and did not affect hazard of infection.

### Sensitivity to true VE in a fully susceptible population

The true VE in a fully susceptible population also affected the magnitude of bias (Figure 4). Pairing active surveillance with a single outcome model resulted in no bias for any VE against any *Shigella* diarrhea. Bias for the other designs showed a U-shaped relationship, with small bias for small or large true VE, and larger bias as VE approached 50%. Consistent with prior results, designs that used recurrent outcome models were more biased than the design pairing symptom-based reporting with a single outcome model.

**Figure 4.**
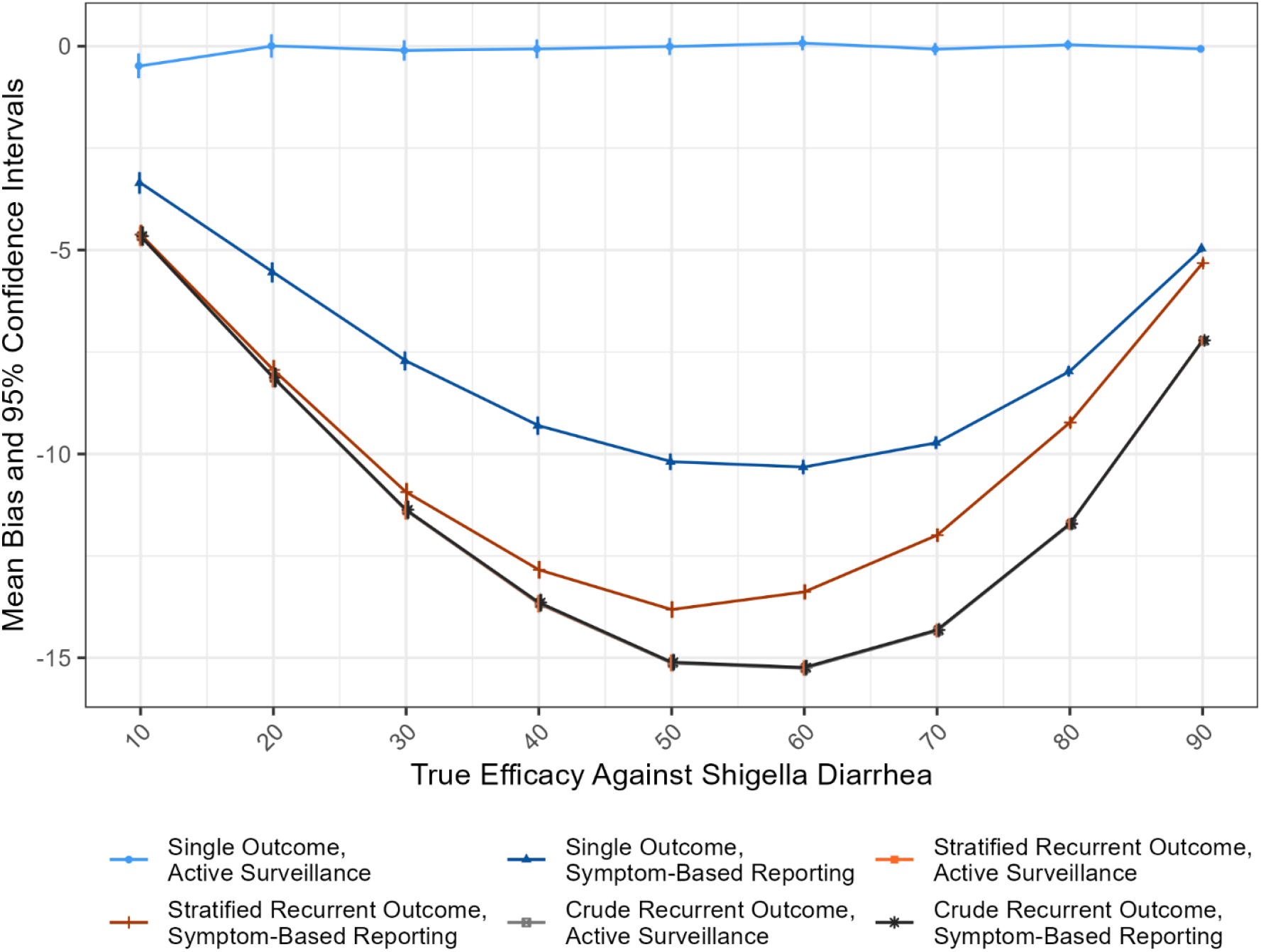
Mean bias of the estimated vaccine efficacy against any *Shigella* diarrhea with varying true efficacy in a fully susceptible population and 95% confidence intervals for well-powered trials conducted at a medium-burden site. These results reflect scenarios when infection- and vaccine-conferred immunity are comparable, vaccination does not reduce the infection hazard, and no hybrid immunity is possible. Note: estimates from the stratified recurrent outcome, active surveillance and the two crude recurrent outcome models are nearly overlapping.

## Discussion

We demonstrate that the VE observed in future *Shigella* vaccine trials is likely to underestimate true VE in a fully susceptible population because of the accrual of natural immunity. While the observed VE will faithfully capture VE specific to the setting of the trial, in which natural immunity will be a relevant factor, we argue that the target VE of interest in Phase III trials is VE in a fully susceptible population, which is likely to be more generalizable across settings and better captures the “biological” vaccine effect. The impact of natural immunity on VE estimates is most extreme when this true VE is close to 50%.

The magnitude of underestimation of the target VE depends on several factors, with the force of infection being the most influential. We demonstrate that VE underestimation worsens as the force of infection increases, with the worst bias – VE estimates an absolute 20% too low – at high-burden sites.

This is especially important as high-burden sites are often favored for clinical trials, since the larger number of expected events means that fewer infants need to be enrolled to obtain the desired statistical power. Our results suggest that trials at high-burden sites may lead to the incorrect conclusion about a null *Shigella* VE as often as 50-60% of the time, and the lower sample size required due to high incidence will be offset by needing a larger sample size to detect a smaller VE. Importantly, because the impact of natural immunity increases with longer duration of follow-up, and is most striking for estimates specific to later periods of follow-up, studies in high-burden settings may incorrectly conclude that vaccine-induced immunity wanes rapidly, when the truth is rather that natural immunity was accruing over follow-up.

To some extent, decisions around a trial’s data collection protocol and analysis can mitigate the magnitude of underestimation at high-burden sites. Active surveillance for *Shigella* infection paired with a single outcome regression model, where individuals are censored at their first infection, was consistently unbiased no matter the site burden or other assumptions around immunity. However, active infection surveillance is unlikely to be realistic as it would require regular testing of asymptomatic individuals. Even symptom-based reporting (with either active or passive surveillance) is unlikely to detect all symptomatic cases, especially when detection of cases at presentation to care is the norm. Our results indicate that bias is most mitigated when a single outcome model is used, although VE will be underestimated by an average of an absolute 10% in most scenarios.

Other aspects of infection- and vaccine-conferred immunity also affect expected bias, with the general principle being that bias is worse when unvaccinated infants can immunologically “catch-up” to vaccinated infants. For example, when hybrid immunity does not occur, vaccinated infants cannot gain additional immunity, which allows unvaccinated infants to more easily catch up to them immunologically. A similar principle applies to our findings around the similarity between infection- and vaccine-conferred immunity. When the two are equal, unvaccinated infants only need one infection to catch up to a vaccinated infant; however, when infection-conferred immunity is inferior, unvaccinated infants must experience multiple infections to catch up to the vaccinated infants. In the unexamined scenario in which infection-conferred immunity is superior to vaccine-conferred immunity, bias would be expected to be even larger.

This study is subject to a few limitations. The primary limitation is that our results, particularly the magnitude of bias, will be sensitive to our input parameters. Very little is known about *Shigella* natural histories beyond data from the few longitudinal birth cohort studies,(2,22) one of which informed our study. We assumed that vaccine immunity did not wane during the follow-up period, but waning is certainly possible. Depending on how quickly *Shigella* vaccine immunity wanes, bias may worsen since waning would allow unvaccinated infants to catch up to vaccinated infants more quickly. We also assumed that all infants were homogeneous with respect to susceptibility and pre-existing immunity. Realistically, it may be difficult to ascertain an infant’s infection history, and trials may have to contend with unobserved differences in baseline infection-conferred immunity.

Overall, we demonstrate how observed VE from clinical trials will underestimate VE in a fully susceptible population depending on the force of infection and mechanisms of immunity. These results help to set expectations for future *Shigella* vaccine trials and inform the selection of trial sites. Furthermore, our findings shed light both on key considerations for trial and analysis design and on gaps in knowledge that may affect *Shigella* vaccine trial results.

## Data Availability

All data produced are available online at GitHub.

https://github.com/AAmin23/paper-replications/tree/56675ffab2e836e604e38dbe7b7c1e532f2dc2bf/shig-trial-sims

## Notes

### Competing Interest Statement

The authors have declared no competing interest.

