## Supplemental Materials for "Planning robust clinical trials for *Shigella* vaccines: A simulation-based evaluation of the impact of naturally-acquired immunity on vaccine performance"

Supplemental Information

Supplemental Figure 1. Simulation structure and process for generating recurrent infections.

Supplemental Equation. Multiplier approach to generate an individual's infection hazard and disease progression probabilities based on their age, vaccination status, and infection history.

Supplemental Figure 2. Full set of simulation scenarios with different assumptions around vaccine effects, infection-acquired immunity, hybrid immunity, and the force of infection.

Supplemental Table 1. *Shigella* natural history parameters and infection incidence rates at low-, medium-, and high-burden sites.

Supplemental Table 2. Summary of analytic approaches to evaluating ideal and realistic data generated from simulated clinical *Shigella* vaccine trials.

Supplemental Table 3. Mean estimate, bias, mean squared error (MSE), coverage, and false negative rate for vaccine efficacy (VE) estimates against severe *Shigella* diarrhea from highly powered trials for all 24 simulation scenarios when using active surveillance for infection and a single outcome regression model.

Supplemental Table 4. Mean estimate, bias, mean squared error (MSE), coverage, and false negative rate for vaccine efficacy (VE) estimates against severe *Shigella* diarrhea from highly powered trials for all 24 simulation scenarios when using symptom-based reporting and a single outcome regression model.

Supplemental Table 5. Mean estimate, bias, mean squared error (MSE), coverage, and false negative rate for vaccine efficacy (VE) estimates against severe *Shigella* diarrhea from highly powered trials for all 24 simulation scenarios when using active surveillance for infection and a stratified recurrent outcome regression model.

Supplemental Table 6. Mean estimate, bias, mean squared error (MSE), coverage, and false negative rate for vaccine efficacy (VE) estimates against severe *Shigella* diarrhea from highly powered trials for all 24 simulation scenarios when using symptom-based reporting and a stratified recurrent outcome regression model.

Supplemental Table 7. Mean estimate, bias, mean squared error (MSE), coverage, and false negative rate for vaccine efficacy (VE) estimates against severe *Shigella* diarrhea from highly powered trials for all 24 simulation scenarios when using active surveillance for infection and a crude recurrent outcome regression model.

Supplemental Table 8. Mean estimate, bias, mean squared error (MSE), coverage, and false negative rate for vaccine efficacy (VE) estimates against severe *Shigella* diarrhea from highly powered trials for all 24 simulation scenarios when using symptom-based reporting and a crude recurrent outcome regression model.

Supplemental Table 9. Mean estimate, bias, mean squared error (MSE), coverage, and false negative rate for vaccine efficacy (VE) estimates against severe *Shigella* diarrhea from highly powered trials for all 6 design approaches applied to stratified datasets for the first and second year of follow-up.

Supplemental Table 10. Mean estimate, bias, mean squared error (MSE), coverage, and false negative rate for vaccine efficacy (VE) estimates against severe *Shigella* diarrhea from realistically sized trials for all 24 simulation scenarios when using active surveillance for infection and a single outcome regression model.

Supplemental Table 11. Mean estimate, bias, mean squared error (MSE), coverage, and false negative rate for vaccine efficacy (VE) estimates against severe *Shigella* diarrhea from realistically sized trials for all 24 simulation scenarios when using symptom-based reporting and a single outcome regression model.

Supplemental Table 12. Mean estimate, bias, mean squared error (MSE), coverage, and false negative rate for vaccine efficacy (VE) estimates against severe *Shigella* diarrhea from realistically sized trials for all 24 simulation scenarios when using active surveillance for infection and a stratified recurrent outcome regression model.

Supplemental Table 13. Mean estimate, bias, mean squared error (MSE), coverage, and false negative rate for vaccine efficacy (VE) estimates against severe *Shigella* diarrhea from realistically sized trials for all 24 simulation scenarios when using symptom-based reporting and a stratified recurrent outcome regression model.

Supplemental Table 14. Mean estimate, bias, mean squared error (MSE), coverage, and false negative rate for vaccine efficacy (VE) estimates against severe *Shigella* diarrhea from realistically sized trials for all 24 simulation scenarios when using active surveillance for infection and a crude recurrent outcome regression model.

Supplemental Table 15. Mean estimate, bias, mean squared error (MSE), coverage, and false negative rate for vaccine efficacy (VE) estimates against severe *Shigella* diarrhea from realistically sized trials for all 24 simulation scenarios when using symptom-based reporting and a crude recurrent outcome regression model.

Supplemental Table 16. Mean estimate, bias, mean squared error (MSE), coverage, and false negative rate for vaccine efficacy (VE) estimates against any *Shigella* diarrhea from highly powered trials for all 24 simulation scenarios when using active surveillance for infection and a single outcome regression model.

Supplemental Table 17. Mean estimate, bias, mean squared error (MSE), coverage, and false negative rate for vaccine efficacy (VE) estimates against any *Shigella* diarrhea from highly powered trials for all 24 simulation scenarios when using symptom-based reporting and a single outcome regression model.

Supplemental Table 18. Mean estimate, bias, mean squared error (MSE), coverage, and false negative rate for vaccine efficacy (VE) estimates against any *Shigella* diarrhea from highly powered trials for all 24 simulation scenarios when using active surveillance for infection and a stratified recurrent outcome regression model.

Supplemental Table 19. Mean estimate, bias, mean squared error (MSE), coverage, and false negative rate for vaccine efficacy (VE) estimates against any *Shigella* diarrhea from highly powered trials for all 24 simulation scenarios when using symptom-based reporting and a stratified recurrent outcome regression model.

Supplemental Table 20. Mean estimate, bias, mean squared error (MSE), coverage, and false negative rate for vaccine efficacy (VE) estimates against any *Shigella* diarrhea from highly powered trials for all 24 simulation scenarios when using active surveillance for infection and a crude recurrent outcome regression model.

Supplemental Table 21. Mean estimate, bias, mean squared error (MSE), coverage, and false negative rate for vaccine efficacy (VE) estimates against any *Shigella* diarrhea from highly powered trials for all 24 simulation scenarios when using symptom-based reporting and a crude recurrent outcome regression model.

Supplemental Table 22. Mean estimate, bias, mean squared error (MSE), coverage, and false negative rate for vaccine efficacy (VE) estimates against any *Shigella* diarrhea from realistically sized trials for all 24 simulation scenarios when using active surveillance for infection and a single outcome regression model.

Supplemental Table 23. Mean estimate, bias, mean squared error (MSE), coverage, and false negative rate for vaccine efficacy (VE) estimates against any *Shigella* diarrhea from realistically sized trials for all 24 simulation scenarios when using symptom-based reporting and a single outcome regression model.

Supplemental Table 24. Mean estimate, bias, mean squared error (MSE), coverage, and false negative rate for vaccine efficacy (VE) estimates against any *Shigella* diarrhea from realistically sized trials for all 24 simulation scenarios when using active surveillance for infection and a stratified recurrent outcome regression model.

Supplemental Table 25. Mean estimate, bias, mean squared error (MSE), coverage, and false negative rate for vaccine efficacy (VE) estimates against any *Shigella* diarrhea from realistically sized trials for all 24 simulation scenarios when using symptom-based reporting and a stratified recurrent outcome regression model.

Supplemental Table 26. Mean estimate, bias, mean squared error (MSE), coverage, and false negative rate for vaccine efficacy (VE) estimates against any *Shigella* diarrhea from realistically sized trials for all 24 simulation scenarios when using active surveillance for infection and a crude recurrent outcome regression model.

Supplemental Table 27. Mean estimate, bias, mean squared error (MSE), coverage, and false negative rate for vaccine efficacy (VE) estimates against any *Shigella* diarrhea from realistically sized trials for all 24 simulation scenarios when using symptom-based reporting and a crude recurrent outcome regression model.

**Supplemental Figure 1. Simulation structure and process for generating recurrent infections.** Infections are simulated for one trial enrollee at a time. An enrollee's baseline hazard changes as they move through different age groups during the course of the trial, though this baseline hazard is constant for the entire time spent in a given age group. Baseline hazard is further modified by vaccine-conferred immunity (when applicable) and immunity accrued from infections that occur during follow-up.

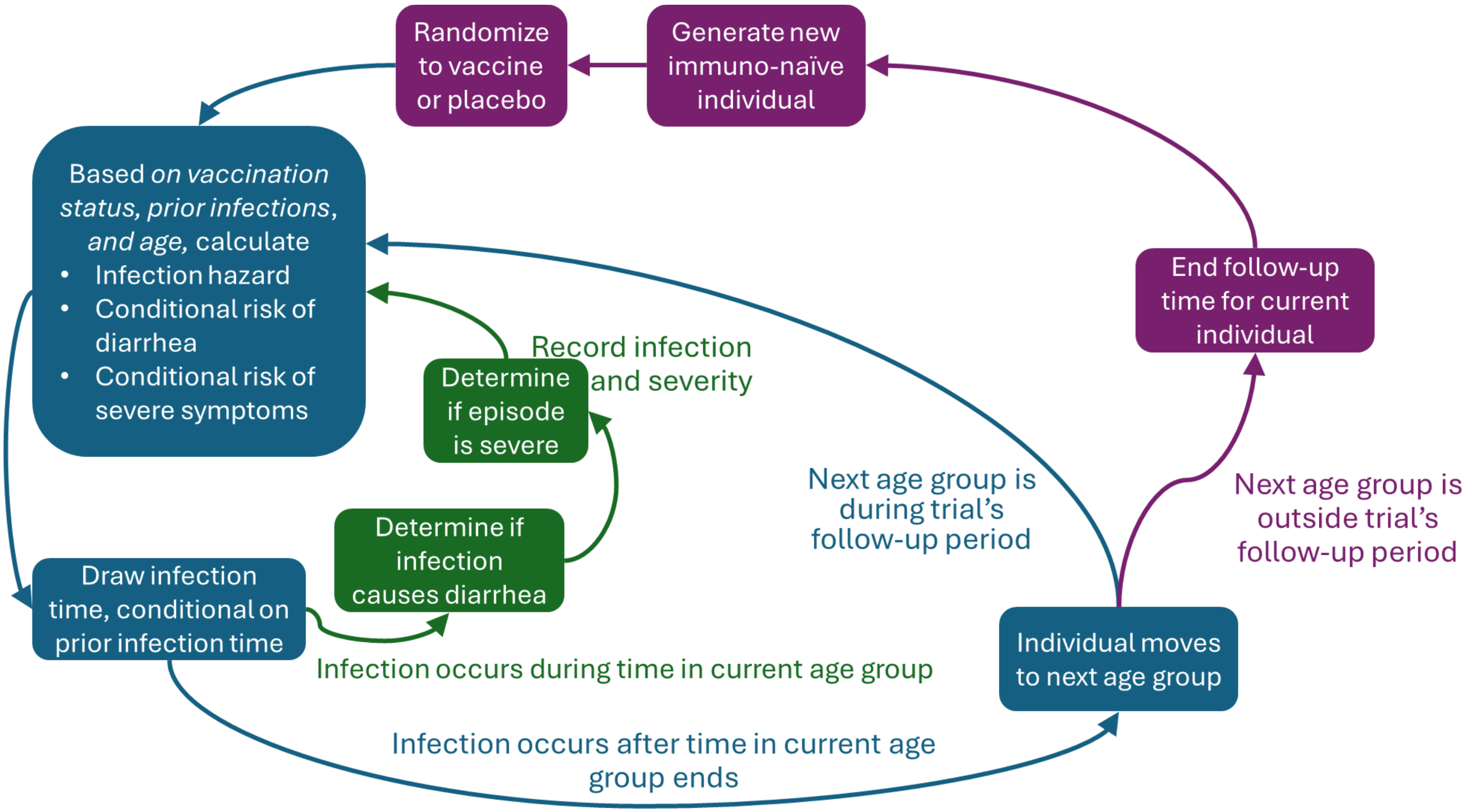

Each infant began the recurrent infection process at 10 months of age with no infection-acquired immunity. An infection hazard reflecting their vaccination status and age was calculated. This hazard was used to generate an infection day $T_i$ from an exponential distribution conditioned on the starting age (in days) for the 10-12 month age group. If $T_i$ occurred while the infant was still in the 10-12 month age group (i.e., through 365 days of age), they were infected. If not infected between 10 and 12 months of age, the hazard was updated to reflect the next age group and the infection generation process repeated.

Upon infection, the severity experienced was then determined with binomial draws using probabilities again calculated based on vaccination status and age. The first determined if they developed diarrhea; if so, a second draw determined if they had a severe outcome. The infection, age at infection (in days), and severity were recorded. For second and later infections, infection hazard and disease probabilities were modified to reflect infection-acquired immunity, the exponential distribution now conditioned on the age of the last infection, and another $T_i$ generated. Once the generated $T_i$ exceeded the end of the age group, the individual then entered the next age group with infection hazard and severity probabilities updated to reflect age and all relevant sources of immunity. This recurrent process continued through the last age group.

**Supplemental Equation. Multiplier approach to generate an individual's infection hazard and disease progression probabilities based on their age, vaccination status, and infection history.**

Among the larger population that the trial is being conducted in, let

$h$ be the *Shigella* infection rate
$d$ be the probability of developing symptoms, given a *Shigella* infection
$s$ be the probability of experiencing severe disease, given a symptomatic *Shigella* infection

To allow for age-based trends in infection hazard, symptom development, and severe disease, let

$a = 1, 2, \ldots, k$ be all the age groups that occur between the start and end ages for trial follow-up
$rh_a$ be the hazard of *Shigella* infection for the $k^{th}$ age group within the larger population relative to the hazard in the referent age group
$rd_a$ be the probability of developing symptoms, given a *Shigella* infection, for the $k^{th}$ age group within the larger population relative to the probability in the referent age group
$rs_a$ be the probability of experiencing severe disease, given a symptomatic *Shigella* infection, for the $k^{th}$ age group within the larger population relative to the probability in the referent age group

Other factors that impact infection hazard and symptom and severity probabilities include

$i = 0, 1, 2, \ldots$, the number of prior infections an individual has had that have conferred immunity
$rh_i$, the hazard of another *Shigella* infection after $i$ previous infections relative to $i - 1$ previous infections
$rd_i$, the probability of developing symptoms, given a *Shigella* infection, after $i$ previous infections relative to $i - 1$ previous infections
$rs_i$, the relative probability of experiencing severe disease, given a symptomatic *Shigella* infection, after $i$ previous infections
$v = 0,1$, where 1 indicates the individual received the experimental *Shigella* vaccine and 0 indicates they did not
$1 - VE_h$, the relative hazard of another *Shigella* infection after vaccination compared to not being vaccinated
$1 - VE_d$, the relative probability of developing symptoms, given a *Shigella* infection, after vaccination compared to not being vaccinated
$1 - VE_s$, the relative probability of experiencing severe disease, given a symptomatic *Shigella* infection, after vaccination compared to not being vaccinated

An individual's infection hazard at a given time during trial follow-up is calculated as

$$h \times [(1 - VE_h)^v \times rh_i \times rh_a]$$

Probability of developing symptoms given infection and probability of experiencing severe disease, given symptomatic infection, are analogously calculated using the corresponding $d$- and $s$-based notation, respectively.

**Supplemental Figure 2. Full set of simulation scenarios with different assumptions around vaccine effects, infection-acquired immunity, hybrid immunity, and the force of infection.** When considering the maximum possible reductions to infection hazard and disease progression probabilities, unvaccinated infants can reach, but not exceed, the maximum possible immunity that vaccinated infants may accrue.

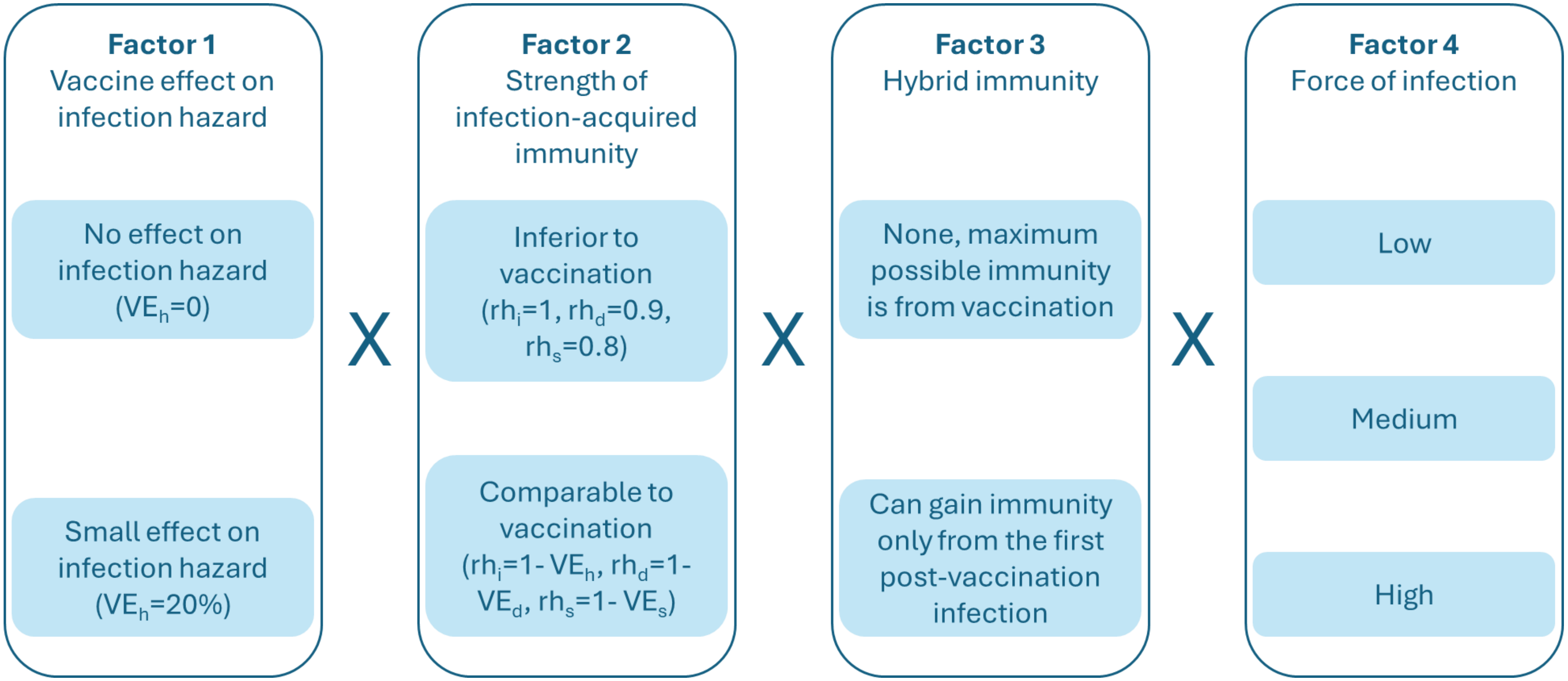

**Supplemental Table 1. *Shigella* natural history parameters and infection incidence rates at low-, medium-, and high-burden sites.** $rh_a$ is the *Shigella* infection hazard for a given age group relative to the hazard in the referent age group. $rd_a$ is the probability of developing symptoms, given a *Shigella* infection, for a given age group relative to the probability in the referent age group. $rs_a$ is the probability of experiencing severe disease, given a symptomatic *Shigella* infection, for a given age group relative to the probability in the referent age group. Absolute hazards were 0.0007147541 infections/day for the low-burden site, 0.002254098 infections/day for the medium-burden site, and 0.003793443 infections/day for the high-burden site. Disease progression probabilities did not vary by site, with the probability of symptoms given infection equal to 0.35 and the probability of severe disease given symptoms equal to 0.084.

| Age (months) | $rh_a$ | $rd_a$ | $rs_a$ |
|---|---|---|---|
| 10-12 | 0.539 | 1.029 | 1.488 |
| 13-15 | 0.669 | 1.114 | 1.845 |
| 16-18 | 0.860 | 1.114 | 1.845 |
| 19-21 | 1.000 | 1.000 | 1.000 |
| 22-24 | 0.920 | 1.000 | 1.000 |
| 25-30* | 0.900 | 0.900 | 0.900 |
| 31-36* | 0.800 | 0.800 | 0.800 |
| *These ages were not part of the original study used to inform parameters. Based on the available evidence suggesting *Shigella* burden peaks in the 2nd year of life, we specified these parameters to represent decreases in hazard and disease progression probabilities. | | | |

**Supplemental Table 2. Summary of analytic approaches to evaluating ideal and realistic data generated from simulated clinical *Shigella* vaccine trials.** Cox proportion hazards models were used for all regression models, with age as the underlying time scale and trial arm (placebo or vaccine) as a model covariate.

| Analytic approach | Regression model type | Data collection approach |
|---|---|---|
| 1 | Single outcome (censored after first infection) | Active surveillance for infection (all infections ascertained) |
| 2 | Single outcome (censored after first diarrheal episode) | Symptom-based reporting (only diarrhea episodes ascertained) |
| 3 | Recurrent outcome (strata based on prior infections) | Active surveillance for infection (all infections ascertained) |
| 4 | Recurrent outcome (strata based on prior diarrheal episodes) | Symptom-based reporting (only diarrhea episodes ascertained) |
| 5 | Crude (event history not considered) | Active surveillance for infection (all infections ascertained) |
| 6 | Crude (event history not considered) | Symptom-based reporting (only diarrhea episodes ascertained) |

**Supplemental Table 3. Mean estimate, bias, mean squared error (MSE), coverage, and false negative rate for vaccine efficacy (VE) estimates against severe *Shigella* diarrhea from highly powered trials for all 24 simulation scenarios when using active surveillance for infection and a single outcome regression model.** Infection-acquired immunity was either inferior or similar to vaccine-acquired immunity. Hybrid immunity was either not possible (no) or possible such that the first post-vaccination infection conferred additional immunity (yes). Bias was calculated as the average VE estimate for a set of simulations minus the true VE (60%). MSE was calculated as the average of the square of the estimate minus the true parameter. Coverage represents the percent of 95% confidence intervals (CIs) that contained the true VE (60%), and the false negative rate indicates the percent of 95% CIs that incorrectly indicated a non-significant effect.

| VE against infection | Strength of infection-acquired immunity | Hybrid immunity | Force of infection | Mean VE estimate (range) | Bias (95% CI) | MSE (95% CI) | Coverage (95% CI) | False negatives (95% CI) |
|---|---|---|---|---|---|---|---|---|
| 0% | Similar | No | Low | 58.8% (15%, 79.2%) | -1.2 (-2.1, -0.3) | 98.8 (83.9, 113.7) | 94.2% (92.2%, 96.2%) | 1.4% (0.4%, 2.4%) |
| | | | Medium | 59.5% (43.7%, 74.9%) | -0.5 (-1, 0.1) | 35.9 (31.7, 40.2) | 95.4% (93.6%, 97.2%) | 0% (0%, 0%) |
| | | | High | 59.6% (43.3%, 73.2%) | -0.4 (-0.8, 0.1) | 27.6 (24.2, 31) | 95.4% (93.6%, 97.2%) | 0% (0%, 0%) |
| | | Yes | Low | 59.2% (27.3%, 79.8%) | -0.8 (-1.6, 0) | 89.8 (77.5, 102.1) | 95.6% (93.8%, 97.4%) | 1.6% (0.5%, 2.7%) |
| | | | Medium | 59.2% (37.2%, 73.4%) | -0.8 (-1.3, -0.2) | 36.9 (32, 41.9) | 95.8% (94%, 97.6%) | 0% (0%, 0%) |
| | | | High | 59.6% (38.7%, 73.7%) | -0.4 (-0.8, 0.1) | 26.5 (22.9, 30.2) | 95.8% (94%, 97.6%) | 0% (0%, 0%) |
| | Inferior | No | Low | 59.2% (27%, 81.2%) | -0.8 (-1.7, 0.1) | 100.1 (85.9, 114.4) | 92.4% (90.1%, 94.7%) | 2.6% (1.2%, 4%) |
| | | | Medium | 59.5% (38.1%, 75.2%) | -0.5 (-1, 0.1) | 38.7 (33.5, 43.9) | 95.6% (93.8%, 97.4%) | 0% (0%, 0%) |
| | | | High | 59.9% (40%, 74.1%) | -0.1 (-0.6, 0.4) | 29.7 (25.8, 33.6) | 95.8% (94%, 97.6%) | 0% (0%, 0%) |
| | | Yes | Low | 59.4% (25.2%, 82.4%) | -0.6 (-1.5, 0.2) | 94.3 (80.5, 108.1) | 93.4% (91.2%, 95.6%) | 1.6% (0.5%, 2.7%) |
| | | | Medium | 59.4% (38.1%, 73.5%) | -0.6 (-1.2, -0.1) | 38.5 (33.5, 43.5) | 95.4% (93.6%, 97.2%) | 0% (0%, 0%) |
| | | | High | 59.8% (38.3%, 74.1%) | -0.2 (-0.6, 0.3) | 28.8 (24.9, 32.7) | 96% (94.3%, 97.7%) | 0% (0%, 0%) |
| 20% | Similar | No | Low | 59.2% (9.5%, 82.3%) | -0.8 (-1.6, 0) | 86 (71.7, 100.4) | 94.6% (92.6%, 96.6%) | 0.6% (-0.1%, 1.3%) |
| | | | Medium | 59.5% (41.7%, 75.9%) | -0.5 (-1, 0) | 35.8 (31.5, 40.1) | 95.2% (93.3%, 97.1%) | 0% (0%, 0%) |
| | | | High | 60.1% (37.1%, 73.4%) | 0.1 (-0.4, 0.6) | 30.7 (26.4, 35) | 92.4% (90.1%, 94.7%) | 0% (0%, 0%) |

| | | | | | | | | |
|---|---|---|---|---|---|---|---|---|
| | | Yes | Low | 59.1% (23.9%, 85.3%) | -0.9 (-1.7, -0.1) | 88.6 (75.6, 101.5) | 95.2% (93.3%, 97.1%) | 1.6% (0.5%, 2.7%) |
| | | | Medium | 59.8% (43.4%, 74.9%) | -0.2 (-0.7, 0.2) | 27.6 (24.1, 31.1) | 97.4% (96%, 98.8%) | 0% (0%, 0%) |
| | | | High | 59.7% (38.2%, 74.7%) | -0.3 (-0.8, 0.1) | 26.2 (22.3, 30) | 96.4% (94.8%, 98%) | 0% (0%, 0%) |
| | Inferior | No | Low | 58.9% (21.6%, 82.3%) | -1.1 (-2, -0.3) | 91.2 (78.1, 104.3) | 94% (91.9%, 96.1%) | 1.2% (0.2%, 2.2%) |
| | | | Medium | 59.6% (40.8%, 73.5%) | -0.4 (-0.9, 0.1) | 32.9 (28.5, 37.3) | 95.2% (93.3%, 97.1%) | 0% (0%, 0%) |
| | | | High | 60.1% (45%, 75.7%) | 0.1 (-0.3, 0.6) | 23.7 (20.7, 26.8) | 96.2% (94.5%, 97.9%) | 0% (0%, 0%) |
| | | Yes | Low | 58.9% (25.7%, 86.3%) | -1.1 (-1.9, -0.2) | 92.2 (79.1, 105.3) | 94.2% (92.2%, 96.2%) | 1.2% (0.2%, 2.2%) |
| | | | Medium | 59.5% (39.1%, 73.3%) | -0.5 (-1, 0) | 33.2 (28.7, 37.8) | 96.2% (94.5%, 97.9%) | 0% (0%, 0%) |
| | | | High | 60.3% (42.8%, 74%) | 0.3 (-0.1, 0.8) | 25.4 (22.3, 28.5) | 94.8% (92.9%, 96.7%) | 0% (0%, 0%) |

**Supplemental Table 4. Mean estimate, bias, mean squared error (MSE), coverage, and false negative rate for vaccine efficacy (VE) estimates against severe *Shigella* diarrhea from highly powered trials for all 24 simulation scenarios when using symptom-based reporting and a single outcome regression model. Infection-acquired immunity was either inferior or similar to vaccine-acquired immunity.** Hybrid immunity was either not possible (no) or possible such that the first post-vaccination infection conferred additional immunity (yes). Bias was calculated as the average VE estimate for a set of simulations minus the true VE (60%). MSE was calculated as the average of the square of the estimate minus the true parameter. Coverage represents the percent of 95% confidence intervals (CIs) that contained the true VE (60%), and the false negative rate indicates the percent of 95% CIs that incorrectly indicated a non-significant effect.

| VE against infection | Strength of infection-acquired immunity | Hybrid immunity | Force of infection | Mean VE estimate (range) | Bias (95% CI) | MSE (95% CI) | Coverage (95% CI) | False negatives (95% CI) |
|---|---|---|---|---|---|---|---|---|
| 0% | Similar | No | Low | 56% (21.4%, 76.4%) | -4 (-4.9, -3.1) | 119.5 (102.2, 136.9) | 92.6% (90.3%, 94.9%) | 2.6% (1.2%, 4%) |
| | | | Medium | 51.7% (32.8%, 68.2%) | -8.3 (-8.8, -7.8) | 104.4 (94.3, 114.6) | 70.2% (66.2%, 74.2%) | 0% (0%, 0%) |
| | | | High | 47.4% (29%, 63.2%) | -12.6 (-13.1, -12.1) | 190.2 (177.2, 203.2) | 28.6% (24.6%, 32.6%) | 0% (0%, 0%) |
| | | Yes | Low | 59.6% (23.3%, 80.8%) | -0.4 (-1.2, 0.4) | 84.8 (73.2, 96.4) | 94.8% (92.9%, 96.7%) | 0.8% (0%, 1.6%) |
| | | | Medium | 59.7% (41.4%, 72.5%) | -0.3 (-0.8, 0.2) | 30.3 (26.5, 34) | 95.8% (94%, 97.6%) | 0% (0%, 0%) |
| | | | High | 58.9% (44.4%, 71.7%) | -1.1 (-1.5, -0.7) | 21.8 (18.8, 24.8) | 94.4% (92.4%, 96.4%) | 0% (0%, 0%) |
| | Inferior | No | Low | 58% (22.8%, 80.2%) | -2 (-2.8, -1.1) | 100.5 (85.5, 115.4) | 91.6% (89.2%, 94%) | 2% (0.8%, 3.2%) |
| | | | Medium | 55.8% (37.9%, 71.9%) | -4.2 (-4.7, -3.7) | 47.8 (42.1, 53.5) | 89% (86.3%, 91.7%) | 0% (0%, 0%) |
| | | | High | 53.5% (37.3%, 65.9%) | -6.5 (-6.9, -6) | 64.5 (57.7, 71.2) | 73% (69.1%, 76.9%) | 0% (0%, 0%) |
| | | Yes | Low | 59.6% (23%, 80.9%) | -0.4 (-1.2, 0.4) | 87.1 (74.6, 99.5) | 92.6% (90.3%, 94.9%) | 1% (0.1%, 1.9%) |
| | | | Medium | 59.5% (43.3%, 73.3%) | -0.5 (-0.9, 0) | 27.2 (23.9, 30.5) | 96.4% (94.8%, 98%) | 0% (0%, 0%) |
| | | | High | 58.9% (40.4%, 70.2%) | -1.1 (-1.5, -0.7) | 20.2 (17.4, 23.1) | 96.2% (94.5%, 97.9%) | 0% (0%, 0%) |
| 20% | Similar | No | Low | 56.6% (16.2%, 80.2%) | -3.4 (-4.2, -2.6) | 98.7 (84, 113.3) | 93.6% (91.5%, 95.7%) | 1.4% (0.4%, 2.4%) |
| | | | Medium | 51.6% (27.1%, 67.6%) | -8.4 (-9, -7.9) | 112.5 (100.5, 124.5) | 69.6% (65.6%, 73.6%) | 0% (0%, 0%) |
| | | | High | 47.8% (30.9%, 62.1%) | -12.2 (-12.7, -11.7) | 184.4 (170.6, 198.2) | 31.2% (27.1%, 35.3%) | 0% (0%, 0%) |

| | | | | | | | | |
|---|---|---|---|---|---|---|---|---|
| | | Yes | Low | 58.8% (23%, 84.2%) | -1.2 (-2, -0.4) | 85.2 (73, 97.5) | 95.2% (93.3%, 97.1%) | 1.2% (0.2%, 2.2%) |
| | | | Medium | 58.5% (41.6%, 73.7%) | -1.5 (-2, -1.1) | 29.3 (25.4, 33.1) | 95.8% (94%, 97.6%) | 0% (0%, 0%) |
| | | | High | 57% (38.2%, 70.3%) | -3 (-3.5, -2.6) | 34.8 (29.9, 39.7) | 90% (87.4%, 92.6%) | 0% (0%, 0%) |
| | Inferior | No | Low | 57.6% (23.5%, 80.1%) | -2.4 (-3.2, -1.5) | 92.2 (79.2, 105.3) | 94.4% (92.4%, 96.4%) | 1% (0.1%, 1.9%) |
| | | | Medium | 55.9% (37%, 71.4%) | -4.1 (-4.6, -3.6) | 46.4 (40.4, 52.3) | 89% (86.3%, 91.7%) | 0% (0%, 0%) |
| | | | High | 54% (39.5%, 69.5%) | -6 (-6.4, -5.6) | 56.8 (51.2, 62.5) | 75.4% (71.6%, 79.2%) | 0% (0%, 0%) |
| | | Yes | Low | 58.9% (22.7%, 81.9%) | -1.1 (-1.9, -0.3) | 85.8 (73.6, 97.9) | 94.2% (92.2%, 96.2%) | 0.8% (0%, 1.6%) |
| | | | Medium | 58.9% (42%, 73.1%) | -1.1 (-1.5, -0.6) | 28.3 (24.6, 32) | 95.4% (93.6%, 97.2%) | 0% (0%, 0%) |
| | | | High | 58.2% (40.4%, 70.1%) | -1.8 (-2.2, -1.4) | 23.5 (20.2, 26.8) | 93.4% (91.2%, 95.6%) | 0% (0%, 0%) |

**Supplemental Table 5. Mean estimate, bias, mean squared error (MSE), coverage, and false negative rate for vaccine efficacy (VE) estimates against severe *Shigella* diarrhea from highly powered trials for all 24 simulation scenarios when using active surveillance for infection and a stratified recurrent outcome regression model.** Hybrid immunity was either not possible (no) or possible such that the first post-vaccination infection conferred additional immunity (yes). Bias was calculated as the average VE estimate for a set of simulations minus the true VE (60%). MSE was calculated as the average of the square of the estimate minus the true parameter. Coverage represents the percent of 95% confidence intervals (CIs) that contained the true VE (60%), and the false negative rate indicates the percent of 95% CIs that incorrectly indicated a non-significant effect.

| VE against infection | Strength of infection-acquired immunity | Hybrid immunity | Force of infection | Mean VE estimate (range) | Bias (95% CI) | MSE (95% CI) | Coverage (95% CI) | False negatives (95% CI) |
|---|---|---|---|---|---|---|---|---|
| 0% | Similar | No | Low | 54.5% (18.9%, 75.9%) | -5.5 (-6.4, -4.6) | 134.5 (115.2, 153.8) | 90.4% (87.8%, 93%) | 2.8% (1.4%, 4.2%) |
| | | | Medium | 47.3% (30.1%, 63.3%) | -12.7 (-13.2, -12.2) | 196.1 (182, 210.2) | 38.6% (34.3%, 42.9%) | 0% (0%, 0%) |
| | | | High | 40.4% (18.2%, 57.2%) | -19.6 (-20.1, -19.1) | 416.4 (395.7, 437.2) | 2.4% (1.1%, 3.7%) | 0% (0%, 0%) |
| | | Yes | Low | 59% (25.6%, 79.7%) | -1 (-1.8, -0.2) | 83.3 (72.2, 94.5) | 96% (94.3%, 97.7%) | 0.8% (0%, 1.6%) |
| | | | Medium | 57.7% (39.7%, 70.8%) | -2.3 (-2.8, -1.8) | 35.2 (30.7, 39.7) | 94.2% (92.2%, 96.2%) | 0% (0%, 0%) |
| | | | High | 55.4% (38.4%, 66.7%) | -4.6 (-5, -4.2) | 44.6 (39, 50.3) | 84.4% (81.2%, 87.6%) | 0% (0%, 0%) |
| | Inferior | No | Low | 57.2% (25.3%, 80.5%) | -2.8 (-3.6, -1.9) | 106 (90.2, 121.8) | 91.4% (88.9%, 93.9%) | 2.2% (0.9%, 3.5%) |
| | | | Medium | 53.3% (36%, 66.8%) | -6.7 (-7.1, -6.2) | 73.2 (65.7, 80.8) | 77.4% (73.7%, 81.1%) | 0% (0%, 0%) |
| | | | High | 49.2% (32.8%, 60.7%) | -10.8 (-11.2, -10.4) | 140.2 (130, 150.4) | 28.8% (24.8%, 32.8%) | 0% (0%, 0%) |
| | | Yes | Low | 59.2% (23.9%, 80.8%) | -0.8 (-1.7, 0) | 88.1 (75.6, 100.6) | 93.2% (91%, 95.4%) | 0.8% (0%, 1.6%) |
| | | | Medium | 58.1% (41.9%, 72.3%) | -1.9 (-2.3, -1.5) | 28.5 (24.9, 32.2) | 94.8% (92.9%, 96.7%) | 0% (0%, 0%) |
| | | | High | 56.2% (40.7%, 66.3%) | -3.8 (-4.2, -3.5) | 33.2 (28.9, 37.4) | 85.8% (82.7%, 88.9%) | 0% (0%, 0%) |
| 20% | Similar | No | Low | 55.7% (18.4%, 79.8%) | -4.3 (-5.1, -3.5) | 104 (89.3, 118.8) | 92.6% (90.3%, 94.9%) | 1.4% (0.4%, 2.4%) |
| | | | Medium | 48.6% (26.4%, 66.8%) | -11.4 (-12, -10.8) | 171.9 (156.7, 187.1) | 49.6% (45.2%, 54%) | 0% (0%, 0%) |
| | | | High | 42.8% (25%, 60.9%) | -17.2 (-17.7, -16.6) | 329.7 (310.8, 348.7) | 8.4% (6%, 10.8%) | 0% (0%, 0%) |

| | | | | | | | | |
|---|---|---|---|---|---|---|---|---|
| | | Yes | Low | 59% (26.4%, 84.6%) | -1 (-1.8, -0.2) | 81.6 (70.1, 93.2) | 95.8% (94%, 97.6%) | 0.8% (0%, 1.6%) |
| | | | Medium | 58.8% (41.4%, 73.2%) | -1.2 (-1.7, -0.8) | 27.1 (23.4, 30.9) | 96.4% (94.8%, 98%) | 0% (0%, 0%) |
| | | | High | 57% (41%, 70.9%) | -3 (-3.5, -2.6) | 33.4 (28.8, 37.9) | 89.8% (87.1%, 92.5%) | 0% (0%, 0%) |
| | Inferior | No | Low | 57.1% (25.2%, 80.8%) | -2.9 (-3.7, -2.1) | 89.7 (77.2, 102.2) | 94% (91.9%, 96.1%) | 1% (0.1%, 1.9%) |
| | | | Medium | 54.3% (35.1%, 69.3%) | -5.7 (-6.1, -5.2) | 60.4 (53.4, 67.4) | 82.2% (78.8%, 85.6%) | 0% (0%, 0%) |
| | | | High | 51% (36.1%, 65.9%) | -9 (-9.4, -8.6) | 100.8 (92.8, 108.8) | 46.2% (41.8%, 50.6%) | 0% (0%, 0%) |
| | | Yes | Low | 58.9% (28.8%, 81.9%) | -1.1 (-1.9, -0.3) | 79 (68.8, 89.3) | 95.6% (93.8%, 97.4%) | 0.8% (0%, 1.6%) |
| | | | Medium | 58.6% (39.1%, 73.3%) | -1.4 (-1.9, -1) | 27.5 (23.8, 31.3) | 94.6% (92.6%, 96.6%) | 0% (0%, 0%) |
| | | | High | 57.2% (40.4%, 70.2%) | -2.8 (-3.2, -2.5) | 27 (23.3, 30.7) | 90% (87.4%, 92.6%) | 0% (0%, 0%) |

**Supplemental Table 6. Mean estimate, bias, mean squared error (MSE), coverage, and false negative rate for vaccine efficacy (VE) estimates against severe *Shigella* diarrhea from highly powered trials for all 24 simulation scenarios when using symptom-based reporting and a stratified recurrent outcome regression model.** Hybrid immunity was either not possible (no) or possible such that the first post-vaccination infection conferred additional immunity (yes). Bias was calculated as the average VE estimate for a set of simulations minus the true VE (60%). MSE was calculated as the average of the square of the estimate minus the true parameter. Coverage represents the percent of 95% confidence intervals (CIs) that contained the true VE (60%), and the false negative rate indicates the percent of 95% CIs that incorrectly indicated a non-significant effect.

| VE against infection | Strength of infection-acquired immunity | Hybrid immunity | Force of infection | Mean VE estimate (range) | Bias (95% CI) | MSE (95% CI) | Coverage (95% CI) | False negatives (95% CI) |
|---|---|---|---|---|---|---|---|---|
| 0% | Similar | No | Low | 54.9% (19.8%, 76%) | -5.1 (-6, -4.2) | 129.7 (110.8, 148.5) | 91.4% (88.9%, 93.9%) | 2.6% (1.2%, 4%) |
| | | | Medium | 48.3% (31.1%, 63.8%) | -11.7 (-12.2, -11.2) | 171.2 (158.2, 184.2) | 47% (42.6%, 51.4%) | 0% (0%, 0%) |
| | | | High | 41.6% (20.5%, 58.4%) | -18.4 (-18.9, -17.9) | 371.1 (351.7, 390.5) | 4.4% (2.6%, 6.2%) | 0% (0%, 0%) |
| | | Yes | Low | 59.6% (26.3%, 79.9%) | -0.4 (-1.2, 0.4) | 80.6 (70, 91.1) | 95.8% (94%, 97.6%) | 0.8% (0%, 1.6%) |
| | | | Medium | 59.1% (40.7%, 71.9%) | -0.9 (-1.3, -0.4) | 29.1 (25.4, 32.8) | 96.4% (94.8%, 98%) | 0% (0%, 0%) |
| | | | High | 57.4% (41.5%, 68.7%) | -2.6 (-3, -2.2) | 28.9 (24.9, 33) | 91.2% (88.7%, 93.7%) | 0% (0%, 0%) |
| | Inferior | No | Low | 57.4% (25.4%, 80.8%) | -2.6 (-3.4, -1.7) | 105.2 (89.5, 120.9) | 91% (88.5%, 93.5%) | 2.2% (0.9%, 3.5%) |
| | | | Medium | 54% (37.1%, 67.4%) | -6 (-6.5, -5.5) | 64.3 (57.4, 71.1) | 81.2% (77.8%, 84.6%) | 0% (0%, 0%) |
| | | | High | 50.1% (34.7%, 61.6%) | -9.9 (-10.3, -9.4) | 119.7 (110.4, 129) | 37.8% (33.5%, 42.1%) | 0% (0%, 0%) |
| | | Yes | Low | 59.4% (25.1%, 80.9%) | -0.6 (-1.4, 0.2) | 87.3 (75, 99.5) | 93% (90.8%, 95.2%) | 0.8% (0%, 1.6%) |
| | | | Medium | 58.9% (43.3%, 72.4%) | -1.1 (-1.5, -0.7) | 25.1 (21.9, 28.2) | 96.6% (95%, 98.2%) | 0% (0%, 0%) |
| | | | High | 57.3% (42%, 67.4%) | -2.7 (-3.1, -2.3) | 24.9 (21.5, 28.3) | 90.2% (87.6%, 92.8%) | 0% (0%, 0%) |
| 20% | Similar | No | Low | 55.5% (18.1%, 79.5%) | -4.5 (-5.3, -3.7) | 106.5 (91.4, 121.7) | 91.8% (89.4%, 94.2%) | 1.4% (0.4%, 2.4%) |
| | | | Medium | 48% (25.1%, 65.6%) | -12 (-12.5, -11.4) | 185.7 (169.8, 201.6) | 45.4% (41%, 49.8%) | 0% (0%, 0%) |
| | | | High | 42% (24.2%, 60.2%) | -18 (-18.5, -17.4) | 358.3 (338.5, 378.2) | 6.2% (4.1%, 8.3%) | 0% (0%, 0%) |

| | | | | | | | | |
|---|---|---|---|---|---|---|---|---|
| | | Yes | Low | 58.8% (25.8%, 84.4%) | -1.2 (-2, -0.5) | 83.5 (71.7, 95.3) | 95.6% (93.8%, 97.4%) | 1% (0.1%, 1.9%) |
| | | | Medium | 58% (40.2%, 72.3%) | -2 (-2.5, -1.6) | 30.6 (26.4, 34.8) | 94.8% (92.9%, 96.7%) | 0% (0%, 0%) |
| | | | High | 55.7% (38.9%, 70.2%) | -4.3 (-4.7, -3.9) | 44.2 (38.6, 49.7) | 85.6% (82.5%, 88.7%) | 0% (0%, 0%) |
| | Inferior | No | Low | 57.1% (25.1%, 80.7%) | -2.9 (-3.7, -2.1) | 91 (78.2, 103.9) | 93.4% (91.2%, 95.6%) | 1% (0.1%, 1.9%) |
| | | | Medium | 54.1% (35.1%, 69.3%) | -5.9 (-6.4, -5.5) | 63.8 (56.5, 71.1) | 81.8% (78.4%, 85.2%) | 0% (0%, 0%) |
| | | | High | 50.7% (34.9%, 66%) | -9.3 (-9.7, -8.9) | 107.3 (99, 115.6) | 42.4% (38.1%, 46.7%) | 0% (0%, 0%) |
| | | Yes | Low | 58.8% (26.8%, 81.8%) | -1.2 (-2, -0.4) | 80.4 (69.7, 91.1) | 95.4% (93.6%, 97.2%) | 0.8% (0%, 1.6%) |
| | | | Medium | 58.2% (39.2%, 73.1%) | -1.8 (-2.2, -1.3) | 28.9 (24.9, 32.9) | 94.2% (92.2%, 96.2%) | 0% (0%, 0%) |
| | | | High | 56.7% (40%, 69.9%) | -3.3 (-3.6, -2.9) | 30 (25.9, 34) | 87.8% (84.9%, 90.7%) | 0% (0%, 0%) |

**Supplemental Table 7. Mean estimate, bias, mean squared error (MSE), coverage, and false negative rate for vaccine efficacy (VE) estimates against severe *Shigella* diarrhea from highly powered trials for all 24 simulation scenarios when using active surveillance for infection and a crude recurrent outcome regression model.** Hybrid immunity was either not possible (no) or possible such that the first post-vaccination infection conferred additional immunity (yes). Bias was calculated as the average VE estimate for a set of simulations minus the true VE (60%). MSE was calculated as the average of the square of the estimate minus the true parameter. Coverage represents the percent of 95% confidence intervals (CIs) that contained the true VE (60%), and the false negative rate indicates the percent of 95% CIs that incorrectly indicated a non-significant effect.

| VE against infection | Strength of infection-acquired immunity | Hybrid immunity | Force of infection | Mean VE estimate (range) | Bias (95% CI) | MSE (95% CI) | Coverage (95% CI) | False negatives (95% CI) |
|---|---|---|---|---|---|---|---|---|
| 0% | Similar | No | Low | 54.5% (19.3%, 75.9%) | -5.5 (-6.4, -4.6) | 134.3 (115, 153.6) | 90.2% (87.6%, 92.8%) | 2.8% (1.4%, 4.2%) |
| | | | Medium | 47.3% (30.1%, 63.1%) | -12.7 (-13.2, -12.1) | 195.6 (181.6, 209.7) | 38% (33.7%, 42.3%) | 0% (0%, 0%) |
| | | | High | 40.4% (18.3%, 57.4%) | -19.6 (-20.1, -19.1) | 416.9 (396.2, 437.6) | 2.6% (1.2%, 4%) | 0% (0%, 0%) |
| | | Yes | Low | 59% (25.9%, 79.8%) | -1 (-1.7, -0.2) | 82.7 (71.7, 93.8) | 96.2% (94.5%, 97.9%) | 0.8% (0%, 1.6%) |
| | | | Medium | 57.7% (39.3%, 70.6%) | -2.3 (-2.8, -1.8) | 34.9 (30.4, 39.5) | 93.8% (91.7%, 95.9%) | 0% (0%, 0%) |
| | | | High | 55.4% (39%, 66.5%) | -4.6 (-5, -4.2) | 44.4 (38.8, 50.1) | 84.8% (81.7%, 87.9%) | 0% (0%, 0%) |
| | Inferior | No | Low | 57.2% (25.4%, 80.4%) | -2.8 (-3.6, -1.9) | 105.6 (89.9, 121.4) | 91.4% (88.9%, 93.9%) | 2.2% (0.9%, 3.5%) |
| | | | Medium | 53.3% (36.2%, 66.8%) | -6.7 (-7.1, -6.2) | 72.8 (65.3, 80.3) | 77.8% (74.2%, 81.4%) | 0% (0%, 0%) |
| | | | High | 49.2% (33%, 60.7%) | -10.8 (-11.2, -10.4) | 140.1 (130, 150.3) | 28.6% (24.6%, 32.6%) | 0% (0%, 0%) |
| | | Yes | Low | 59.2% (24.1%, 80.7%) | -0.8 (-1.7, 0) | 87.8 (75.4, 100.3) | 93.2% (91%, 95.4%) | 0.8% (0%, 1.6%) |
| | | | Medium | 58.1% (41.9%, 72.3%) | -1.9 (-2.3, -1.5) | 28.3 (24.7, 31.9) | 94.4% (92.4%, 96.4%) | 0% (0%, 0%) |
| | | | High | 56.2% (41%, 66.4%) | -3.8 (-4.2, -3.5) | 32.9 (28.8, 37.1) | 85.2% (82.1%, 88.3%) | 0% (0%, 0%) |
| 20% | Similar | No | Low | 55.1% (18%, 79.6%) | -4.9 (-5.7, -4.1) | 110.5 (95, 126) | 91.8% (89.4%, 94.2%) | 1.8% (0.6%, 3%) |
| | | | Medium | 47.1% (23.9%, 65.3%) | -12.9 (-13.5, -12.3) | 209.6 (192.6, 226.6) | 38.4% (34.1%, 42.7%) | 0% (0%, 0%) |
| | | | High | 40.9% (22.9%, 59.3%) | -19.1 (-19.6, -18.5) | 400.2 (379.1, 421.4) | 3.8% (2.1%, 5.5%) | 0% (0%, 0%) |

| | | | | | | | | |
|---|---|---|---|---|---|---|---|---|
| | | Yes | Low | 58.2%<br>(25.4%, 84.2%) | -1.8<br>(-2.6, -1) | 86.8<br>(74.4, 99.2) | 95.2%<br>(93.3%, 97.1%) | 1.4%<br>(0.4%, 2.4%) |
| | | | Medium | 56.6%<br>(37.9%, 71.5%) | -3.4<br>(-3.9, -3) | 39.6<br>(34.4, 44.9) | 93.2%<br>(91%, 95.4%) | 0%<br>(0%, 0%) |
| | | | High | 53.8%<br>(36.6%, 68.7%) | -6.2<br>(-6.6, -5.7) | 65.5<br>(58.2, 72.8) | 73.2%<br>(69.3%, 77.1%) | 0%<br>(0%, 0%) |
| | Inferior | No | Low | 56.9%<br>(26.2%, 80.6%) | -3.1<br>(-3.9, -2.3) | 91.6<br>(79, 104.2) | 93.8%<br>(91.7%, 95.9%) | 1%<br>(0.1%, 1.9%) |
| | | | Medium | 53.5%<br>(34.5%, 68.7%) | -6.5<br>(-7, -6.1) | 71.6<br>(63.8, 79.4) | 77.6%<br>(73.9%, 81.3%) | 0%<br>(0%, 0%) |
| | | | High | 49.8%<br>(34.7%, 64.9%) | -10.2<br>(-10.6, -9.8) | 124.5<br>(115.5, 133.5) | 34.6%<br>(30.4%, 38.8%) | 0%<br>(0%, 0%) |
| | | Yes | Low | 58.5%<br>(29.3%, 81.8%) | -1.5<br>(-2.3, -0.7) | 80.8<br>(70.4, 91.2) | 95.2%<br>(93.3%, 97.1%) | 0.6%<br>(-0.1%, 1.3%) |
| | | | Medium | 57.5%<br>(38.3%, 72.3%) | -2.5<br>(-2.9, -2) | 32.8<br>(28.3, 37.2) | 93%<br>(90.8%, 95.2%) | 0%<br>(0%, 0%) |
| | | | High | 55.6%<br>(38.2%, 69%) | -4.4<br>(-4.8, -4) | 38.9<br>(34.2, 43.6) | 82.4%<br>(79.1%, 85.7%) | 0%<br>(0%, 0%) |

**Supplemental Table 8. Mean estimate, bias, mean squared error (MSE), coverage, and false negative rate for vaccine efficacy (VE) estimates against severe *Shigella* diarrhea from highly powered trials for all 24 simulation scenarios when using symptom-based reporting and a crude recurrent outcome regression model.** Hybrid immunity was either not possible (no) or possible such that the first post-vaccination infection conferred additional immunity (yes). Bias was calculated as the average VE estimate for a set of simulations minus the true VE (60%). MSE was calculated as the average of the square of the estimate minus the true parameter. Coverage represents the percent of 95% confidence intervals (CIs) that contained the true VE (60%), and the false negative rate indicates the percent of 95% CIs that incorrectly indicated a non-significant effect.

| VE against infection | Strength of infection-acquired immunity | Hybrid immunity | Force of infection | Mean VE estimate (range) | Bias (95% CI) | MSE (95% CI) | Coverage (95% CI) | False negatives (95% CI) |
|---|---|---|---|---|---|---|---|---|
| 0% | Similar | No | Low | 54.5% (19.3%, 75.9%) | -5.5 (-6.4, -4.6) | 134.2 (114.9, 153.5) | 90.2% (87.6%, 92.8%) | 2.8% (1.4%, 4.2%) |
| | | | Medium | 47.3% (30.1%, 63.1%) | -12.7 (-13.2, -12.1) | 195.2 (181.2, 209.2) | 38.2% (33.9%, 42.5%) | 0% (0%, 0%) |
| | | | High | 40.4% (18.3%, 57.5%) | -19.6 (-20.1, -19.1) | 416 (395.3, 436.6) | 2.6% (1.2%, 4%) | 0% (0%, 0%) |
| | | Yes | Low | 59.1% (25.9%, 79.8%) | -0.9 (-1.7, -0.2) | 82.7 (71.7, 93.7) | 96.2% (94.5%, 97.9%) | 0.8% (0%, 1.6%) |
| | | | Medium | 57.7% (39.3%, 70.7%) | -2.3 (-2.8, -1.8) | 34.8 (30.3, 39.4) | 94% (91.9%, 96.1%) | 0% (0%, 0%) |
| | | | High | 55.5% (39.1%, 66.6%) | -4.5 (-5, -4.1) | 44.2 (38.6, 49.8) | 84.8% (81.7%, 87.9%) | 0% (0%, 0%) |
| | Inferior | No | Low | 57.2% (25.4%, 80.4%) | -2.8 (-3.6, -1.9) | 105.6 (89.9, 121.3) | 91.4% (88.9%, 93.9%) | 2.2% (0.9%, 3.5%) |
| | | | Medium | 53.4% (36.2%, 66.8%) | -6.6 (-7.1, -6.2) | 72.5 (65, 79.9) | 77.8% (74.2%, 81.4%) | 0% (0%, 0%) |
| | | | High | 49.2% (33%, 60.8%) | -10.8 (-11.2, -10.4) | 139.3 (129.2, 149.4) | 28.8% (24.8%, 32.8%) | 0% (0%, 0%) |
| | | Yes | Low | 59.2% (24.1%, 80.7%) | -0.8 (-1.7, 0) | 87.8 (75.3, 100.2) | 93.2% (91%, 95.4%) | 0.8% (0%, 1.6%) |
| | | | Medium | 58.1% (41.9%, 72.3%) | -1.9 (-2.3, -1.4) | 28.2 (24.6, 31.8) | 94.4% (92.4%, 96.4%) | 0% (0%, 0%) |
| | | | High | 56.2% (41%, 66.4%) | -3.8 (-4.2, -3.4) | 32.6 (28.5, 36.8) | 85.6% (82.5%, 88.7%) | 0% (0%, 0%) |
| 20% | Similar | No | Low | 55.1% (18%, 79.5%) | -4.9 (-5.7, -4.1) | 110.6 (95.1, 126.1) | 91.8% (89.4%, 94.2%) | 1.8% (0.6%, 3%) |
| | | | Medium | 47.1% (23.9%, 65.3%) | -12.9 (-13.5, -12.3) | 209.7 (192.7, 226.8) | 38.2% (33.9%, 42.5%) | 0% (0%, 0%) |
| | | | High | 40.9% (22.9%, 59.3%) | -19.1 (-19.6, -18.5) | 400.6 (379.4, 421.8) | 3.8% (2.1%, 5.5%) | 0% (0%, 0%) |

| | | | | | | | | |
|---|---|---|---|---|---|---|---|---|
| | | Yes | Low | 58.2%<br>(25.4%, 84.2%) | -1.8<br>(-2.6, -1) | 86.8<br>(74.4, 99.3) | 95.2%<br>(93.3%, 97.1%) | 1.4%<br>(0.4%, 2.4%) |
| | | | Medium | 56.6%<br>(37.9%, 71.5%) | -3.4<br>(-3.9, -3) | 39.7<br>(34.5, 44.9) | 93.2%<br>(91%, 95.4%) | 0%<br>(0%, 0%) |
| | | | High | 53.8%<br>(36.6%, 68.7%) | -6.2<br>(-6.6, -5.7) | 65.7<br>(58.3, 73) | 73.2%<br>(69.3%, 77.1%) | 0%<br>(0%, 0%) |
| | Inferior | No | Low | 56.9%<br>(26.2%, 80.6%) | -3.1<br>(-3.9, -2.4) | 91.6<br>(79, 104.2) | 93.8%<br>(91.7%, 95.9%) | 1%<br>(0.1%, 1.9%) |
| | | | Medium | 53.5%<br>(34.5%, 68.7%) | -6.5<br>(-7, -6.1) | 71.7<br>(63.9, 79.6) | 77.6%<br>(73.9%, 81.3%) | 0%<br>(0%, 0%) |
| | | | High | 49.8%<br>(34.6%, 64.8%) | -10.2<br>(-10.6, -9.8) | 125<br>(116, 134.1) | 34.2%<br>(30%, 38.4%) | 0%<br>(0%, 0%) |
| | | Yes | Low | 58.5%<br>(29.3%, 81.8%) | -1.5<br>(-2.3, -0.7) | 80.8<br>(70.4, 91.2) | 95.2%<br>(93.3%, 97.1%) | 0.6%<br>(-0.1%, 1.3%) |
| | | | Medium | 57.5%<br>(38.3%, 72.3%) | -2.5<br>(-2.9, -2) | 32.8<br>(28.4, 37.3) | 92.8%<br>(90.5%, 95.1%) | 0%<br>(0%, 0%) |
| | | | High | 55.6%<br>(38.2%, 68.9%) | -4.4<br>(-4.8, -4) | 39.1<br>(34.4, 43.9) | 82.2%<br>(78.8%, 85.6%) | 0%<br>(0%, 0%) |

**Supplemental Table 9. Mean estimate, bias, mean squared error (MSE), coverage, and false negative rate for vaccine efficacy (VE) estimates against severe *Shigella* diarrhea from highly powered trials for all 6 design approaches applied to stratified datasets for the first and second year of follow-up. 24 simulation scenarios when using symptom-based reporting and a crude recurrent outcome regression model.** These results are for scenarios where VE against infection was 0%, infection- and vaccine-acquired immunity were comparable, and hybrid immunity was not possible. Bias was calculated as the average VE estimate for a set of simulations minus the true VE (60%). MSE was calculated as the average of the square of the estimate minus the true parameter. Coverage represents the percent of 95% confidence intervals (CIs) that contained the true VE (60%), and the false negative rate indicates the percent of 95% CIs that incorrectly indicated a non-significant effect.

| Regression model type | Data collection approach | Force of infection | Year of life | Mean VE estimate (range) | Bias (95% CI) | MSE (95% CI) | Coverage (95% CI) | False negatives (95% CI) |
|---|---|---|---|---|---|---|---|---|
| Single outcome | Active surveillance | Low | First | 58.8% (11.6%, 80.6%) | -1.2 (-2.1, -0.3) | 104.4 (87.9, 120.9) | 94.4% (92.4%, 96.4%) | 2.4% (1.1%, 3.7%) |
| | | | Second | 57.9% (-24.4%, 93.5%) | -2.1 (-3.7, -0.4) | 364.4 (302.2, 426.5) | 95.6% (93.8%, 97.4%) | 37.4% (33.2%, 41.6%) |
| | | Medium | First | 59.5% (42.8%, 74.1%) | -0.5 (-1, 0.1) | 38 (33.7, 42.4) | 95.8% (94%, 97.6%) | 0% (0%, 0%) |
| | | | Second | 59.4% (-2.6%, 85.9%) | -0.6 (-1.8, 0.6) | 181.6 (151.4, 211.8) | 97.2% (95.8%, 98.6%) | 13.2% (10.2%, 16.2%) |
| | | High | First | 59.6% (42.6%, 74%) | -0.4 (-0.8, 0.1) | 28.7 (25.2, 32.2) | 95% (93.1%, 96.9%) | 0% (0%, 0%) |
| | | | Second | 58.2% (13.4%, 91.3%) | -1.8 (-3.1, -0.5) | 221.1 (191.9, 250.3) | 93.6% (91.5%, 95.7%) | 21.6% (18%, 25.2%) |
| | Symptom-based reporting | Low | First | 55.9% (18.3%, 79.1%) | -4.1 (-5, -3.2) | 127.6 (108.7, 146.6) | 92.4% (90.1%, 94.7%) | 3.2% (1.7%, 4.7%) |
| | | | Second | 52.2% (-30.9%, 86.9%) | -7.8 (-9.6, -6.1) | 458.3 (375.6, 541) | 92.8% (90.5%, 95.1%) | 41.4% (37.1%, 45.7%) |
| | | Medium | First | 51.9% (31.4%, 70%) | -8.1 (-8.6, -7.6) | 103.1 (92.6, 113.6) | 73.2% (69.3%, 77.1%) | 0% (0%, 0%) |
| | | | Second | 43.7% (-34.4%, 77.3%) | -16.3 (-17.5, -15.1) | 454.2 (389.7, 518.7) | 75% (71.2%, 78.8%) | 28.2% (24.3%, 32.1%) |
| | | High | First | 47.9% (31.1%, 65.7%) | -12.1 (-12.6, -11.6) | 179 (166.3, 191.7) | 32.6% (28.5%, 36.7%) | 0% (0%, 0%) |
| | | | Second | 32.6% (-45.5%, 70.3%) | -27.4 (-28.7, -26) | 979.8 (885, 1074.6) | 33% (28.9%, 37.1%) | 48% (43.6%, 52.4%) |
| | Active surveillance | Low | First | 55.7% (18.6%, 80%) | -4.3 (-5.2, -3.3) | 127.8 (108.4, 147.3) | 92.4% (90.1%, 94.7%) | 3.2% (1.7%, 4.7%) |

| | | | | | | | | |
|---|---|---|---|---|---|---|---|---|
| Stratified recurrent outcome | | | Second | 49.8% (-33.6%, 86.9%) | -10.2 (-11.9, -8.4) | 503.7 (415, 592.4) | 91.4% (88.9%, 93.9%) | 46.6% (42.2%, 51%) |
| | | Medium | First | 50.7% (32.7%, 65.2%) | -9.3 (-9.8, -8.8) | 124.7 (113.1, 136.3) | 68% (63.9%, 72.1%) | 0% (0%, 0%) |
| | | | Second | 36.7% (-21.5%, 70.1%) | -23.3 (-24.5, -22.2) | 717.4 (649.3, 785.5) | 45.8% (41.4%, 50.2%) | 39% (34.7%, 43.3%) |
| | | High | First | 45.5% (25%, 62.3%) | -14.5 (-15, -14) | 243.5 (227.7, 259.4) | 19% (15.6%, 22.4%) | 0% (0%, 0%) |
| | | | Second | 22.8% (-34.3%, 57.3%) | -37.2 (-38.5, -36) | 1,602.9 (1,491.5, 1,714.2) | 6.6% (4.4%, 8.8%) | 63.2% (59%, 67.4%) |
| | Symptom-based reporting | Low | First | 55.2% (15.3%, 78.2%) | -4.8 (-5.8, -3.9) | 136.2 (115.6, 156.7) | 91.2% (88.7%, 93.7%) | 3.6% (2%, 5.2%) |
| | | | Second | 50.5% (-33.1%, 87.3%) | -9.5 (-11.3, -7.8) | 488.1 (400.3, 575.8) | 92.2% (89.8%, 94.6%) | 44.2% (39.8%, 48.6%) |
| | | Medium | First | 49.4% (28.5%, 66.2%) | -10.6 (-11.2, -10.1) | 150.7 (137.8, 163.5) | 57% (52.7%, 61.3%) | 0% (0%, 0%) |
| | | | Second | 38.2% (-20.4%, 71.9%) | -21.8 (-22.9, -20.6) | 641.9 (578.1, 705.8) | 52% (47.6%, 56.4%) | 37.8% (33.5%, 42.1%) |
| | | High | First | 43.5% (23%, 62.7%) | -16.5 (-17, -15.9) | 304.6 (287.1, 322.1) | 10.6% (7.9%, 13.3%) | 0% (0%, 0%) |
| | | | Second | 24.4% (-35.9%, 58.2%) | -35.6 (-36.8, -34.3) | 1,474 (1,369.1, 1,578.8) | 9.4% (6.8%, 12%) | 58.8% (54.5%, 63.1%) |
| Crude recurrent outcome | Active surveillance | Low | First | 55.7% (18.5%, 80%) | -4.3 (-5.2, -3.3) | 127.8 (108.3, 147.2) | 92.4% (90.1%, 94.7%) | 3.2% (1.7%, 4.7%) |
| | | | Second | 49.8% (-33.3%, 87%) | -10.2 (-11.9, -8.4) | 502.8 (414.2, 591.5) | 91.4% (88.9%, 93.9%) | 45.8% (41.4%, 50.2%) |
| | | Medium | First | 50.7% (32.7%, 65.2%) | -9.3 (-9.8, -8.8) | 124.5 (112.9, 136.1) | 68.4% (64.3%, 72.5%) | 0% (0%, 0%) |
| | | | Second | 36.7% (-21%, 70.1%) | -23.3 (-24.5, -22.1) | 715.7 (647.8, 783.7) | 45.6% (41.2%, 50%) | 39.4% (35.1%, 43.7%) |
| | | High | First | 45.5% (25.1%, 62.6%) | -14.5 (-15, -14) | 244.1 (228.3, 259.8) | 18.8% (15.4%, 22.2%) | 0% (0%, 0%) |
| | | | Second | 22.7% (-34%, 57.6%) | -37.3 (-38.5, -36) | 1,603.8 (1,492.6, 1,714.9) | 6.6% (4.4%, 8.8%) | 63.2% (59%, 67.4%) |
| | Symptom-based reporting | Low | First | 54.8% (15%, 78.5%) | -5.2 (-6.1, -4.2) | 139.3 (118.5, 160.1) | 91% (88.5%, 93.5%) | 3.8% (2.1%, 5.5%) |

| | | | | | | | | |
|---|---|---|---|---|---|---|---|---|
| | | | Second | 49.8% (-33.3%, 87%) | -10.2 (-11.9, -8.4) | 502.6 (414, 591.2) | 91.4% (88.9%, 93.9%) | 45.8% (41.4%, 50.2%) |
| | | Medium | First | 48.6% (28.3%, 65.9%) | -11.4 (-12, -10.9) | 169.5 (155.8, 183.3) | 50.8% (46.4%, 55.2%) | 0% (0%, 0%) |
| | | | Second | 36.7% (-20.9%, 70.2%) | -23.3 (-24.4, -22.1) | 715.1 (647.2, 783) | 45.6% (41.2%, 50%) | 39.4% (35.1%, 43.7%) |
| | | High | First | 42.4% (21.2%, 61.8%) | -17.6 (-18.1, -17.1) | 342.9 (324.1, 361.6) | 7% (4.8%, 9.2%) | 0% (0%, 0%) |
| | | | Second | 22.8% (-34%, 57.6%) | -37.2 (-38.5, -36) | 1,602.6 (1,491.5, 1,713.7) | 6.6% (4.4%, 8.8%) | 63.2% (59%, 67.4%) |

**Supplemental Table 10. Mean size of each trial arm, estimate, bias, mean squared error (MSE), coverage, and false negative rate for vaccine efficacy (VE) estimates against severe *Shigella* diarrhea from realistically sized trials for all 24 simulation scenarios when using active surveillance for infection and a single outcome regression model.** Hybrid immunity was either not possible (no) or possible such that the first post-vaccination infection conferred additional immunity (yes). Bias was calculated as the average VE estimate for a set of simulations minus the true VE (60%). MSE was calculated as the average of the square of the estimate minus the true parameter. Coverage represents the percent of 95% confidence intervals (CIs) that contained the true VE (60%), and the false negative rate indicates the percent of 95% CIs that incorrectly indicated a non-significant effect.

| VE against infection | Strength of infection-acquired immunity | Hybrid immunity | Force of infection | Mean N (range) | Mean VE estimate (range) | Bias (95% CI) | MSE (95% CI) | Coverage (95% CI) | False negatives (95% CI) |
|---|---|---|---|---|---|---|---|---|---|
| 0% | Similar | No | Low | 2,251 (1,553, 3,745) | 57.6% (0.9%, 90.6%) | -2.4 (-3.8, -1.1) | 247.7 (208.8, 286.7) | 94.2% (92.2%, 96.2%) | 22% (18.4%, 25.6%) |
| | | | Medium | 964 (721, 1,349) | 58% (-10.1%, 91.5%) | -2 (-3.4, -0.7) | 241.7 (203.7, 279.6) | 95.2% (93.3%, 97.1%) | 22.4% (18.7%, 26.1%) |
| | | | High | 749 (551, 992) | 57.4% (-6.9%, 89.6%) | -2.6 (-4, -1.1) | 270.3 (226, 314.6) | 93.6% (91.5%, 95.7%) | 24% (20.3%, 27.7%) |
| | | Yes | Low | 2230 (1,420, 3,924) | 57.2% (-34.2%, 91.8%) | -2.8 (-4.2, -1.5) | 245.6 (194.9, 296.2) | 95.8% (94%, 97.6%) | 24.2% (20.4%, 28%) |
| | | | Medium | 976 (710, 1,350) | 59% (14.6%, 93.3%) | -1 (-2.2, 0.3) | 205.3 (177.8, 232.7) | 96.4% (94.8%, 98%) | 20.8% (17.2%, 24.4%) |
| | | | High | 752 (558, 1,028) | 58.8% (-1.7%, 88.8%) | -1.2 (-2.5, 0.2) | 240.7 (204.8, 276.6) | 96% (94.3%, 97.7%) | 22.8% (19.1%, 26.5%) |
| | Inferior | No | Low | 2238 (1,444, 3,583) | 57.8% (-62.6%, 94.3%) | -2.2 (-3.8, -0.7) | 313.4 (229.4, 397.4) | 94% (91.9%, 96.1%) | 21.4% (17.8%, 25%) |
| | | | Medium | 968 (721, 1,307) | 58.5% (-19.3%, 88.3%) | -1.5 (-2.9, -0.2) | 242 (198.5, 285.5) | 93.8% (91.7%, 95.9%) | 21% (17.4%, 24.6%) |
| | | | High | 748 (596, 991) | 57.4% (6.1%, 88.8%) | -2.6 (-3.8, -1.3) | 219.7 (187.8, 251.7) | 96.2% (94.5%, 97.9%) | 23.2% (19.5%, 26.9%) |
| | | Yes | Low | 2,237 (1,498, 3,583) | 57.9% (-50.1%, 94.1%) | -2.1 (-3.6, -0.6) | 291.2 (218.8, 363.6) | 93.2% (91%, 95.4%) | 20.8% (17.2%, 24.4%) |
| | | | Medium | 968 (747, 1372) | 57.9% (-15.4%, 89.2%) | -2.1 (-3.5, -0.8) | 235.5 (192, 279) | 95.8% (94%, 97.6%) | 22.2% (18.6%, 25.8%) |
| | | | High | 749 (559, 1,003) | 58.1% (-15.4%, 93.7%) | -1.9 (-3.1, -0.6) | 215.4 (178.8, 252) | 95.8% (94%, 97.6%) | 20.8% (17.2%, 24.4%) |
| 20% | Similar | No | Low | 2,235 (1,350, 3,745) | 57.6% (-4%, 93.5%) | -2.4 (-3.7, -1.1) | 225 (184.6, 265.3) | 96% (94.3%, 97.7%) | 20% (16.5%, 23.5%) |
| | | | Medium | 972 (690, 1,327) | 57.9% (-3.8%, 86%) | -2.1 (-3.5, -0.8) | 230.1 (190.5, 269.6) | 95.2% (93.3%, 97.1%) | 19.2% (15.7%, 22.7%) |
| | | | High | 750 (561, 1029) | 59% (-18.4%, 87.3%) | -1 (-2.2, 0.3) | 202 (166.4, 237.6) | 96% (94.3%, 97.7%) | 15.4% (12.2%, 18.6%) |

| | | | | | | | | | |
|---|---|---|---|---|---|---|---|---|---|
| | | Yes | Low | 2,229<br>(1,229, 3,433) | 57.6%<br>(-3.8%, 94.6%) | -2.4<br>(-3.7, -1.1) | 227.6<br>(190.9, 264.2) | 95.2%<br>(93.3%, 97.1%) | 21.4%<br>(17.8%, 25%) |
| | | | Medium | 973<br>(735, 1,395) | 58.5%<br>(-9.1%, 89.9%) | -1.5<br>(-2.8, -0.3) | 198.8<br>(163.1, 234.6) | 97.2%<br>(95.8%, 98.6%) | 18%<br>(14.6%, 21.4%) |
| | | | High | 750<br>(586, 1,084) | 58.1%<br>(-52.3%, 89.8%) | -1.9<br>(-3.2, -0.6) | 220.8<br>(163, 278.7) | 95.8%<br>(94%, 97.6%) | 17.2%<br>(13.9%, 20.5%) |
| | Inferior | No | Low | 2,249<br>(1,471, 4,847) | 57.7%<br>(-4.2%, 92.5%) | -2.3<br>(-3.7, -1) | 240.2<br>(206, 274.4) | 94.6%<br>(92.6%, 96.6%) | 22%<br>(18.4%, 25.6%) |
| | | | Medium | 971<br>(721, 1,374) | 58.7%<br>(1%, 87.5%) | -1.3<br>(-2.5, 0) | 211.7<br>(177.1, 246.3) | 94.4%<br>(92.4%, 96.4%) | 17.4%<br>(14.1%, 20.7%) |
| | | | High | 744<br>(544, 1,028) | 59%<br>(-1.5%, 91.1%) | -1<br>(-2.3, 0.2) | 207.2<br>(174.2, 240.3) | 94%<br>(91.9%, 96.1%) | 18%<br>(14.6%, 21.4%) |
| | | Yes | Low | 2,244<br>(1498, 4,847) | 58%<br>(2.2%, 88.5%) | -2<br>(-3.4, -0.7) | 239.5<br>(205.1, 273.9) | 95%<br>(93.1%, 96.9%) | 21.2%<br>(17.6%, 24.8%) |
| | | | Medium | 971<br>(715, 1,349) | 58.1%<br>(-3.6%, 88.2%) | -1.9<br>(-3.3, -0.6) | 239<br>(196.1, 281.9) | 93.4%<br>(91.2%, 95.6%) | 17.4%<br>(14.1%, 20.7%) |
| | | | High | 743<br>(523, 1,111) | 58.8%<br>(6.5%, 90.5%) | -1.2<br>(-2.4, 0) | 188.4<br>(159.7, 217.1) | 95%<br>(93.1%, 96.9%) | 17.6%<br>(14.3%, 20.9%) |

**Supplemental Table 11. Mean estimate, bias, mean squared error (MSE), coverage, and false negative rate for vaccine efficacy (VE) estimates against severe *Shigella* diarrhea from realistically sized trials for all 24 simulation scenarios when using symptom-based reporting and a single outcome regression model.** Hybrid immunity was either not possible (no) or possible such that the first post-vaccination infection conferred additional immunity (yes). Bias was calculated as the average VE estimate for a set of simulations minus the true VE (60%). MSE was calculated as the average of the square of the estimate minus the true parameter. Coverage represents the percent of 95% confidence intervals (CIs) that contained the true VE (60%), and the false negative rate indicates the percent of 95% CIs that incorrectly indicated a non-significant effect.

| VE against infection | Strength of infection-acquired immunity | Hybrid immunity | Force of infection | Mean N (range) | Mean VE estimate (range) | Bias (95% CI) | MSE (95% CI) | Coverage (95% CI) | False negatives (95% CI) |
|---|---|---|---|---|---|---|---|---|---|
| 0% | Similar | No | Low | 2,181 (1,497, 3,745) | 54.8% (-4.9%, 88.3%) | -5.2 (-6.7, -3.8) | 288.4 (241.8, 335) | 93.6% (91.5%, 95.7%) | 25.6% (21.8%, 29.4%) |
| | | | Medium | 856 (662, 1,128) | 49.3% (-16.3%, 82.7%) | -10.7 (-12.2, -9.3) | 404.1 (338.1, 470.1) | 87.6% (84.7%, 90.5%) | 34% (29.8%, 38.2%) |
| | | | High | 618 (474, 814) | 44.4% (-64.4%, 84.4%) | -15.6 (-17.4, -13.8) | 650.8 (525.3, 776.3) | 82.2% (78.8%, 85.6%) | 42.2% (37.9%, 46.5%) |
| | | Yes | Low | 2,164 (1,420, 3,745) | 58% (-36.2%, 92.4%) | -2 (-3.4, -0.7) | 236.2 (186.1, 286.3) | 95.2% (93.3%, 97.1%) | 22% (18.4%, 25.6%) |
| | | | Medium | 883 (653, 1,193) | 58.9% (13.4%, 93%) | -1.1 (-2.3, 0.1) | 189.6 (163, 216.2) | 95.4% (93.6%, 97.2%) | 17.6% (14.3%, 20.9%) |
| | | | High | 652 (469, 831) | 57.7% (-33.4%, 91%) | -2.3 (-3.7, -1) | 244.9 (197.1, 292.7) | 95.2% (93.3%, 97.1%) | 20.6% (17.1%, 24.1%) |
| | Inferior | No | Low | 2,090 (1,419, 3,433) | 57% (-24.3%, 89%) | -3 (-4.5, -1.6) | 273.6 (218.7, 328.5) | 94.8% (92.9%, 96.7%) | 23.8% (20.1%, 27.5%) |
| | | | Medium | 795 (627, 1,083) | 54% (-41.5%, 86.9%) | -6 (-7.4, -4.6) | 289.4 (230.5, 348.3) | 93.4% (91.2%, 95.6%) | 26.6% (22.7%, 30.5%) |
| | | | High | 561 (441, 702) | 51.6% (-19%, 84.9%) | -8.4 (-9.8, -7) | 328.4 (267.7, 389) | 91.8% (89.4%, 94.2%) | 29% (25%, 33%) |
| | | Yes | Low | 2,095 (1,444, 3,433) | 58.5% (-15.3%, 90.8%) | -1.5 (-2.8, -0.1) | 241.7 (197.2, 286.3) | 94.2% (92.2%, 96.2%) | 20% (16.5%, 23.5%) |
| | | | Medium | 796 (637, 1,159) | 57.3% (-4.4%, 88.4%) | -2.7 (-4, -1.4) | 226.4 (188.9, 264) | 95.8% (94%, 97.6%) | 21.4% (17.8%, 25%) |
| | | | High | 565 (443, 734) | 57.3% (2.6%, 85.2%) | -2.7 (-3.9, -1.5) | 193.3 (164.3, 222.3) | 96% (94.3%, 97.7%) | 20.2% (16.7%, 23.7%) |
| 20% | Similar | No | Low | 2,164 (1,328, 3,745) | 54.8% (-47.8%, 86.9%) | -5.2 (-6.5, -3.8) | 272.6 (211.6, 333.7) | 95% (93.1%, 96.9%) | 24.8% (21%, 28.6%) |
| | | | Medium | 861 (642, 1,176) | 49.3% (-26.9%, 88.6%) | -10.7 (-12.2, -9.2) | 394.1 (326.7, 461.4) | 89.2% (86.5%, 91.9%) | 33.8% (29.7%, 37.9%) |
| | | | High | 618 (500, 783) | 45.9% (-64.5%, 82.6%) | -14.1 (-15.6, -12.6) | 502.5 (411.3, 593.8) | 84.8% (81.7%, 87.9%) | 39.2% (34.9%, 43.5%) |

| | | | | | | | | | |
|---|---|---|---|---|---|---|---|---|---|
| | | Yes | Low | 2,158 (1,193, 3,296) | 57% (-12.3%, 94.3%) | -3 (-4.4, -1.7) | 244.1 (201.6, 286.6) | 93.6% (91.5%, 95.7%) | 24.2% (20.4%, 28%) |
| | | | Medium | 878 (673, 1,248) | 57% (-21.4%, 90.3%) | -3 (-4.3, -1.8) | 211 (168.9, 253.1) | 96.2% (94.5%, 97.9%) | 19.4% (15.9%, 22.9%) |
| | | | High | 641 (496, 839) | 55.2% (-12.3%, 89.7%) | -4.8 (-6.1, -3.4) | 251.8 (203.7, 299.9) | 94.2% (92.2%, 96.2%) | 21.8% (18.2%, 25.4%) |
| | Inferior | No | Low | 2,097 (1,349, 3,745) | 56.3% (-12.1%, 95.7%) | -3.7 (-5.1, -2.3) | 256.4 (212, 300.8) | 93.8% (91.7%, 95.9%) | 24.4% (20.6%, 28.2%) |
| | | | Medium | 797 (604, 1,083) | 54.2% (-6.2%, 87.6%) | -5.8 (-7.1, -4.4) | 266.8 (222.2, 311.4) | 93% (90.8%, 95.2%) | 24.8% (21%, 28.6%) |
| | | | High | 556 (445, 708) | 52.3% (-1.3%, 88%) | -7.7 (-9.1, -6.4) | 301 (255, 347) | 91.8% (89.4%, 94.2%) | 31.6% (27.5%, 35.7%) |
| | | Yes | Low | 2,096 (1,372, 3,924) | 57.6% (-4.2%, 91.8%) | -2.4 (-3.7, -1) | 241.2 (203.1, 279.3) | 93.6% (91.5%, 95.7%) | 20.6% (17.1%, 24.1%) |
| | | | Medium | 799 (609, 1,112) | 56.9% (-5.2%, 87%) | -3.1 (-4.4, -1.7) | 251.4 (205.4, 297.5) | 93.4% (91.2%, 95.6%) | 19.8% (16.3%, 23.3%) |
| | | | High | 560 (429, 768) | 56.6% (-1.1%, 87.1%) | -3.4 (-4.7, -2.2) | 211.1 (176.9, 245.3) | 94.8% (92.9%, 96.7%) | 21.8% (18.2%, 25.4%) |

**Supplemental Table 1. Mean estimate, bias, mean squared error (MSE), coverage, and false negative rate for vaccine efficacy (VE) estimates against severe *Shigella* diarrhea from realistically sized trials for all 24 simulation scenarios when using active surveillance for infection and a stratified recurrent outcome regression model.** Hybrid immunity was either not possible (no) or possible such that the first post-vaccination infection conferred additional immunity (yes). Bias was calculated as the average VE estimate for a set of simulations minus the true VE (60%). MSE was calculated as the average of the square of the estimate minus the true parameter. Coverage represents the percent of 95% confidence intervals (CIs) that contained the true VE (60%), and the false negative rate indicates the percent of 95% CIs that incorrectly indicated a non-significant effect.

| VE against infection | Strength of infection-acquired immunity | Hybrid immunity | Force of infection | Mean N (range) | Mean VE estimate (range) | Bias (95% CI) | MSE (95% CI) | Coverage (95% CI) | False negatives (95% CI) |
|---|---|---|---|---|---|---|---|---|---|
| 0% | Similar | No | Low | 2,103 (1,419, 3,580) | 53.5% (-8.4%, 88.3%) | -6.5 (-8, -5.1) | 313.7 (260.5, 366.9) | 93% (90.8%, 95.2%) | 30% (26%, 34%) |
| | | | Medium | 767 (600, 1043) | 44.7% (-22.6%, 80.4%) | -15.3 (-16.8, -13.7) | 554.9 (477, 632.8) | 82.6% (79.3%, 85.9%) | 45.8% (41.4%, 50.2%) |
| | | | High | 518 (403, 653) | 37.2% (-59.3%, 79.7%) | -22.8 (-24.7, -20.8) | 988.5 (834.3, 1142.7) | 71.6% (67.6%, 75.6%) | 58.2% (53.9%, 62.5%) |
| | | Yes | Low | 2,098 (1,395, 3,582) | 57.6% (-19.6%, 92.4%) | -2.4 (-3.7, -1.1) | 229.2 (187.1, 271.3) | 96.2% (94.5%, 97.9%) | 22.2% (18.6%, 25.8%) |
| | | | Medium | 808 (618, 1,055) | 56.6% (6.8%, 95.2%) | -3.4 (-4.7, -2.1) | 229.7 (195.7, 263.7) | 94% (91.9%, 96.1%) | 23.6% (19.9%, 27.3%) |
| | | | High | 573 (419, 716) | 53.9% (-28.8%, 89.2%) | -6.1 (-7.6, -4.6) | 332.8 (268.1, 397.4) | 91.6% (89.2%, 94%) | 27.6% (23.7%, 31.5%) |
| | Inferior | No | Low | 1,984 (1,328, 3,295) | 56.4% (-26.8%, 89.3%) | -3.6 (-5, -2.1) | 275.3 (225.1, 325.5) | 93.8% (91.7%, 95.9%) | 27% (23.1%, 30.9%) |
| | | | Medium | 682 (545, 895) | 51.4% (-13.7%, 86.4%) | -8.6 (-10, -7.2) | 338.9 (283.4, 394.4) | 91.6% (89.2%, 94%) | 33.8% (29.7%, 37.9%) |
| | | | High | 444 (368, 548) | 47% (-13.7%, 80.4%) | -13 (-14.5, -11.5) | 469.3 (403.7, 535) | 87.4% (84.5%, 90.3%) | 43.4% (39.1%, 47.7%) |
| | | Yes | Low | 1,991 (1,351, 3,295) | 58.4% (-21.5%, 90.6%) | -1.6 (-2.9, -0.2) | 242.9 (199.1, 286.8) | 94.4% (92.4%, 96.4%) | 21.6% (18%, 25.2%) |
| | | | Medium | 686 (555, 980) | 55.3% (-26.8%, 89.8%) | -4.7 (-6.1, -3.3) | 273.8 (222.5, 325.1) | 94% (91.9%, 96.1%) | 25.6% (21.8%, 29.4%) |
| | | | High | 451 (366, 552) | 54.4% (-16.2%, 85.5%) | -5.6 (-7, -4.2) | 274.4 (225.9, 322.9) | 93.8% (91.7%, 95.9%) | 27.2% (23.3%, 31.1%) |
| 20% | Similar | No | Low | 2,087 (1,287, 3,582) | 53.9% (-7.1%, 90.2%) | -6.1 (-7.4, -4.7) | 283.4 (235.4, 331.4) | 94% (91.9%, 96.1%) | 26.6% (22.7%, 30.5%) |
| | | | Medium | 772 (583, 1,083) | 46.1% (-25.7%, 87%) | -13.9 (-15.4, -12.3) | 500.4 (424.2, 576.7) | 84.6% (81.4%, 87.8%) | 44% (39.6%, 48.4%) |
| | | | High | 516 (407, 642) | 41% (-57.6%, 81.6%) | -19 (-20.7, -17.3) | 736.6 (618.2, 855) | 79.2% (75.6%, 82.8%) | 54.8% (50.4%, 59.2%) |

| | | | | | | | | | |
|---|---|---|---|---|---|---|---|---|---|
| | | Yes | Low | 2,093 (1,210, 3,169) | 57.3% (-8.2%, 94.3%) | -2.7 (-4, -1.4) | 233.3 (195.8, 270.8) | 95.2% (93.3%, 97.1%) | 23% (19.3%, 26.7%) |
| | | | Medium | 804 (632, 1,113) | 57.3% (-1.3%, 87.4%) | -2.7 (-3.9, -1.5) | 203.8 (167.8, 239.7) | 96.2% (94.5%, 97.9%) | 22.2% (18.6%, 25.8%) |
| | | | High | 561 (442, 710) | 55.4% (-16.1%, 84.2%) | -4.6 (-5.9, -3.3) | 241.4 (193.5, 289.3) | 96.6% (95%, 98.2%) | 23.4% (19.7%, 27.1%) |
| | Inferior | No | Low | 1,996 (1,307, 3,296) | 55.7% (-16.2%, 91.7%) | -4.3 (-5.6, -2.9) | 264.2 (217.1, 311.4) | 94.2% (92.2%, 96.2%) | 26% (22.2%, 29.8%) |
| | | | Medium | 683 (527, 895) | 52.4% (-15%, 89.1%) | -7.6 (-9, -6.1) | 326.5 (270, 383) | 92.8% (90.5%, 95.1%) | 31.4% (27.3%, 35.5%) |
| | | | High | 438 (350, 563) | 49.4% (-12.7%, 83.5%) | -10.6 (-12, -9.2) | 382.1 (326.8, 437.4) | 90.6% (88%, 93.2%) | 39% (34.7%, 43.3%) |
| | | Yes | Low | 1,994 (1,327, 3,433) | 57.5% (-3.5%, 88.1%) | -2.5 (-3.8, -1.1) | 240.6 (201.3, 279.9) | 93.4% (91.2%, 95.6%) | 22% (18.4%, 25.6%) |
| | | | Medium | 688 (527, 946) | 56.5% (-15.9%, 85.8%) | -3.5 (-4.9, -2) | 275.9 (224.1, 327.8) | 93.8% (91.7%, 95.9%) | 22% (18.4%, 25.6%) |
| | | | High | 448 (345, 587) | 55.5% (-14.7%, 87.2%) | -4.5 (-5.8, -3.2) | 240.3 (198.5, 282) | 95.2% (93.3%, 97.1%) | 24% (20.3%, 27.7%) |

**Supplemental Table 13. Mean estimate, bias, mean squared error (MSE), coverage, and false negative rate for vaccine efficacy (VE) estimates against severe *Shigella* diarrhea from realistically sized trials for all 24 simulation scenarios when using symptom-based reporting and a stratified recurrent outcome regression model.** Hybrid immunity was either not possible (no) or possible such that the first post-vaccination infection conferred additional immunity (yes). Bias was calculated as the average VE estimate for a set of simulations minus the true VE (60%). MSE was calculated as the average of the square of the estimate minus the true parameter. Coverage represents the percent of 95% confidence intervals (CIs) that contained the true VE (60%), and the false negative rate indicates the percent of 95% CIs that incorrectly indicated a non-significant effect.

| VE against infection | Strength of infection-acquired immunity | Hybrid immunity | Force of infection | Mean N (range) | Mean VE estimate (range) | Bias (95% CI) | MSE (95% CI) | Coverage (95% CI) | False negatives (95% CI) |
|---|---|---|---|---|---|---|---|---|---|
| 0% | Similar | No | Low | 2,151 (1,419, 3,744) | 53.7% (-9.2%, 88.5%) | -6.3 (-7.7, -4.9) | 304.1 (252.9, 355.3) | 93.2% (91%, 95.4%) | 28.6% (24.6%, 32.6%) |
| | | | Medium | 780 (613, 1,056) | 45.8% (-17.8%, 80.8%) | -14.2 (-15.8, -12.7) | 514.1 (441, 587.2) | 85% (81.9%, 88.1%) | 41.8% (37.5%, 46.1%) |
| | | | High | 528 (408, 674) | 38.1% (-76.8%, 78.7%) | -21.9 (-23.8, -20) | 942.9 (788.8, 1096.9) | 73.4% (69.5%, 77.3%) | 56.2% (51.9%, 60.5%) |
| | | Yes | Low | 2,141 (1,395, 3,582) | 57.8% (-20%, 90.2%) | -2.2 (-3.5, -0.9) | 223.7 (178.6, 268.7) | 96.4% (94.8%, 98%) | 21% (17.4%, 24.6%) |
| | | | Medium | 823 (622, 1,097) | 58.3% (9.1%, 95.4%) | -1.7 (-3, -0.5) | 201.2 (171.4, 231.1) | 95.2% (93.3%, 97.1%) | 19.8% (16.3%, 23.3%) |
| | | | High | 587 (421, 741) | 56.1% (-25.5%, 89.1%) | -3.9 (-5.3, -2.5) | 275.3 (222.2, 328.4) | 92.2% (89.8%, 94.6%) | 25% (21.2%, 28.8%) |
| | Inferior | No | Low | 2,021 (1,371, 3,295) | 56.6% (-26.6%, 89.3%) | -3.4 (-4.8, -2) | 268.9 (219.3, 318.4) | 94% (91.9%, 96.1%) | 24% (20.3%, 27.7%) |
| | | | Medium | 692 (559, 935) | 52.3% (-22.2%, 86.5%) | -7.7 (-9.2, -6.3) | 320.3 (263.3, 377.3) | 92.4% (90.1%, 94.7%) | 31.2% (27.1%, 35.3%) |
| | | | High | 452 (370, 551) | 48.1% (-5.2%, 81.9%) | -11.9 (-13.4, -10.4) | 432.5 (371.5, 493.5) | 87.8% (84.9%, 90.7%) | 39.6% (35.3%, 43.9%) |
| | | Yes | Low | 2,028 (1,419, 3,295) | 58.7% (-12.1%, 90.9%) | -1.3 (-2.7, 0) | 234.8 (196.5, 273.1) | 94% (91.9%, 96.1%) | 20.8% (17.2%, 24.4%) |
| | | | Medium | 697 (559, 1,003) | 56.4% (-23.1%, 90.2%) | -3.6 (-5, -2.3) | 254.2 (203, 305.4) | 94.4% (92.4%, 96.4%) | 23.4% (19.7%, 27.1%) |
| | | | High | 459 (371, 563) | 55.7% (-11%, 86.8%) | -4.3 (-5.7, -3) | 248.1 (204, 292.3) | 94.4% (92.4%, 96.4%) | 24.6% (20.8%, 28.4%) |
| 20% | Similar | No | Low | 2,132 (1,349, 3,582) | 53.7% (-10.3%, 90.2%) | -6.3 (-7.7, -4.9) | 284 (235.9, 332) | 94.2% (92.2%, 96.2%) | 26.4% (22.5%, 30.3%) |
| | | | Medium | 784 (591, 1,069) | 45.6% (-23.8%, 87.3%) | -14.4 (-15.9, -12.9) | 508.5 (434.4, 582.6) | 84.4% (81.2%, 87.6%) | 45.6% (41.2%, 50%) |
| | | | High | 526 (411, 668) | 40% (-65.4%, 81.1%) | -20 (-21.7, -18.2) | 787.4 (657.1, 917.6) | 76.4% (72.7%, 80.1%) | 54.8% (50.4%, 59.2%) |

| | | | | | | | | | |
|---|---|---|---|---|---|---|---|---|---|
| | | Yes | Low | 2,138 (1,210, 3,295) | 57.1% (-5.6%, 94.3%) | -2.9 (-4.2, -1.6) | 241.4 (201.4, 281.4) | 94.6% (92.6%, 96.6%) | 22.6% (18.9%, 26.3%) |
| | | | Medium | 819 (642, 1,113) | 56.6% (-8.4%, 87.4%) | -3.4 (-4.7, -2.2) | 214.8 (176.5, 253) | 96.2% (94.5%, 97.9%) | 22.2% (18.6%, 25.8%) |
| | | | High | 573 (451, 747) | 54.1% (-19.4%, 84.9%) | -5.9 (-7.2, -4.6) | 267.3 (214.6, 319.9) | 94.8% (92.9%, 96.7%) | 24.6% (20.8%, 28.4%) |
| | Inferior | No | Low | 2,032 (1,327, 3,296) | 55.6% (-13.1%, 91.6%) | -4.4 (-5.7, -3) | 263.7 (216.8, 310.7) | 93.4% (91.2%, 95.6%) | 25% (21.2%, 28.8%) |
| | | | Medium | 694 (530, 924) | 52.1% (-11.5%, 89%) | -7.9 (-9.4, -6.5) | 329.2 (272.9, 385.6) | 92.4% (90.1%, 94.7%) | 30.4% (26.4%, 34.4%) |
| | | | High | 446 (353, 570) | 49.1% (-14.2%, 84.3%) | -10.9 (-12.3, -9.5) | 384.7 (330.9, 438.5) | 88.8% (86%, 91.6%) | 39.2% (34.9%, 43.5%) |
| | | Yes | Low | 2,033 (1,327, 3,581) | 57.4% (-3%, 87.5%) | -2.6 (-4, -1.2) | 245.8 (205, 286.5) | 93.4% (91.2%, 95.6%) | 21.6% (18%, 25.2%) |
| | | | Medium | 699 (523, 946) | 56.2% (-15.3%, 86%) | -3.8 (-5.2, -2.4) | 271.9 (221.1, 322.7) | 94.4% (92.4%, 96.4%) | 22% (18.4%, 25.6%) |
| | | | High | 455 (353, 591) | 55.1% (-5.2%, 87.1%) | -4.9 (-6.2, -3.6) | 237.2 (199.9, 274.6) | 95% (93.1%, 96.9%) | 26% (22.2%, 29.8%) |

**Supplemental Table 14. Mean estimate, bias, mean squared error (MSE), coverage, and false negative rate for vaccine efficacy (VE) estimates against severe *Shigella* diarrhea from realistically sized trials for all 24 simulation scenarios when using active surveillance for infection and a crude recurrent outcome regression model.** Hybrid immunity was either not possible (no) or possible such that the first post-vaccination infection conferred additional immunity (yes). Bias was calculated as the average VE estimate for a set of simulations minus the true VE (60%). MSE was calculated as the average of the square of the estimate minus the true parameter. Coverage represents the percent of 95% confidence intervals (CIs) that contained the true VE (60%), and the false negative rate indicates the percent of 95% CIs that incorrectly indicated a non-significant effect.

| VE against infection | Strength of infection-acquired immunity | Hybrid immunity | Force of infection | Mean N (range) | Mean VE estimate (range) | Bias (95% CI) | MSE (95% CI) | Coverage (95% CI) | False negatives (95% CI) |
|---|---|---|---|---|---|---|---|---|---|
| 0% | Similar | No | Low | 2,103 (1,419, 3,580) | 53.5% (-8.3%, 88.2%) | -6.5 (-8, -5.1) | 313.2 (260, 366.4) | 93% (90.8%, 95.2%) | 29.4% (25.4%, 33.4%) |
| | | | Medium | 767 (600, 1,043) | 44.8% (-21.5%, 80.6%) | -15.2 (-16.7, -13.6) | 550.5 (473.5, 627.5) | 83% (79.7%, 86.3%) | 45.2% (40.8%, 49.6%) |
| | | | High | 518 (403, 653) | 37.2% (-56.9%, 80%) | -22.8 (-24.7, -20.9) | 992.6 (836.4, 1148.9) | 71.2% (67.2%, 75.2%) | 58.8% (54.5%, 63.1%) |
| | | Yes | Low | 2,098 (1,395, 3,582) | 57.6% (-20%, 92.3%) | -2.4 (-3.7, -1.1) | 227.2 (185.5, 268.9) | 96.6% (95%, 98.2%) | 21.8% (18.2%, 25.4%) |
| | | | Medium | 808 (618, 1,055) | 56.7% (4.6%, 95.1%) | -3.3 (-4.6, -2) | 225.1 (191.7, 258.6) | 93.6% (91.5%, 95.7%) | 22.8% (19.1%, 26.5%) |
| | | | High | 573 (419, 716) | 54% (-25%, 89.3%) | -6 (-7.5, -4.5) | 325.5 (262.9, 388) | 91.6% (89.2%, 94%) | 27.6% (23.7%, 31.5%) |
| | Inferior | No | Low | 1,984 (1,328, 3,295) | 56.4% (-26.3%, 89.2%) | -3.6 (-5, -2.2) | 274.7 (224.7, 324.8) | 94% (91.9%, 96.1%) | 26.8% (22.9%, 30.7%) |
| | | | Medium | 682 (545, 895) | 51.5% (-12.6%, 86.2%) | -8.5 (-9.9, -7.1) | 334.3 (279.6, 388.9) | 92.2% (89.8%, 94.6%) | 32.8% (28.7%, 36.9%) |
| | | | High | 444 (368, 548) | 46.9% (-12.5%, 80.6%) | -13.1 (-14.6, -11.5) | 468.5 (403.2, 533.8) | 86.6% (83.6%, 89.6%) | 42.4% (38.1%, 46.7%) |
| | | Yes | Low | 1,991 (1,351, 3,295) | 58.4% (-21.4%, 90.6%) | -1.6 (-2.9, -0.2) | 242.3 (198.5, 286.1) | 94.6% (92.6%, 96.6%) | 21.6% (18%, 25.2%) |
| | | | Medium | 686 (555, 980) | 55.4% (-25.1%, 89.7%) | -4.6 (-6, -3.2) | 268.1 (218.4, 317.9) | 94.2% (92.2%, 96.2%) | 25.4% (21.6%, 29.2%) |
| | | | High | 451 (366, 552) | 54.4% (-15.9%, 85.7%) | -5.6 (-7, -4.3) | 273.5 (224.9, 322.2) | 93.6% (91.5%, 95.7%) | 27% (23.1%, 30.9%) |
| 20% | Similar | No | Low | 2,087 (1,287, 3,582) | 53.3% (-7.1%, 90%) | -6.7 (-8, -5.3) | 294.7 (244.4, 344.9) | 93.4% (91.2%, 95.6%) | 28% (24.1%, 31.9%) |
| | | | Medium | 772 (583, 1,083) | 44.6% (-26.1%, 86.9%) | -15.4 (-17, -13.8) | 552.9 (472.8, 632.9) | 82.2% (78.8%, 85.6%) | 47.2% (42.8%, 51.6%) |
| | | | High | 516 (407, 642) | 39.1% (-60.1%, 80%) | -20.9 (-22.7, -19.2) | 831.1 (702.3, 959.8) | 75.6% (71.8%, 79.4%) | 57.2% (52.9%, 61.5%) |

| | | | | | | | | | |
|---|---|---|---|---|---|---|---|---|---|
| | | Yes | Low | 2,093 (1,210, 3,169) | 56.5% (-10%, 94.1%) | -3.5 (-4.9, -2.2) | 247 (207.2, 286.8) | 95% (93.1%, 96.9%) | 24.8% (21%, 28.6%) |
| | | | Medium | 804 (632, 1,113) | 55.1% (-5.1%, 86.1%) | -4.9 (-6.2, -3.6) | 238.9 (195.4, 282.3) | 95.2% (93.3%, 97.1%) | 24.8% (21%, 28.6%) |
| | | | High | 561 (442, 710) | 52.2% (-19.7%, 83.4%) | -7.8 (-9.2, -6.4) | 303.2 (245.5, 361) | 94% (91.9%, 96.1%) | 31.6% (27.5%, 35.7%) |
| | Inferior | No | Low | 1,996 (1,307, 3,296) | 55.5% (-15.3%, 91.7%) | -4.5 (-5.9, -3.2) | 267.3 (220.1, 314.5) | 94% (91.9%, 96.1%) | 26.4% (22.5%, 30.3%) |
| | | | Medium | 683 (527, 895) | 51.5% (-17.5%, 88.6%) | -8.5 (-9.9, -7) | 346.1 (286, 406.2) | 91.6% (89.2%, 94%) | 32.8% (28.7%, 36.9%) |
| | | | High | 438 (350, 563) | 48.1% (-6.3%, 82.4%) | -11.9 (-13.4, -10.5) | 416 (358.6, 473.4) | 88% (85.2%, 90.8%) | 41.6% (37.3%, 45.9%) |
| | | Yes | Low | 1,994 (1,327, 3,433) | 57.2% (-4.7%, 88%) | -2.8 (-4.2, -1.5) | 245.2 (205.3, 285.2) | 93.8% (91.7%, 95.9%) | 22% (18.4%, 25.6%) |
| | | | Medium | 688 (527, 946) | 55.4% (-15.7%, 85.7%) | -4.6 (-6, -3.1) | 294.4 (238, 350.8) | 93.6% (91.5%, 95.7%) | 23.8% (20.1%, 27.5%) |
| | | | High | 448 (345, 587) | 53.9% (-18%, 86.7%) | -6.1 (-7.4, -4.8) | 265.2 (220.2, 310.2) | 94% (91.9%, 96.1%) | 26.2% (22.3%, 30.1%) |

**Supplemental Table 15. Mean estimate, bias, mean squared error (MSE), coverage, and false negative rate for vaccine efficacy (VE) estimates against severe *Shigella* diarrhea from realistically sized trials for all 24 simulation scenarios when using symptom-based reporting and a crude recurrent outcome regression model.** Hybrid immunity was either not possible (no) or possible such that the first post-vaccination infection conferred additional immunity (yes). Bias was calculated as the average VE estimate for a set of simulations minus the true VE (60%). MSE was calculated as the average of the square of the estimate minus the true parameter. Coverage represents the percent of 95% confidence intervals (CIs) that contained the true VE (60%), and the false negative rate indicates the percent of 95% CIs that incorrectly indicated a non-significant effect.

| VE against infection | Strength of infection-acquired immunity | Hybrid immunity | Force of infection | Mean N (range) | Mean VE estimate (range) | Bias (95% CI) | MSE (95% CI) | Coverage (95% CI) | False negatives (95% CI) |
|---|---|---|---|---|---|---|---|---|---|
| 0% | Similar | No | Low | 2,151 (1,419, 3,744) | 53.3% (-7.7%, 88.2%) | -6.7 (-8.1, -5.2) | 308.4 (257, 359.9) | 92.6% (90.3%, 94.9%) | 28.8% (24.8%, 32.8%) |
| | | | Medium | 780 (613, 1,056) | 44.8% (-21.4%, 80.7%) | -15.2 (-16.8, -13.7) | 550.6 (473.7, 627.5) | 83.2% (79.9%, 86.5%) | 44.4% (40%, 48.8%) |
| | | | High | 528 (408, 674) | 37% (-55.5%, 78.4%) | -23 (-24.9, -21.2) | 991 (841, 1141) | 70.8% (66.8%, 74.8%) | 58.2% (53.9%, 62.5%) |
| | | Yes | Low | 2,141 (1,395, 3,582) | 57.3% (-21%, 90%) | -2.7 (-4, -1.4) | 229.9 (183.6, 276.2) | 96.4% (94.8%, 98%) | 21.4% (17.8%, 25%) |
| | | | Medium | 823 (622, 1,097) | 56.9% (4.4%, 95.2%) | -3.1 (-4.4, -1.9) | 220.3 (186.8, 253.9) | 93.8% (91.7%, 95.9%) | 22% (18.4%, 25.6%) |
| | | | High | 587 (421, 741) | 54.2% (-24.9%, 89.3%) | -5.8 (-7.3, -4.4) | 307.6 (249.3, 365.9) | 91.6% (89.2%, 94%) | 27.2% (23.3%, 31.1%) |
| | Inferior | No | Low | 2,021 (1,371, 3,295) | 56.4% (-26.3%, 89.2%) | -3.6 (-5, -2.2) | 269.5 (220, 319) | 94.8% (92.9%, 96.7%) | 24.6% (20.8%, 28.4%) |
| | | | Medium | 692 (559, 935) | 51.6% (-11.7%, 86.2%) | -8.4 (-9.8, -7) | 331.8 (276.2, 387.4) | 92.4% (90.1%, 94.7%) | 31.4% (27.3%, 35.5%) |
| | | | High | 452 (370, 551) | 47.1% (-5.9%, 81.1%) | -12.9 (-14.4, -11.4) | 455.7 (392.3, 519.2) | 86.4% (83.4%, 89.4%) | 41.4% (37.1%, 45.7%) |
| | | Yes | Low | 2,028 (1,419, 3,295) | 58.4% (-13.3%, 90.6%) | -1.6 (-2.9, -0.2) | 236.8 (198, 275.7) | 94.8% (92.9%, 96.7%) | 20.8% (17.2%, 24.4%) |
| | | | Medium | 697 (559, 1,003) | 55.6% (-22.2%, 89.8%) | -4.4 (-5.8, -3) | 264.8 (214.4, 315.2) | 93.8% (91.7%, 95.9%) | 24.6% (20.8%, 28.4%) |
| | | | High | 459 (371, 563) | 54.5% (-15.7%, 87.2%) | -5.5 (-6.9, -4.2) | 264.4 (217.6, 311.2) | 93.6% (91.5%, 95.7%) | 27.6% (23.7%, 31.5%) |
| 20% | Similar | No | Low | 2,132 (1,349, 3,582) | 53.3% (-7.1%, 90%) | -6.7 (-8, -5.3) | 290.2 (241.2, 339.3) | 93.8% (91.7%, 95.9%) | 27% (23.1%, 30.9%) |
| | | | Medium | 784 (591, 1,069) | 44.6% (-26.1%, 87.2%) | -15.4 (-16.9, -13.8) | 541.9 (464.7, 619.1) | 82.6% (79.3%, 85.9%) | 46.8% (42.4%, 51.2%) |
| | | | High | 526 (411, 668) | 38.8% (-73.4%, 80.6%) | -21.2 (-22.9, -19.4) | 843.9 (706.6, 981.2) | 75% (71.2%, 78.8%) | 56.2% (51.9%, 60.5%) |

| | | | | | | | | | |
|---|---|---|---|---|---|---|---|---|---|
| | | Yes | Low | 2,138 (1,210, 3,295) | 56.5% (-6.2%, 94.1%) | -3.5 (-4.8, -2.1) | 250.8 (209.2, 292.5) | 94.8% (92.9%, 96.7%) | 23.2% (19.5%, 26.9%) |
| | | | Medium | 819 (642, 1,113) | 55.2% (-10.5%, 86.9%) | -4.8 (-6.1, -3.6) | 235.7 (193.3, 278.1) | 95.2% (93.3%, 97.1%) | 24% (20.3%, 27.7%) |
| | | | High | 573 (451, 747) | 52.2% (-25%, 84.1%) | -7.8 (-9.1, -6.4) | 305.3 (244.5, 366.1) | 93.8% (91.7%, 95.9%) | 30.6% (26.6%, 34.6%) |
| | Inferior | No | Low | 2,032 (1,327, 3,296) | 55.5% (-15.4%, 91.7%) | -4.5 (-5.9, -3.2) | 263.5 (216.2, 310.8) | 93.4% (91.2%, 95.6%) | 25% (21.2%, 28.8%) |
| | | | Medium | 694 (530, 924) | 51.5% (-17.6%, 88.9%) | -8.5 (-9.9, -7.1) | 341.6 (283.1, 400.1) | 91% (88.5%, 93.5%) | 31.6% (27.5%, 35.7%) |
| | | | High | 446 (353, 570) | 48.2% (-6.4%, 83.3%) | -11.8 (-13.3, -10.4) | 403.9 (349, 458.8) | 88.2% (85.4%, 91%) | 40% (35.7%, 44.3%) |
| | | Yes | Low | 2,033 (1,327, 3,581) | 57.2% (-4.8%, 87.5%) | -2.8 (-4.2, -1.5) | 246.3 (205.8, 286.8) | 92.8% (90.5%, 95.1%) | 21.8% (18.2%, 25.4%) |
| | | | Medium | 699 (523, 946) | 55.5% (-15.7%, 85.7%) | -4.5 (-6, -3.1) | 287.3 (232.7, 342) | 93.4% (91.2%, 95.6%) | 23% (19.3%, 26.7%) |
| | | | High | 455 (353, 591) | 53.9% (-8.3%, 86.7%) | -6.1 (-7.4, -4.8) | 256.8 (216.3, 297.3) | 94% (91.9%, 96.1%) | 27% (23.1%, 30.9%) |

**Supplemental Table 16. Mean estimate, bias, mean squared error (MSE), coverage, and false negative rate for vaccine efficacy (VE) estimates against any *Shigella* diarrhea from highly powered trials for all 24 simulation scenarios when using active surveillance for infection and a single outcome regression model.** Infection-acquired immunity was either inferior or similar to vaccine-acquired immunity. Hybrid immunity was either not possible (no) or possible such that the first post-vaccination infection conferred additional immunity (yes). Bias was calculated as the average VE estimate for a set of simulations minus the true VE (60%). MSE was calculated as the average of the square of the estimate minus the true parameter. Coverage represents the percent of 95% confidence intervals (CIs) that contained the true VE (60%), and the false negative rate indicates the percent of 95% CIs that incorrectly indicated a non-significant effect.

| VE against infection | Strength of infection-acquired immunity | Hybrid immunity | Force of infection | Mean VE estimate (range) | Bias (95% CI) | MSE (95% CI) | Coverage (95% CI) | False negatives (95% CI) |
|---|---|---|---|---|---|---|---|---|
| 0% | Similar | No | Low | 39.6% (23.3%, 49.1%) | -0.4 (-0.7, -0.1) | 14.4 (12.3, 16.4) | 95.4% (93.6%, 97.2%) | 0% (0%, 0%) |
| | | | Medium | 39.9% (28.2%, 46.9%) | -0.1 (-0.3, 0.2) | 6.9 (5.9, 7.9) | 95.6% (93.8%, 97.4%) | 0% (0%, 0%) |
| | | | High | 40% (33.6%, 47.4%) | 0 (-0.2, 0.3) | 6 (5.3, 6.7) | 95.4% (93.6%, 97.2%) | 0% (0%, 0%) |
| | | Yes | Low | 39.6% (27.3%, 48.8%) | -0.4 (-0.7, -0.1) | 13.8 (12, 15.6) | 96.4% (94.8%, 98%) | 0% (0%, 0%) |
| | | | Medium | 39.9% (30.3%, 47.1%) | -0.1 (-0.3, 0.2) | 7.5 (6.5, 8.4) | 95% (93.1%, 96.9%) | 0% (0%, 0%) |
| | | | High | 40% (33.3%, 46.1%) | 0 (-0.3, 0.2) | 5.6 (4.9, 6.3) | 95% (93.1%, 96.9%) | 0% (0%, 0%) |
| | Inferior | No | Low | 39.8% (26%, 51.4%) | -0.2 (-0.6, 0.1) | 15.3 (13.3, 17.3) | 94.6% (92.6%, 96.6%) | 0% (0%, 0%) |
| | | | Medium | 39.8% (30.6%, 49.3%) | -0.2 (-0.5, 0) | 7 (6.1, 7.9) | 95.8% (94%, 97.6%) | 0% (0%, 0%) |
| | | | High | 39.9% (33.1%, 45.9%) | -0.1 (-0.3, 0.1) | 5.9 (5.2, 6.6) | 94.8% (92.9%, 96.7%) | 0% (0%, 0%) |
| | | Yes | Low | 39.8% (28.5%, 51.3%) | -0.2 (-0.6, 0.1) | 15.1 (13.2, 17) | 95% (93.1%, 96.9%) | 0% (0%, 0%) |
| | | | Medium | 39.8% (30.1%, 47.4%) | -0.2 (-0.4, 0.1) | 6.8 (5.9, 7.7) | 95.8% (94%, 97.6%) | 0% (0%, 0%) |
| | | | High | 39.9% (33%, 46.8%) | -0.1 (-0.3, 0.1) | 5.9 (5.2, 6.6) | 95.2% (93.3%, 97.1%) | 0% (0%, 0%) |
| 20% | Similar | No | Low | 39.8% (26.9%, 52.1%) | -0.2 (-0.6, 0.1) | 15.4 (13.5, 17.3) | 94.8% (92.9%, 96.7%) | 0% (0%, 0%) |
| | | | Medium | 39.9% (31.2%, 47.4%) | -0.1 (-0.4, 0.1) | 6.9 (6.1, 7.8) | 94.4% (92.4%, 96.4%) | 0% (0%, 0%) |
| | | | High | 39.9% (31.8%, 45.9%) | -0.1 (-0.3, 0.1) | 4.9 (4.3, 5.6) | 95.4% (93.6%, 97.2%) | 0% (0%, 0%) |

| | | | | | | | | |
|---|---|---|---|---|---|---|---|---|
| | | Yes | Low | 40% (28.4%, 50.1%) | 0 (-0.3, 0.3) | 13.8 (12.2, 15.5) | 94.8% (92.9%, 96.7%) | 0% (0%, 0%) |
| | | | Medium | 39.9% (31.8%, 46.2%) | -0.1 (-0.4, 0.1) | 6.2 (5.5, 6.9) | 96.6% (95%, 98.2%) | 0% (0%, 0%) |
| | | | High | 39.8% (33.9%, 46.7%) | -0.2 (-0.4, 0) | 5.3 (4.7, 6) | 94.8% (92.9%, 96.7%) | 0% (0%, 0%) |
| | Inferior | No | Low | 39.7% (28.1%, 50.4%) | -0.3 (-0.6, 0.1) | 14.5 (12.8, 16.1) | 95.8% (94%, 97.6%) | 0% (0%, 0%) |
| | | | Medium | 40% (30.8%, 46.5%) | 0 (-0.3, 0.2) | 6.6 (5.8, 7.5) | 95.6% (93.8%, 97.4%) | 0% (0%, 0%) |
| | | | High | 39.8% (30.8%, 47.2%) | -0.2 (-0.4, 0.1) | 5.9 (5.1, 6.7) | 94% (91.9%, 96.1%) | 0% (0%, 0%) |
| | | Yes | Low | 39.8% (27.2%, 51.3%) | -0.2 (-0.6, 0.1) | 14.5 (12.8, 16.3) | 95.4% (93.6%, 97.2%) | 0% (0%, 0%) |
| | | | Medium | 40% (31.9%, 48.2%) | 0 (-0.2, 0.2) | 6.7 (5.8, 7.5) | 94.2% (92.2%, 96.2%) | 0% (0%, 0%) |
| | | | High | 39.9% (31.3%, 47.4%) | -0.1 (-0.4, 0.1) | 5.5 (4.7, 6.3) | 94.4% (92.4%, 96.4%) | 0% (0%, 0%) |

**Supplemental Table 17. Mean estimate, bias, mean squared error (MSE), coverage, and false negative rate for vaccine efficacy (VE) estimates against any *Shigella* diarrhea from highly powered trials for all 24 simulation scenarios when using symptom-based reporting and a single outcome regression model. Infection-acquired immunity was either inferior or similar to vaccine-acquired immunity.** Hybrid immunity was either not possible (no) or possible such that the first post-vaccination infection conferred additional immunity (yes). Bias was calculated as the average VE estimate for a set of simulations minus the true VE (60%). MSE was calculated as the average of the square of the estimate minus the true parameter. Coverage represents the percent of 95% confidence intervals (CIs) that contained the true VE (60%), and the false negative rate indicates the percent of 95% CIs that incorrectly indicated a non-significant effect.

| VE against infection | Strength of infection-acquired immunity | Hybrid immunity | Force of infection | Mean VE estimate (range) | Bias (95% CI) | MSE (95% CI) | Coverage (95% CI) | False negatives (95% CI) |
|---|---|---|---|---|---|---|---|---|
| 0% | Similar | No | Low | 36.3% (20.8%, 48.3%) | -3.7 (-4, -3.4) | 27 (23.6, 30.4) | 85.2% (82.1%, 88.3%) | 0% (0%, 0%) |
| | | | Medium | 30.7% (21.8%, 38%) | -9.3 (-9.5, -9.1) | 93.1 (88.8, 97.4) | 2.8% (1.4%, 4.2%) | 0% (0%, 0%) |
| | | | High | 26.5% (19.9%, 32.9%) | -13.5 (-13.7, -13.3) | 188.2 (182.6, 193.9) | 0% (0%, 0%) | 0% (0%, 0%) |
| | | Yes | Low | 40% (27.6%, 49.4%) | 0 (-0.3, 0.3) | 12.4 (10.7, 14) | 96% (94.3%, 97.7%) | 0% (0%, 0%) |
| | | | Medium | 39.5% (31%, 46.5%) | -0.5 (-0.7, -0.3) | 5.8 (5, 6.6) | 94.2% (92.2%, 96.2%) | 0% (0%, 0%) |
| | | | High | 37.3% (30.6%, 43.6%) | -2.7 (-2.9, -2.5) | 11.5 (10.4, 12.6) | 71.8% (67.9%, 75.7%) | 0% (0%, 0%) |
| | Inferior | No | Low | 39% (29.1%, 49.2%) | -1 (-1.4, -0.7) | 14.1 (12.3, 15.8) | 93.8% (91.7%, 95.9%) | 0% (0%, 0%) |
| | | | Medium | 37.2% (30.3%, 44%) | -2.8 (-3, -2.6) | 13.4 (12.1, 14.7) | 72.6% (68.7%, 76.5%) | 0% (0%, 0%) |
| | | | High | 35.6% (30.2%, 41.3%) | -4.4 (-4.5, -4.2) | 22.3 (20.9, 23.8) | 34.6% (30.4%, 38.8%) | 0% (0%, 0%) |
| | | Yes | Low | 39.9% (29%, 50.7%) | -0.1 (-0.4, 0.2) | 12.5 (11, 14.1) | 95.6% (93.8%, 97.4%) | 0% (0%, 0%) |
| | | | Medium | 39.6% (32%, 45.1%) | -0.4 (-0.6, -0.2) | 4.9 (4.3, 5.5) | 95.2% (93.3%, 97.1%) | 0% (0%, 0%) |
| | | | High | 38.7% (33.7%, 44.1%) | -1.3 (-1.4, -1.1) | 4.9 (4.4, 5.4) | 88.8% (86%, 91.6%) | 0% (0%, 0%) |
| 20% | Similar | No | Low | 36.5% (23.6%, 47.6%) | -3.5 (-3.8, -3.2) | 27.6 (24.5, 30.6) | 84.4% (81.2%, 87.6%) | 0% (0%, 0%) |
| | | | Medium | 30.7% (22.5%, 37.2%) | -9.3 (-9.5, -9.1) | 92.6 (88.3, 96.9) | 2.8% (1.4%, 4.2%) | 0% (0%, 0%) |
| | | | High | 26.4% (20.8%, 32.6%) | -13.6 (-13.8, -13.4) | 189.7 (184.7, 194.8) | 0% (0%, 0%) | 0% (0%, 0%) |

| | | | | | | | | |
|---|---|---|---|---|---|---|---|---|
| | | Yes | Low | 39.5% (27.7%, 49%) | -0.5 (-0.8, -0.2) | 13.8 (12, 15.5) | 94.4% (92.4%, 96.4%) | 0% (0%, 0%) |
| | | | Medium | 37.7% (30.3%, 43.5%) | -2.3 (-2.5, -2.1) | 10.4 (9.3, 11.6) | 84.4% (81.2%, 87.6%) | 0% (0%, 0%) |
| | | | High | 35.3% (28.4%, 41.8%) | -4.7 (-4.9, -4.5) | 26.6 (24.7, 28.5) | 36.8% (32.6%, 41%) | 0% (0%, 0%) |
| | Inferior | No | Low | 39% (26.9%, 48.6%) | -1 (-1.3, -0.7) | 13.8 (12.2, 15.5) | 94.8% (92.9%, 96.7%) | 0% (0%, 0%) |
| | | | Medium | 37.3% (28.2%, 43.8%) | -2.7 (-2.9, -2.5) | 12.4 (11, 13.8) | 76.2% (72.5%, 79.9%) | 0% (0%, 0%) |
| | | | High | 35.7% (29.2%, 42.9%) | -4.3 (-4.5, -4.1) | 22.9 (21.3, 24.6) | 36.2% (32%, 40.4%) | 0% (0%, 0%) |
| | | Yes | Low | 39.8% (28%, 49.7%) | -0.2 (-0.5, 0.1) | 12.7 (11.2, 14.1) | 94.4% (92.4%, 96.4%) | 0% (0%, 0%) |
| | | | Medium | 39.2% (32.1%, 45%) | -0.8 (-1, -0.6) | 5.4 (4.7, 6.1) | 94.4% (92.4%, 96.4%) | 0% (0%, 0%) |
| | | | High | 38.2% (31.8%, 44.7%) | -1.8 (-2, -1.6) | 7.1 (6.2, 7.9) | 80% (76.5%, 83.5%) | 0% (0%, 0%) |

**Supplemental Table 18. Mean estimate, bias, mean squared error (MSE), coverage, and false negative rate for vaccine efficacy (VE) estimates against any *Shigella* diarrhea from highly powered trials for all 24 simulation scenarios when using active surveillance for infection and a stratified recurrent outcome regression model.** Hybrid immunity was either not possible (no) or possible such that the first post-vaccination infection conferred additional immunity (yes). Bias was calculated as the average VE estimate for a set of simulations minus the true VE (60%). MSE was calculated as the average of the square of the estimate minus the true parameter. Coverage represents the percent of 95% confidence intervals (CIs) that contained the true VE (60%), and the false negative rate indicates the percent of 95% CIs that incorrectly indicated a non-significant effect.

| VE against infection | Strength of infection-acquired immunity | Hybrid immunity | Force of infection | Mean VE estimate (range) | Bias (95% CI) | MSE (95% CI) | Coverage (95% CI) | False negatives (95% CI) |
|---|---|---|---|---|---|---|---|---|
| 0% | Similar | No | Low | 34.6% (17%, 47.2%) | -5.4 (-5.7, -5) | 42.6 (38.2, 47.1) | 71.6% (67.6%, 75.6%) | 0% (0%, 0%) |
| 0% | Similar | No | Medium | 26.3% (16.8%, 33.2%) | -13.7 (-13.9, -13.5) | 193 (187, 198.9) | 0% (0%, 0%) | 0% (0%, 0%) |
| 0% | Similar | No | High | 20.5% (14.3%, 26.5%) | -19.5 (-19.7, -19.3) | 383.8 (376.1, 391.4) | 0% (0%, 0%) | 0% (0%, 0%) |
| 0% | Similar | Yes | Low | 39.2% (27.2%, 48.9%) | -0.8 (-1.1, -0.5) | 13.1 (11.3, 14.8) | 96.4% (94.8%, 98%) | 0% (0%, 0%) |
| 0% | Similar | Yes | Medium | 36.7% (28.9%, 44.3%) | -3.3 (-3.5, -3.1) | 16.2 (14.6, 17.7) | 68.6% (64.5%, 72.7%) | 0% (0%, 0%) |
| 0% | Similar | Yes | High | 32.9% (26.5%, 38.5%) | -7.1 (-7.3, -6.9) | 53.9 (51.5, 56.4) | 3.6% (2%, 5.2%) | 0% (0%, 0%) |
| 0% | Inferior | No | Low | 38.5% (26.9%, 47.8%) | -1.5 (-1.9, -1.2) | 15.1 (13.2, 16.9) | 93.2% (91%, 95.4%) | 0% (0%, 0%) |
| 0% | Inferior | No | Medium | 35.4% (28.2%, 41.3%) | -4.6 (-4.7, -4.4) | 25.1 (23.3, 26.9) | 39.8% (35.5%, 44.1%) | 0% (0%, 0%) |
| 0% | Inferior | No | High | 32.6% (27%, 38.1%) | -7.4 (-7.6, -7.3) | 57.7 (55.5, 59.9) | 0.8% (0%, 1.6%) | 0% (0%, 0%) |
| 0% | Inferior | Yes | Low | 39.6% (27.3%, 50%) | -0.4 (-0.7, -0.1) | 12.3 (10.8, 13.9) | 95% (93.1%, 96.9%) | 0% (0%, 0%) |
| 0% | Inferior | Yes | Medium | 38.4% (31.3%, 43.6%) | -1.6 (-1.7, -1.4) | 6.3 (5.6, 7.1) | 90% (87.4%, 92.6%) | 0% (0%, 0%) |
| 0% | Inferior | Yes | High | 36.6% (32%, 41.5%) | -3.4 (-3.5, -3.2) | 14.1 (13.1, 15.1) | 44.8% (40.4%, 49.2%) | 0% (0%, 0%) |
| 20% | Similar | No | Low | 35.4% (23.5%, 46.1%) | -4.6 (-4.9, -4.2) | 36 (32.4, 39.6) | 74.8% (71%, 78.6%) | 0% (0%, 0%) |
| 20% | Similar | No | Medium | 27.6% (18.6%, 35.7%) | -12.4 (-12.6, -12.2) | 159.8 (154.1, 165.5) | 0% (0%, 0%) | 0% (0%, 0%) |
| 20% | Similar | No | High | 21.9% (16.4%, 27.3%) | -18.1 (-18.3, -18) | 333.3 (326.8, 339.9) | 0% (0%, 0%) | 0% (0%, 0%) |

| | | | | | | | | |
|---|---|---|---|---|---|---|---|---|
| | | Yes | Low | 39.7% (26.5%, 48.6%) | -0.3 (-0.6, 0) | 13.2 (11.5, 14.9) | 94.8% (92.9%, 96.7%) | 0% (0%, 0%) |
| | | | Medium | 37.7% (32.1%, 42.9%) | -2.3 (-2.5, -2.1) | 9.7 (8.7, 10.7) | 84.4% (81.2%, 87.6%) | 0% (0%, 0%) |
| | | | High | 34.8% (29.4%, 40.9%) | -5.2 (-5.4, -5) | 31.3 (29.4, 33.2) | 20.8% (17.2%, 24.4%) | 0% (0%, 0%) |
| | Inferior | No | Low | 38.7% (26.6%, 48.3%) | -1.3 (-1.6, -1) | 13.7 (12, 15.5) | 94.4% (92.4%, 96.4%) | 0% (0%, 0%) |
| | | | Medium | 36.3% (27.4%, 42.4%) | -3.7 (-3.9, -3.5) | 18.2 (16.5, 19.8) | 57% (52.7%, 61.3%) | 0% (0%, 0%) |
| | | | High | 33.8% (27.6%, 39.7%) | -6.2 (-6.3, -6) | 41.4 (39.3, 43.5) | 3.6% (2%, 5.2%) | 0% (0%, 0%) |
| | | Yes | Low | 39.8% (28.2%, 48%) | -0.2 (-0.5, 0.1) | 11.7 (10.4, 13.1) | 96.2% (94.5%, 97.9%) | 0% (0%, 0%) |
| | | | Medium | 39% (32.6%, 44.8%) | -1 (-1.2, -0.9) | 5.3 (4.6, 6) | 90.6% (88%, 93.2%) | 0% (0%, 0%) |
| | | | High | 37.2% (31.4%, 42.2%) | -2.8 (-2.9, -2.6) | 10.7 (9.7, 11.7) | 60.2% (55.9%, 64.5%) | 0% (0%, 0%) |

**Supplemental Table 19. Mean estimate, bias, mean squared error (MSE), coverage, and false negative rate for vaccine efficacy (VE) estimates against any *Shigella* diarrhea from highly powered trials for all 24 simulation scenarios when using symptom-based reporting and a stratified recurrent outcome regression model.** Hybrid immunity was either not possible (no) or possible such that the first post-vaccination infection conferred additional immunity (yes). Bias was calculated as the average VE estimate for a set of simulations minus the true VE (60%). MSE was calculated as the average of the square of the estimate minus the true parameter. Coverage represents the percent of 95% confidence intervals (CIs) that contained the true VE (60%), and the false negative rate indicates the percent of 95% CIs that incorrectly indicated a non-significant effect.

| VE against infection | Strength of infection-acquired immunity | Hybrid immunity | Force of infection | Mean VE estimate (range) | Bias (95% CI) | MSE (95% CI) | Coverage (95% CI) | False negatives (95% CI) |
|---|---|---|---|---|---|---|---|---|
| 0% | Similar | No | Low | 35.1% (17.8%, 48%) | -4.9 (-5.2, -4.6) | 38 (33.9, 42.2) | 75.6% (71.8%, 79.4%) | 0% (0%, 0%) |
| | | | Medium | 27.2% (17.6%, 34%) | -12.8 (-13.1, -12.6) | 170.9 (165.3, 176.5) | 0% (0%, 0%) | 0% (0%, 0%) |
| | | | High | 21.4% (15.1%, 27.2%) | -18.6 (-18.8, -18.5) | 352.8 (345.4, 360.2) | 0% (0%, 0%) | 0% (0%, 0%) |
| | | Yes | Low | 39.9% (28.2%, 49.3%) | -0.1 (-0.4, 0.2) | 12.3 (10.7, 13.9) | 96.2% (94.5%, 97.9%) | 0% (0%, 0%) |
| | | | Medium | 38.3% (30.2%, 45.7%) | -1.7 (-1.9, -1.5) | 8.3 (7.2, 9.3) | 87% (84.1%, 89.9%) | 0% (0%, 0%) |
| | | | High | 34.8% (28.2%, 40.4%) | -5.2 (-5.3, -5) | 30.4 (28.6, 32.2) | 21.2% (17.6%, 24.8%) | 0% (0%, 0%) |
| | Inferior | No | Low | 38.6% (27.5%, 48.3%) | -1.4 (-1.7, -1.1) | 14.7 (12.8, 16.5) | 93.6% (91.5%, 95.7%) | 0% (0%, 0%) |
| | | | Medium | 35.9% (28.5%, 41.9%) | -4.1 (-4.3, -4) | 21.5 (19.8, 23.1) | 48% (43.6%, 52.4%) | 0% (0%, 0%) |
| | | | High | 33.2% (27.3%, 38.7%) | -6.8 (-6.9, -6.6) | 48.9 (46.8, 51) | 1.4% (0.4%, 2.4%) | 0% (0%, 0%) |
| | | Yes | Low | 39.8% (27.9%, 50.4%) | -0.2 (-0.5, 0.1) | 12.2 (10.7, 13.7) | 95.2% (93.3%, 97.1%) | 0% (0%, 0%) |
| | | | Medium | 38.9% (31.8%, 44%) | -1.1 (-1.2, -0.9) | 5 (4.4, 5.7) | 93.6% (91.5%, 95.7%) | 0% (0%, 0%) |
| | | | High | 37.3% (32.4%, 42.1%) | -2.7 (-2.8, -2.5) | 9.8 (8.9, 10.6) | 63.8% (59.6%, 68%) | 0% (0%, 0%) |
| 20% | Similar | No | Low | 35.2% (23.2%, 45.8%) | -4.8 (-5.1, -4.4) | 37.7 (33.9, 41.5) | 73.6% (69.7%, 77.5%) | 0% (0%, 0%) |
| | | | Medium | 27.1% (18%, 35.6%) | -12.9 (-13.1, -12.7) | 172.5 (166.6, 178.5) | 0% (0%, 0%) | 0% (0%, 0%) |
| | | | High | 21.2% (15.6%, 26.6%) | -18.8 (-18.9, -18.6) | 356.2 (349.4, 363) | 0% (0%, 0%) | 0% (0%, 0%) |

| | | | | | | | | |
|---|---|---|---|---|---|---|---|---|
| | | Yes | Low | 39.4% (25.9%, 48.4%) | -0.6 (-0.9, -0.3) | 13.6 (11.8, 15.4) | 95% (93.1%, 96.9%) | 0% (0%, 0%) |
| | | | Medium | 36.7% (30.6%, 42.1%) | -3.3 (-3.4, -3.1) | 15.1 (13.7, 16.4) | 70% (66%, 74%) | 0% (0%, 0%) |
| | | | High | 33.4% (27.8%, 39.5%) | -6.6 (-6.8, -6.4) | 48.5 (46, 50.9) | 6.6% (4.4%, 8.8%) | 0% (0%, 0%) |
| | Inferior | No | Low | 38.7% (26.8%, 48.4%) | -1.3 (-1.6, -1) | 13.9 (12.2, 15.7) | 94% (91.9%, 96.1%) | 0% (0%, 0%) |
| | | | Medium | 36.2% (27.4%, 42.3%) | -3.8 (-4, -3.6) | 19.5 (17.7, 21.2) | 53.2% (48.8%, 57.6%) | 0% (0%, 0%) |
| | | | High | 33.6% (27.3%, 39.8%) | -6.4 (-6.5, -6.2) | 43.9 (41.7, 46) | 3.6% (2%, 5.2%) | 0% (0%, 0%) |
| | | Yes | Low | 39.7% (28.3%, 48.4%) | -0.3 (-0.6, 0) | 11.9 (10.5, 13.2) | 95.8% (94%, 97.6%) | 0% (0%, 0%) |
| | | | Medium | 38.8% (32.5%, 44.4%) | -1.2 (-1.4, -1.1) | 5.9 (5.2, 6.6) | 89.6% (86.9%, 92.3%) | 0% (0%, 0%) |
| | | | High | 37% (31.2%, 42.2%) | -3 (-3.1, -2.8) | 12 (11, 13.1) | 57% (52.7%, 61.3%) | 0% (0%, 0%) |

**Supplemental Table 20. Mean estimate, bias, mean squared error (MSE), coverage, and false negative rate for vaccine efficacy (VE) estimates against any *Shigella* diarrhea from highly powered trials for all 24 simulation scenarios when using active surveillance for infection and a crude recurrent outcome regression model.** Hybrid immunity was either not possible (no) or possible such that the first post-vaccination infection conferred additional immunity (yes). Bias was calculated as the average VE estimate for a set of simulations minus the true VE (60%). MSE was calculated as the average of the square of the estimate minus the true parameter. Coverage represents the percent of 95% confidence intervals (CIs) that contained the true VE (60%), and the false negative rate indicates the percent of 95% CIs that incorrectly indicated a non-significant effect.

| VE against infection | Strength of infection-acquired immunity | Hybrid immunity | Force of infection | Mean VE estimate (range) | Bias (95% CI) | MSE (95% CI) | Coverage (95% CI) | False negatives (95% CI) |
|---|---|---|---|---|---|---|---|---|
| 0% | Similar | No | Low | 34.6% (17.3%, 47.2%) | -5.4 (-5.7, -5) | 42.3 (37.9, 46.6) | 71.2% (67.2%, 75.2%) | 0% (0%, 0%) |
| | | | Medium | 26.3% (17.1%, 33%) | -13.7 (-13.9, -13.5) | 192.4 (186.6, 198.2) | 0% (0%, 0%) | 0% (0%, 0%) |
| | | | High | 20.5% (14.5%, 26.3%) | -19.5 (-19.7, -19.3) | 384.1 (376.6, 391.6) | 0% (0%, 0%) | 0% (0%, 0%) |
| | | Yes | Low | 39.2% (27.5%, 48.7%) | -0.8 (-1.1, -0.5) | 12.5 (10.9, 14.2) | 96.2% (94.5%, 97.9%) | 0% (0%, 0%) |
| | | | Medium | 36.7% (28.9%, 43.9%) | -3.3 (-3.5, -3.1) | 15.7 (14.2, 17.1) | 67.6% (63.5%, 71.7%) | 0% (0%, 0%) |
| | | | High | 32.9% (26.8%, 38.2%) | -7.1 (-7.2, -6.9) | 53.5 (51.2, 55.8) | 2.6% (1.2%, 4%) | 0% (0%, 0%) |
| | Inferior | No | Low | 38.5% (27%, 47.9%) | -1.5 (-1.9, -1.2) | 15 (13.1, 16.9) | 93.2% (91%, 95.4%) | 0% (0%, 0%) |
| | | | Medium | 35.4% (28.2%, 41.3%) | -4.6 (-4.7, -4.4) | 25 (23.2, 26.7) | 38.8% (34.5%, 43.1%) | 0% (0%, 0%) |
| | | | High | 32.6% (27%, 37.9%) | -7.4 (-7.6, -7.3) | 57.6 (55.4, 59.8) | 0.8% (0%, 1.6%) | 0% (0%, 0%) |
| | | Yes | Low | 39.6% (27.4%, 50%) | -0.4 (-0.7, -0.1) | 12.2 (10.7, 13.7) | 95% (93.1%, 96.9%) | 0% (0%, 0%) |
| | | | Medium | 38.4% (31.3%, 43.6%) | -1.6 (-1.7, -1.4) | 6.2 (5.4, 7) | 90.2% (87.6%, 92.8%) | 0% (0%, 0%) |
| | | | High | 36.6% (32.1%, 41.4%) | -3.4 (-3.5, -3.2) | 14 (13, 15) | 43% (38.7%, 47.3%) | 0% (0%, 0%) |
| 20% | Similar | No | Low | 34.8% (22.9%, 45.4%) | -5.2 (-5.5, -4.9) | 41.7 (37.7, 45.6) | 69.2% (65.2%, 73.2%) | 0% (0%, 0%) |
| | | | Medium | 26.3% (17.5%, 34.5%) | -13.7 (-13.9, -13.4) | 193 (186.8, 199.2) | 0% (0%, 0%) | 0% (0%, 0%) |
| | | | High | 20.4% (15%, 25.6%) | -19.6 (-19.7, -19.4) | 386.5 (379.6, 393.4) | 0% (0%, 0%) | 0% (0%, 0%) |

| | | | | | | | | |
|---|---|---|---|---|---|---|---|---|
| | | Yes | Low | 38.7%<br>(25.6%, 47.6%) | -1.3<br>(-1.6, -0.9) | 14.6<br>(12.7, 16.5) | 93%<br>(90.8%, 95.2%) | 0%<br>(0%, 0%) |
| | | | Medium | 35.2%<br>(28.8%, 40.1%) | -4.8<br>(-5, -4.6) | 27.1<br>(25.3, 28.9) | 38.8%<br>(34.5%, 43.1%) | 0%<br>(0%, 0%) |
| | | | High | 31.5%<br>(26.2%, 37.4%) | -8.5<br>(-8.7, -8.3) | 76.8<br>(73.7, 79.8) | 1%<br>(0.1%, 1.9%) | 0%<br>(0%, 0%) |
| | Inferior | No | Low | 38.5%<br>(26.8%, 48.2%) | -1.5<br>(-1.8, -1.2) | 14.2<br>(12.4, 16) | 93.8%<br>(91.7%, 95.9%) | 0%<br>(0%, 0%) |
| | | | Medium | 35.8%<br>(27.2%, 41.8%) | -4.2<br>(-4.3, -4) | 21.9<br>(20.1, 23.7) | 47.4%<br>(43%, 51.8%) | 0%<br>(0%, 0%) |
| | | | High | 33.2%<br>(27.2%, 39%) | -6.8<br>(-7, -6.6) | 49.6<br>(47.3, 51.8) | 1.6%<br>(0.5%, 2.7%) | 0%<br>(0%, 0%) |
| | | Yes | Low | 39.5%<br>(28.3%, 48.1%) | -0.5<br>(-0.8, -0.2) | 11.9<br>(10.5, 13.3) | 95.6%<br>(93.8%, 97.4%) | 0%<br>(0%, 0%) |
| | | | Medium | 38.3%<br>(32.2%, 44%) | -1.7<br>(-1.9, -1.5) | 7<br>(6.2, 7.9) | 85%<br>(81.9%, 88.1%) | 0%<br>(0%, 0%) |
| | | | High | 36.4%<br>(30.7%, 41.2%) | -3.6<br>(-3.8, -3.5) | 16.1<br>(14.9, 17.2) | 39%<br>(34.7%, 43.3%) | 0%<br>(0%, 0%) |

**Supplemental Table 21. Mean estimate, bias, mean squared error (MSE), coverage, and false negative rate for vaccine efficacy (VE) estimates against any *Shigella* diarrhea from highly powered trials for all 24 simulation scenarios when using symptom-based reporting and a crude recurrent outcome regression model.** Hybrid immunity was either not possible (no) or possible such that the first post-vaccination infection conferred additional immunity (yes). Bias was calculated as the average VE estimate for a set of simulations minus the true VE (60%). MSE was calculated as the average of the square of the estimate minus the true parameter. Coverage represents the percent of 95% confidence intervals (CIs) that contained the true VE (60%), and the false negative rate indicates the percent of 95% CIs that incorrectly indicated a non-significant effect.

| VE against infection | Strength of infection-acquired immunity | Hybrid immunity | Force of infection | Mean VE estimate (range) | Bias (95% CI) | MSE (95% CI) | Coverage (95% CI) | False negatives (95% CI) |
|---|---|---|---|---|---|---|---|---|
| 0% | Similar | No | Low | 34.7% (17.3%, 47.2%) | -5.3 (-5.7, -5) | 42.2 (37.8, 46.5) | 71.2% (67.2%, 75.2%) | 0% (0%, 0%) |
| 0% | Similar | No | Medium | 26.4% (17.1%, 33%) | -13.6 (-13.9, -13.4) | 191.8 (186, 197.6) | 0% (0%, 0%) | 0% (0%, 0%) |
| 0% | Similar | No | High | 20.5% (14.5%, 26.4%) | -19.5 (-19.6, -19.3) | 383.1 (375.6, 390.5) | 0% (0%, 0%) | 0% (0%, 0%) |
| 0% | Similar | Yes | Low | 39.2% (27.5%, 48.7%) | -0.8 (-1.1, -0.4) | 12.5 (10.9, 14.2) | 96.2% (94.5%, 97.9%) | 0% (0%, 0%) |
| 0% | Similar | Yes | Medium | 36.8% (28.9%, 43.9%) | -3.2 (-3.4, -3) | 15.5 (14, 17) | 67.6% (63.5%, 71.7%) | 0% (0%, 0%) |
| 0% | Similar | Yes | High | 33% (26.9%, 38.2%) | -7 (-7.2, -6.9) | 53 (50.7, 55.3) | 2.8% (1.4%, 4.2%) | 0% (0%, 0%) |
| 0% | Inferior | No | Low | 38.5% (27%, 47.9%) | -1.5 (-1.9, -1.2) | 15 (13.1, 16.8) | 93.4% (91.2%, 95.6%) | 0% (0%, 0%) |
| 0% | Inferior | No | Medium | 35.5% (28.3%, 41.3%) | -4.5 (-4.7, -4.3) | 24.7 (23, 26.5) | 39.4% (35.1%, 43.7%) | 0% (0%, 0%) |
| 0% | Inferior | No | High | 32.6% (27.1%, 38%) | -7.4 (-7.5, -7.2) | 57 (54.8, 59.2) | 0.8% (0%, 1.6%) | 0% (0%, 0%) |
| 0% | Inferior | Yes | Low | 39.6% (27.4%, 50%) | -0.4 (-0.7, -0.1) | 12.2 (10.7, 13.7) | 95.2% (93.3%, 97.1%) | 0% (0%, 0%) |
| 0% | Inferior | Yes | Medium | 38.5% (31.4%, 43.6%) | -1.5 (-1.7, -1.4) | 6.1 (5.4, 6.9) | 90.2% (87.6%, 92.8%) | 0% (0%, 0%) |
| 0% | Inferior | Yes | High | 36.7% (32.1%, 41.5%) | -3.3 (-3.5, -3.2) | 13.7 (12.7, 14.7) | 44.6% (40.2%, 49%) | 0% (0%, 0%) |
| 20% | Similar | No | Low | 34.8% (22.9%, 45.4%) | -5.2 (-5.5, -4.9) | 41.7 (37.8, 45.7) | 69.2% (65.2%, 73.2%) | 0% (0%, 0%) |
| 20% | Similar | No | Medium | 26.3% (17.5%, 34.5%) | -13.7 (-13.9, -13.4) | 193.3 (187.1, 199.4) | 0% (0%, 0%) | 0% (0%, 0%) |
| 20% | Similar | No | High | 20.4% (15%, 25.6%) | -19.6 (-19.7, -19.4) | 386.9 (380, 393.8) | 0% (0%, 0%) | 0% (0%, 0%) |

| | | | | | | | | |
|---|---|---|---|---|---|---|---|---|
| | | Yes | Low | 38.7% (25.6%, 47.6%) | -1.3 (-1.6, -0.9) | 14.6 (12.7, 16.5) | 93% (90.8%, 95.2%) | 0% (0%, 0%) |
| | | | Medium | 35.2% (28.7%, 40.1%) | -4.8 (-5, -4.6) | 27.2 (25.4, 29.1) | 38.4% (34.1%, 42.7%) | 0% (0%, 0%) |
| | | | High | 31.5% (26.2%, 37.3%) | -8.5 (-8.7, -8.4) | 77.1 (74, 80.1) | 1% (0.1%, 1.9%) | 0% (0%, 0%) |
| | Inferior | No | Low | 38.5% (26.8%, 48.2%) | -1.5 (-1.8, -1.2) | 14.2 (12.4, 16) | 93.8% (91.7%, 95.9%) | 0% (0%, 0%) |
| | | | Medium | 35.8% (27.2%, 41.8%) | -4.2 (-4.4, -4) | 22.1 (20.3, 23.9) | 46.8% (42.4%, 51.2%) | 0% (0%, 0%) |
| | | | High | 33.2% (27.2%, 39%) | -6.8 (-7, -6.7) | 50.1 (47.9, 52.4) | 1.6% (0.5%, 2.7%) | 0% (0%, 0%) |
| | | Yes | Low | 39.5% (28.3%, 48.1%) | -0.5 (-0.8, -0.2) | 11.9 (10.5, 13.3) | 95.6% (93.8%, 97.4%) | 0% (0%, 0%) |
| | | | Medium | 38.3% (32.1%, 44%) | -1.7 (-1.9, -1.5) | 7.1 (6.2, 7.9) | 84.8% (81.7%, 87.9%) | 0% (0%, 0%) |
| | | | High | 36.4% (30.7%, 41.2%) | -3.6 (-3.8, -3.5) | 16.3 (15.1, 17.5) | 38% (33.7%, 42.3%) | 0% (0%, 0%) |

**Supplemental Table 22. Mean size of each trial arm, estimate, bias, mean squared error (MSE), coverage, and false negative rate for vaccine efficacy (VE) estimates against any *Shigella* diarrhea from realistically sized trials for all 24 simulation scenarios when using active surveillance for infection and a single outcome regression model.** Hybrid immunity was either not possible (no) or possible such that the first post-vaccination infection conferred additional immunity (yes). Bias was calculated as the average VE estimate for a set of simulations minus the true VE (60%). MSE was calculated as the average of the square of the estimate minus the true parameter. Coverage represents the percent of 95% confidence intervals (CIs) that contained the true VE (60%), and the false negative rate indicates the percent of 95% CIs that incorrectly indicated a non-significant effect.

| VE against infection | Strength of infection-acquired immunity | Hybrid immunity | Force of infection | Mean N (range) | Mean VE estimate (range) | Bias (95% CI) | MSE (95% CI) | Coverage (95% CI) | False negatives (95% CI) |
|---|---|---|---|---|---|---|---|---|---|
| 0% | Similar | No | Low | 2,251 (1,553, 3,745) | 39% (20.2%, 53.3%) | -1 (-1.5, -0.5) | 34.9 (30.5, 39.3) | 96.6% (95%, 98.2%) | 0% (0%, 0%) |
| | | | Medium | 964 (721, 1,349) | 39.3% (17.1%, 56.9%) | -0.7 (-1.3, -0.2) | 38.6 (33.5, 43.6) | 94.8% (92.9%, 96.7%) | 0% (0%, 0%) |
| | | | High | 749 (551, 992) | 39.8% (18.1%, 56%) | -0.2 (-0.8, 0.4) | 43.2 (38, 48.5) | 95.2% (93.3%, 97.1%) | 0.4% (-0.2%, 1%) |
| | | Yes | Low | 2,230 (1,420, 3,924) | 39.2% (22%, 54.9%) | -0.8 (-1.3, -0.3) | 33.9 (29.7, 38) | 95.6% (93.8%, 97.4%) | 0% (0%, 0%) |
| | | | Medium | 976 (710, 1,350) | 39.8% (20.6%, 55.8%) | -0.2 (-0.7, 0.4) | 42.4 (37, 47.8) | 92.8% (90.5%, 95.1%) | 0% (0%, 0%) |
| | | | High | 752 (558, 1,028) | 40% (19.4%, 56%) | 0 (-0.6, 0.5) | 38.6 (33.8, 43.3) | 95% (93.1%, 96.9%) | 0% (0%, 0%) |
| | Inferior | No | Low | 2238 (1,444, 3,583) | 39.4% (21.6%, 59.1%) | -0.6 (-1.1, -0.1) | 36.1 (31.5, 40.7) | 93.4% (91.2%, 95.6%) | 0% (0%, 0%) |
| | | | Medium | 968 (721, 1,307) | 39.5% (20.6%, 59.1%) | -0.5 (-1.1, 0) | 41.6 (36.2, 47) | 93% (90.8%, 95.2%) | 0% (0%, 0%) |
| | | | High | 748 (596, 991) | 39.5% (18.9%, 58.4%) | -0.5 (-1, 0.1) | 37.6 (32.7, 42.6) | 95.4% (93.6%, 97.2%) | 0% (0%, 0%) |
| | | Yes | Low | 2,237 (1,498, 3,583) | 39.5% (19.7%, 57.4%) | -0.5 (-1, 0) | 36.5 (31.8, 41.3) | 94.2% (92.2%, 96.2%) | 0% (0%, 0%) |
| | | | Medium | 968 (747, 1,372) | 39.6% (18.3%, 58.9%) | -0.4 (-1, 0.1) | 38.1 (33, 43.3) | 94% (91.9%, 96.1%) | 0% (0%, 0%) |
| | | | High | 749 (559, 1,003) | 39.6% (20.8%, 56.7%) | -0.4 (-0.9, 0.2) | 38.7 (33.8, 43.6) | 95.6% (93.8%, 97.4%) | 0% (0%, 0%) |
| 20% | Similar | No | Low | 2,235 (1,350, 3,745) | 39.4% (20.5%, 60.4%) | -0.6 (-1.2, -0.1) | 35.5 (31, 40) | 94.4% (92.4%, 96.4%) | 0% (0%, 0%) |
| | | | Medium | 972 (690, 1,327) | 39.6% (20.6%, 53.6%) | -0.4 (-0.9, 0.1) | 32.7 (28.7, 36.6) | 95.4% (93.6%, 97.2%) | 0% (0%, 0%) |
| | | | High | 750 (561, 1,029) | 40% (22.7%, 57.1%) | 0 (-0.5, 0.6) | 37.6 (33.3, 41.9) | 95.2% (93.3%, 97.1%) | 0% (0%, 0%) |

| | | | | | | | | | |
|---|---|---|---|---|---|---|---|---|---|
| | | Yes | Low | 2,229 (1,229, 3,433) | 39.8% (19.8%, 54.8%) | -0.2 (-0.7, 0.3) | 34.1 (29.7, 38.4) | 94.4% (92.4%, 96.4%) | 0% (0%, 0%) |
| | | | Medium | 973 (735, 1,395) | 39.6% (18.3%, 53.8%) | -0.4 (-0.9, 0.1) | 34 (29, 39.1) | 94.6% (92.6%, 96.6%) | 0% (0%, 0%) |
| | | | High | 750 (586, 1,084) | 39.6% (22.5%, 55.1%) | -0.4 (-0.9, 0.2) | 35.8 (31.1, 40.4) | 93.8% (91.7%, 95.9%) | 0% (0%, 0%) |
| | Inferior | No | Low | 2,249 (1,471, 4,847) | 39.5% (22.1%, 53.5%) | -0.5 (-1, 0) | 31.9 (27.9, 36) | 95.2% (93.3%, 97.1%) | 0% (0%, 0%) |
| | | | Medium | 971 (721, 1,374) | 39.7% (20.6%, 52.4%) | -0.3 (-0.8, 0.3) | 33.8 (29.5, 38) | 96.2% (94.5%, 97.9%) | 0% (0%, 0%) |
| | | | High | 744 (544, 1,028) | 39.8% (14.6%, 55.3%) | -0.2 (-0.8, 0.3) | 38.4 (33.1, 43.6) | 94.8% (92.9%, 96.7%) | 0.2% (-0.2%, 0.6%) |
| | | Yes | Low | 2,244 (1,498, 4,847) | 39.4% (21%, 55.2%) | -0.6 (-1.1, -0.1) | 31.8 (27.6, 35.9) | 95.2% (93.3%, 97.1%) | 0% (0%, 0%) |
| | | | Medium | 971 (715, 1,349) | 40% (21%, 53.8%) | 0 (-0.5, 0.5) | 33 (29, 37) | 95.6% (93.8%, 97.4%) | 0% (0%, 0%) |
| | | | High | 743 (523, 1,111) | 39.7% (13.9%, 53.3%) | -0.3 (-0.8, 0.2) | 38 (32.9, 43.1) | 95.2% (93.3%, 97.1%) | 0.2% (-0.2%, 0.6%) |

**Supplemental Table 23. Mean estimate, bias, mean squared error (MSE), coverage, and false negative rate for vaccine efficacy (VE) estimates against any *Shigella* diarrhea from realistically sized trials for all 24 simulation scenarios when using symptom-based reporting and a single outcome regression model.** Hybrid immunity was either not possible (no) or possible such that the first post-vaccination infection conferred additional immunity (yes). Bias was calculated as the average VE estimate for a set of simulations minus the true VE (60%). MSE was calculated as the average of the square of the estimate minus the true parameter. Coverage represents the percent of 95% confidence intervals (CIs) that contained the true VE (60%), and the false negative rate indicates the percent of 95% CIs that incorrectly indicated a non-significant effect.

| VE against infection | Strength of infection-acquired immunity | Hybrid immunity | Force of infection | Mean N (range) | Mean VE estimate (range) | Bias (95% CI) | MSE (95% CI) | Coverage (95% CI) | False negatives (95% CI) |
|---|---|---|---|---|---|---|---|---|---|
| 0% | Similar | No | Low | 2,181 (1,497, 3,745) | 35.8% (19.1%, 51.3%) | -4.2 (-4.7, -3.7) | 52.3 (46, 58.6) | 89.8% (87.1%, 92.5%) | 0% (0%, 0%) |
| | | | Medium | 856 (662, 1,128) | 30.1% (9.9%, 49.4%) | -9.9 (-10.5, -9.4) | 138.2 (125.5, 150.9) | 59.6% (55.3%, 63.9%) | 1.8% (0.6%, 3%) |
| | | | High | 618 (474, 814) | 26.4% (5.3%, 45.2%) | -13.6 (-14.2, -13.1) | 227.5 (210.2, 244.8) | 36.4% (32.2%, 40.6%) | 5.6% (3.6%, 7.6%) |
| | | Yes | Low | 2,164 (1,420, 3,745) | 39.7% (23.7%, 54.8%) | -0.3 (-0.8, 0.2) | 32.4 (28.4, 36.3) | 95.2% (93.3%, 97.1%) | 0% (0%, 0%) |
| | | | Medium | 883 (653, 1,193) | 39.4% (21.8%, 55.8%) | -0.6 (-1.1, -0.1) | 33.5 (29.2, 37.7) | 94.6% (92.6%, 96.6%) | 0% (0%, 0%) |
| | | | High | 652 (469, 831) | 37.2% (17%, 52%) | -2.8 (-3.3, -2.3) | 39.8 (34.6, 45.1) | 92.4% (90.1%, 94.7%) | 0% (0%, 0%) |
| | Inferior | No | Low | 2,090 (1,419, 3,433) | 38.8% (20.2%, 57.3%) | -1.2 (-1.7, -0.8) | 32.6 (28.6, 36.7) | 94.4% (92.4%, 96.4%) | 0% (0%, 0%) |
| | | | Medium | 795 (627, 1,083) | 36.9% (19.6%, 51.5%) | -3.1 (-3.6, -2.6) | 42.1 (36.6, 47.5) | 92% (89.6%, 94.4%) | 0% (0%, 0%) |
| | | | High | 561 (441, 702) | 35% (10.6%, 49%) | -5 (-5.5, -4.5) | 60.3 (52.6, 67.9) | 85.2% (82.1%, 88.3%) | 0.2% (-0.2%, 0.6%) |
| | | Yes | Low | 2,095 (1,444, 3,433) | 39.8% (22.3%, 57.3%) | -0.2 (-0.7, 0.2) | 30.1 (26.2, 34) | 94.8% (92.9%, 96.7%) | 0% (0%, 0%) |
| | | | Medium | 796 (637, 1,159) | 39.3% (22.8%, 53.8%) | -0.7 (-1.2, -0.2) | 31.9 (27.9, 35.9) | 94.6% (92.6%, 96.6%) | 0% (0%, 0%) |
| | | | High | 565 (443, 734) | 38.2% (20.9%, 53.1%) | -1.8 (-2.3, -1.3) | 33.2 (29.2, 37.3) | 94.8% (92.9%, 96.7%) | 0% (0%, 0%) |
| 20% | Similar | No | Low | 2,164 (1,328, 3,745) | 36.1% (16.7%, 55.7%) | -3.9 (-4.4, -3.4) | 52.8 (46.4, 59.2) | 89.4% (86.7%, 92.1%) | 0.2% (-0.2%, 0.6%) |
| | | | Medium | 861 (642, 1,176) | 30.3% (11.1%, 47.8%) | -9.7 (-10.2, -9.2) | 128.8 (117.2, 140.4) | 61.4% (57.1%, 65.7%) | 1.4% (0.4%, 2.4%) |
| | | | High | 618 (500, 783) | 26.4% (5.2%, 42.7%) | -13.6 (-14.2, -13.1) | 224.3 (207.9, 240.7) | 34% (29.8%, 38.2%) | 5.2% (3.3%, 7.1%) |

| | | | | | | | | | |
|---|---|---|---|---|---|---|---|---|---|
| | | Yes | Low | 2,158<br>(1,193, 3,296) | 39.2%<br>(20.3%, 53.1%) | -0.8<br>(-1.3, -0.3) | 33.8<br>(29.5, 38.1) | 94%<br>(91.9%, 96.1%) | 0%<br>(0%, 0%) |
| | | | Medium | 878<br>(673, 1,248) | 37.5%<br>(18.5%, 51.4%) | -2.5<br>(-3, -2) | 36.3<br>(31.2, 41.5) | 92.4%<br>(90.1%, 94.7%) | 0%<br>(0%, 0%) |
| | | | High | 641<br>(496, 839) | 35.1%<br>(16.6%, 49.5%) | -4.9<br>(-5.4, -4.4) | 57.7<br>(50.4, 64.9) | 86%<br>(83%, 89%) | 0%<br>(0%, 0%) |
| | Inferior | No | Low | 2,097<br>(1,349, 3,745) | 38.7%<br>(19.2%, 51.5%) | -1.3<br>(-1.8, -0.9) | 32.7<br>(28.4, 37) | 95.4%<br>(93.6%, 97.2%) | 0%<br>(0%, 0%) |
| | | | Medium | 797<br>(604, 1,083) | 37%<br>(19.9%, 50.2%) | -3<br>(-3.5, -2.5) | 43<br>(37.5, 48.5) | 90.2%<br>(87.6%, 92.8%) | 0%<br>(0%, 0%) |
| | | | High | 556<br>(445, 708) | 35.3%<br>(16.4%, 51%) | -4.7<br>(-5.2, -4.2) | 52.8<br>(46.7, 58.9) | 88.4%<br>(85.6%, 91.2%) | 0%<br>(0%, 0%) |
| | | Yes | Low | 2,096<br>(1,372, 3,924) | 39.4%<br>(21.5%, 53.5%) | -0.6<br>(-1.1, -0.1) | 30.7<br>(26.9, 34.5) | 96.2%<br>(94.5%, 97.9%) | 0%<br>(0%, 0%) |
| | | | Medium | 799<br>(609, 1,112) | 39%<br>(22%, 58.4%) | -1<br>(-1.5, -0.5) | 31.9<br>(27.9, 36) | 94.6%<br>(92.6%, 96.6%) | 0%<br>(0%, 0%) |
| | | | High | 560<br>(429, 768) | 37.9%<br>(18.4%, 51.2%) | -2.1<br>(-2.6, -1.6) | 35.6<br>(31, 40.1) | 94.2%<br>(92.2%, 96.2%) | 0%<br>(0%, 0%) |

**Supplemental Table 24. Mean estimate, bias, mean squared error (MSE), coverage, and false negative rate for vaccine efficacy (VE) estimates against any *Shigella* diarrhea from realistically sized trials for all 24 simulation scenarios when using active surveillance for infection and a stratified recurrent outcome regression model.** Hybrid immunity was either not possible (no) or possible such that the first post-vaccination infection conferred additional immunity (yes). Bias was calculated as the average VE estimate for a set of simulations minus the true VE (60%). MSE was calculated as the average of the square of the estimate minus the true parameter. Coverage represents the percent of 95% confidence intervals (CIs) that contained the true VE (60%), and the false negative rate indicates the percent of 95% CIs that incorrectly indicated a non-significant effect.

| VE against infection | Strength of infection-acquired immunity | Hybrid immunity | Force of infection | Mean N (range) | Mean VE estimate (range) | Bias (95% CI) | MSE (95% CI) | Coverage (95% CI) | False negatives (95% CI) |
|---|---|---|---|---|---|---|---|---|---|
| 0% | Similar | No | Low | 2,103 (1,419, 3,580) | 34.1% (16.6%, 50.8%) | -5.9 (-6.4, -5.4) | 70.5 (62.7, 78.3) | 80.8% (77.3%, 84.3%) | 0% (0%, 0%) |
| | | | Medium | 767 (600, 1,043) | 25.7% (6.3%, 43%) | -14.3 (-14.9, -13.7) | 245 (226.9, 263.2) | 31.2% (27.1%, 35.3%) | 7% (4.8%, 9.2%) |
| | | | High | 518 (403, 653) | 20.3% (-3.2%, 37.1%) | -19.7 (-20.3, -19.2) | 435.2 (409.7, 460.8) | 8% (5.6%, 10.4%) | 19.6% (16.1%, 23.1%) |
| | | Yes | Low | 2,098 (1,395, 3,582) | 38.9% (23.9%, 55.2%) | -1.1 (-1.6, -0.6) | 33.5 (29.4, 37.5) | 95% (93.1%, 96.9%) | 0% (0%, 0%) |
| | | | Medium | 808 (618, 1,055) | 36.7% (18.9%, 52.8%) | -3.3 (-3.8, -2.8) | 46.4 (40.7, 52.2) | 88.8% (86%, 91.6%) | 0% (0%, 0%) |
| | | | High | 573 (419, 716) | 33% (10.3%, 47.9%) | -7 (-7.6, -6.5) | 84.9 (75.3, 94.6) | 78.2% (74.6%, 81.8%) | 0.4% (-0.2%, 1%) |
| | Inferior | No | Low | 1,984 (1,328, 3,295) | 38.3% (20%, 57.8%) | -1.7 (-2.2, -1.2) | 33.7 (29.5, 38) | 95.6% (93.8%, 97.4%) | 0% (0%, 0%) |
| | | | Medium | 682 (545, 895) | 35.2% (13.4%, 52%) | -4.8 (-5.4, -4.3) | 59.2 (51.4, 66.9) | 85.2% (82.1%, 88.3%) | 0.6% (-0.1%, 1.3%) |
| | | | High | 444 (368, 548) | 32.1% (6.7%, 46.7%) | -7.9 (-8.4, -7.4) | 94.1 (84.5, 103.7) | 70% (66%, 74%) | 0.4% (-0.2%, 1%) |
| | | Yes | Low | 1,991 (1,351, 3,295) | 39.5% (21%, 58.9%) | -0.5 (-1, 0) | 30.7 (26.7, 34.7) | 95.4% (93.6%, 97.2%) | 0% (0%, 0%) |
| | | | Medium | 686 (555, 980) | 38.2% (21.5%, 52.9%) | -1.8 (-2.3, -1.3) | 34.4 (30.1, 38.6) | 93.2% (91%, 95.4%) | 0% (0%, 0%) |
| | | | High | 451 (366, 552) | 36.1% (16.1%, 54%) | -3.9 (-4.3, -3.4) | 44.7 (39.2, 50.1) | 89.8% (87.1%, 92.5%) | 0% (0%, 0%) |
| 20% | Similar | No | Low | 2,087 (1,287, 3,582) | 35% (13%, 54.9%) | -5 (-5.6, -4.5) | 63.5 (55.9, 71) | 87.8% (84.9%, 90.7%) | 0.6% (-0.1%, 1.3%) |
| | | | Medium | 772 (583, 1,083) | 27.4% (11.8%, 44.3%) | -12.6 (-13.2, -12.1) | 194.5 (180.7, 208.4) | 41% (36.7%, 45.3%) | 2.6% (1.2%, 4%) |
| | | | High | 516 (407, 642) | 21.4% (-5.4%, 38.5%) | -18.6 (-19.2, -18) | 389.4 (366.2, 412.7) | 13.6% (10.6%, 16.6%) | 16% (12.8%, 19.2%) |

| | | | | | | | | | |
|---|---|---|---|---|---|---|---|---|---|
| | | Yes | Low | 2,093<br>(1,210, 3,169) | 39.5%<br>(20%, 55%) | -0.5<br>(-1, 0) | 33.2<br>(29.1, 37.3) | 94.8%<br>(92.9%, 96.7%) | 0%<br>(0%, 0%) |
| | | | Medium | 804<br>(632, 1,113) | 37.4%<br>(18.9%, 51.8%) | -2.6<br>(-3, -2.1) | 37.9<br>(33.1, 42.7) | 92.2%<br>(89.8%, 94.6%) | 0%<br>(0%, 0%) |
| | | | High | 561<br>(442, 710) | 34.6%<br>(15.4%, 48.8%) | -5.4<br>(-5.9, -4.9) | 61.2<br>(53.5, 68.8) | 84%<br>(80.8%, 87.2%) | 0.2%<br>(-0.2%, 0.6%) |
| | Inferior | No | Low | 1,996<br>(1,307, 3,296) | 38.5%<br>(21.5%, 54.1%) | -1.5<br>(-2, -1) | 33.6<br>(29.3, 38) | 95.8%<br>(94%, 97.6%) | 0%<br>(0%, 0%) |
| | | | Medium | 683<br>(527, 895) | 35.9%<br>(15.6%, 51.1%) | -4.1<br>(-4.7, -3.6) | 54.4<br>(47.4, 61.4) | 85.6%<br>(82.5%, 88.7%) | 0.2%<br>(-0.2%, 0.6%) |
| | | | High | 438<br>(350, 563) | 33.3%<br>(13%, 46.5%) | -6.7<br>(-7.1, -6.2) | 75.1<br>(67, 83.3) | 77.6%<br>(73.9%, 81.3%) | 0.2%<br>(-0.2%, 0.6%) |
| | | Yes | Low | 1,994<br>(1,327, 3,433) | 39.5%<br>(20.2%, 52.7%) | -0.5<br>(-1, 0) | 30.1<br>(26.3, 33.9) | 95.8%<br>(94%, 97.6%) | 0%<br>(0%, 0%) |
| | | | Medium | 688<br>(527, 946) | 38.9%<br>(19.5%, 55.3%) | -1.1<br>(-1.6, -0.6) | 32.3<br>(28, 36.7) | 94.2%<br>(92.2%, 96.2%) | 0%<br>(0%, 0%) |
| | | | High | 448<br>(345, 587) | 36.9%<br>(19.4%, 50.2%) | -3.1<br>(-3.6, -2.6) | 39.4<br>(34.3, 44.4) | 90%<br>(87.4%, 92.6%) | 0%<br>(0%, 0%) |

**Supplemental Table 25. Mean estimate, bias, mean squared error (MSE), coverage, and false negative rate for vaccine efficacy (VE) estimates against any *Shigella* diarrhea from realistically sized trials for all 24 simulation scenarios when using symptom-based reporting and a stratified recurrent outcome regression model.** Hybrid immunity was either not possible (no) or possible such that the first post-vaccination infection conferred additional immunity (yes). Bias was calculated as the average VE estimate for a set of simulations minus the true VE (60%). MSE was calculated as the average of the square of the estimate minus the true parameter. Coverage represents the percent of 95% confidence intervals (CIs) that contained the true VE (60%), and the false negative rate indicates the percent of 95% CIs that incorrectly indicated a non-significant effect.

| VE against infection | Strength of infection-acquired immunity | Hybrid immunity | Force of infection | Mean N (range) | Mean VE estimate (range) | Bias (95% CI) | MSE (95% CI) | Coverage (95% CI) | False negatives (95% CI) |
|---|---|---|---|---|---|---|---|---|---|
| 0% | Similar | No | Low | 2,151 (1,419, 3,744) | 34.5% (18%, 49.3%) | -5.5 (-6, -5) | 64.8 (57.5, 72.1) | 84% (80.8%, 87.2%) | 0.2% (-0.2%, 0.6%) |
| | | | Medium | 780 (613, 1,056) | 26.6% (7.6%, 44.9%) | -13.4 (-13.9, -12.8) | 219.1 (202.1, 236.2) | 39.2% (34.9%, 43.5%) | 5.4% (3.4%, 7.4%) |
| | | | High | 528 (408, 674) | 21% (-1.8%, 37.7%) | -19 (-19.6, -18.4) | 406.3 (382, 430.5) | 12% (9.2%, 14.8%) | 16.8% (13.5%, 20.1%) |
| | | Yes | Low | 2,141 (1,395, 3,582) | 39.6% (24.6%, 54.9%) | -0.4 (-0.9, 0.1) | 31.6 (27.8, 35.4) | 94.8% (92.9%, 96.7%) | 0% (0%, 0%) |
| | | | Medium | 823 (622, 1,097) | 38.3% (22.1%, 55.4%) | -1.7 (-2.2, -1.2) | 35.6 (31.2, 39.9) | 92.6% (90.3%, 94.9%) | 0% (0%, 0%) |
| | | | High | 587 (421, 741) | 34.9% (11.9%, 49.1%) | -5.1 (-5.7, -4.6) | 60.3 (52.3, 68.3) | 86.4% (83.4%, 89.4%) | 0.4% (-0.2%, 1%) |
| | Inferior | No | Low | 2,021 (1,371, 3,295) | 38.4% (20.6%, 57.6%) | -1.6 (-2.1, -1.1) | 33 (28.9, 37.1) | 94.6% (92.6%, 96.6%) | 0% (0%, 0%) |
| | | | Medium | 692 (559, 935) | 35.7% (14.4%, 52.9%) | -4.3 (-4.8, -3.8) | 53 (46.2, 59.7) | 88% (85.2%, 90.8%) | 0.2% (-0.2%, 0.6%) |
| | | | High | 452 (370, 551) | 32.9% (6.9%, 46.5%) | -7.1 (-7.6, -6.7) | 82.4 (73.7, 91.1) | 75.4% (71.6%, 79.2%) | 0.4% (-0.2%, 1%) |
| | | Yes | Low | 2,028 (1,419, 3,295) | 39.7% (23.2%, 58.7%) | -0.3 (-0.8, 0.2) | 30.1 (26.2, 33.9) | 95.4% (93.6%, 97.2%) | 0% (0%, 0%) |
| | | | Medium | 697 (559, 1,003) | 38.7% (22.9%, 53.3%) | -1.3 (-1.8, -0.8) | 32.5 (28.7, 36.3) | 94.2% (92.2%, 96.2%) | 0% (0%, 0%) |
| | | | High | 459 (371, 563) | 36.9% (15.9%, 55%) | -3.1 (-3.6, -2.6) | 38.8 (33.9, 43.8) | 92.4% (90.1%, 94.7%) | 0% (0%, 0%) |
| 20% | Similar | No | Low | 2,132 (1,349, 3,582) | 34.8% (13.3%, 54.2%) | -5.2 (-5.8, -4.7) | 65.3 (57.9, 72.7) | 85.6% (82.5%, 88.7%) | 0.4% (-0.2%, 1%) |
| | | | Medium | 784 (591, 1,069) | 26.9% (10.5%, 44.1%) | -13.1 (-13.6, -12.6) | 206.3 (192.2, 220.3) | 38% (33.7%, 42.3%) | 3.2% (1.7%, 4.7%) |
| | | | High | 526 (411, 668) | 20.8% (-1.3%, 37%) | -19.2 (-19.8, -18.6) | 410.7 (387.4, 434.1) | 11% (8.3%, 13.7%) | 17.2% (13.9%, 20.5%) |

| | | | | | | | | | |
|---|---|---|---|---|---|---|---|---|---|
| | | Yes | Low | 2,138<br>(1,210, 3,295) | 39.1%<br>(19.1%, 53.5%) | -0.9<br>(-1.4, -0.4) | 33.8<br>(29.5, 38) | 93.8%<br>(91.7%, 95.9%) | 0%<br>(0%, 0%) |
| | | | Medium | 819<br>(642, 1,113) | 36.5%<br>(20.7%, 50.5%) | -3.5<br>(-4, -3) | 42.8<br>(37.6, 48) | 90.4%<br>(87.8%, 93%) | 0%<br>(0%, 0%) |
| | | | High | 573<br>(451, 747) | 33.2%<br>(15.4%, 48.3%) | -6.8<br>(-7.3, -6.3) | 78.7<br>(69.9, 87.5) | 78%<br>(74.4%, 81.6%) | 0.2%<br>(-0.2%, 0.6%) |
| | Inferior | No | Low | 2,032<br>(1,327, 3,296) | 38.4%<br>(21.3%, 53.1%) | -1.6<br>(-2.1, -1.1) | 33.8<br>(29.5, 38.1) | 95.2%<br>(93.3%, 97.1%) | 0%<br>(0%, 0%) |
| | | | Medium | 694<br>(530, 924) | 35.8%<br>(15.2%, 51.9%) | -4.2<br>(-4.8, -3.7) | 55.5<br>(48.3, 62.7) | 85%<br>(81.9%, 88.1%) | 0.2%<br>(-0.2%, 0.6%) |
| | | | High | 446<br>(353, 570) | 33.2%<br>(13.2%, 46%) | -6.8<br>(-7.3, -6.3) | 77.3<br>(69.2, 85.5) | 76.6%<br>(72.9%, 80.3%) | 0.2%<br>(-0.2%, 0.6%) |
| | | Yes | Low | 2,033<br>(1,327, 3,581) | 39.4%<br>(20.8%, 52.6%) | -0.6<br>(-1.1, -0.1) | 30.5<br>(26.6, 34.3) | 95.2%<br>(93.3%, 97.1%) | 0%<br>(0%, 0%) |
| | | | Medium | 699<br>(523, 946) | 38.7%<br>(19.7%, 55.7%) | -1.3<br>(-1.8, -0.8) | 32.5<br>(28.1, 37) | 93.2%<br>(91%, 95.4%) | 0%<br>(0%, 0%) |
| | | | High | 455<br>(353, 591) | 36.7%<br>(17.5%, 49.5%) | -3.3<br>(-3.8, -2.8) | 40.7<br>(35.4, 46.1) | 91%<br>(88.5%, 93.5%) | 0%<br>(0%, 0%) |

**Supplemental Table 26. Mean estimate, bias, mean squared error (MSE), coverage, and false negative rate for vaccine efficacy (VE) estimates against any *Shigella* diarrhea from realistically sized trials for all 24 simulation scenarios when using active surveillance for infection and a crude recurrent outcome regression model.** Hybrid immunity was either not possible (no) or possible such that the first post-vaccination infection conferred additional immunity (yes). Bias was calculated as the average VE estimate for a set of simulations minus the true VE (60%). MSE was calculated as the average of the square of the estimate minus the true parameter. Coverage represents the percent of 95% confidence intervals (CIs) that contained the true VE (60%), and the false negative rate indicates the percent of 95% CIs that incorrectly indicated a non-significant effect.

| VE against infection | Strength of infection-acquired immunity | Hybrid immunity | Force of infection | Mean N (range) | Mean VE estimate (range) | Bias (95% CI) | MSE (95% CI) | Coverage (95% CI) | False negatives (95% CI) |
|---|---|---|---|---|---|---|---|---|---|
| 0% | Similar | No | Low | 2,103 (1,419, 3,580) | 34.1% (16.5%, 50.6%) | -5.9 (-6.4, -5.4) | 69.6 (61.9, 77.2) | 80.8% (77.3%, 84.3%) | 0% (0%, 0%) |
| | | | Medium | 767 (600, 1,043) | 25.8% (6.5%, 43.2%) | -14.2 (-14.8, -13.7) | 242.1 (224.5, 259.7) | 29.4% (25.4%, 33.4%) | 6.4% (4.3%, 8.5%) |
| | | | High | 518 (403, 653) | 20.2% (-2.9%, 35.8%) | -19.8 (-20.4, -19.2) | 434.4 (409.4, 459.4) | 6.8% (4.6%, 9%) | 18% (14.6%, 21.4%) |
| | | Yes | Low | 2,098 (1,395, 3,582) | 38.9% (24.2%, 54.8%) | -1.1 (-1.6, -0.6) | 32.2 (28.3, 36.1) | 95.2% (93.3%, 97.1%) | 0% (0%, 0%) |
| | | | Medium | 808 (618, 1,055) | 36.8% (20.8%, 51.9%) | -3.2 (-3.7, -2.7) | 42.5 (37.3, 47.7) | 88.8% (86%, 91.6%) | 0% (0%, 0%) |
| | | | High | 573 (419, 716) | 33% (11.6%, 47.5%) | -7 (-7.5, -6.5) | 81.4 (72.3, 90.5) | 76% (72.3%, 79.7%) | 0.4% (-0.2%, 1%) |
| | Inferior | No | Low | 1,984 (1,328, 3,295) | 38.3% (20.1%, 57.7%) | -1.7 (-2.2, -1.2) | 33.7 (29.5, 38) | 95.2% (93.3%, 97.1%) | 0% (0%, 0%) |
| | | | Medium | 682 (545, 895) | 35.2% (13.4%, 52.1%) | -4.8 (-5.3, -4.3) | 58.1 (50.5, 65.6) | 85.2% (82.1%, 88.3%) | 0.4% (-0.2%, 1%) |
| | | | High | 444 (368, 548) | 32.1% (6.8%, 46.7%) | -7.9 (-8.4, -7.4) | 93.3 (83.9, 102.8) | 69% (64.9%, 73.1%) | 0.4% (-0.2%, 1%) |
| | | Yes | Low | 1,991 (1,351, 3,295) | 39.5% (21.2%, 58.8%) | -0.5 (-1, 0) | 30.6 (26.6, 34.5) | 95.4% (93.6%, 97.2%) | 0% (0%, 0%) |
| | | | Medium | 686 (555, 980) | 38.2% (22.5%, 52.8%) | -1.8 (-2.3, -1.3) | 33.3 (29.2, 37.3) | 93.4% (91.2%, 95.6%) | 0% (0%, 0%) |
| | | | High | 451 (366, 552) | 36.1% (16.9%, 54.1%) | -3.9 (-4.3, -3.4) | 44 (38.7, 49.3) | 89.2% (86.5%, 91.9%) | 0% (0%, 0%) |
| 20% | Similar | No | Low | 2,087 (1,287, 3,582) | 34.4% (12.4%, 54.3%) | -5.6 (-6.2, -5.1) | 69.4 (61.4, 77.3) | 84.2% (81%, 87.4%) | 0.6% (-0.1%, 1.3%) |
| | | | Medium | 772 (583, 1,083) | 26.1% (10.7%, 43.4%) | -13.9 (-14.4, -13.4) | 227.2 (212.4, 242) | 30.8% (26.8%, 34.8%) | 4% (2.3%, 5.7%) |
| | | | High | 516 (407, 642) | 20% (-5.2%, 36.9%) | -20 (-20.6, -19.4) | 442.8 (418.6, 467) | 7.8% (5.4%, 10.2%) | 18.8% (15.4%, 22.2%) |

| | | | | | | | | | |
|---|---|---|---|---|---|---|---|---|---|
| | | Yes | Low | 2,093 (1,210, 3,169) | 38.5% (19.6%, 54.5%) | -1.5 (-2, -1) | 35.1 (30.7, 39.5) | 93.6% (91.5%, 95.7%) | 0% (0%, 0%) |
| | | | Medium | 804 (632, 1,113) | 34.9% (18.2%, 49.4%) | -5.1 (-5.5, -4.6) | 55.5 (49.2, 61.9) | 85.2% (82.1%, 88.3%) | 0% (0%, 0%) |
| | | | High | 561 (442, 710) | 31.3% (13.7%, 45.9%) | -8.7 (-9.2, -8.2) | 106.8 (96.6, 117) | 65.2% (61%, 69.4%) | 0.2% (-0.2%, 0.6%) |
| | Inferior | No | Low | 1,996 (1,307, 3,296) | 38.3% (20.9%, 53.9%) | -1.7 (-2.2, -1.2) | 33.9 (29.5, 38.3) | 95.8% (94%, 97.6%) | 0% (0%, 0%) |
| | | | Medium | 683 (527, 895) | 35.4% (15.7%, 51.1%) | -4.6 (-5.1, -4.1) | 57.5 (50.2, 64.8) | 84.4% (81.2%, 87.6%) | 0.4% (-0.2%, 1%) |
| | | | High | 438 (350, 563) | 32.7% (13%, 45.9%) | -7.3 (-7.8, -6.8) | 82.9 (74.4, 91.4) | 74% (70.2%, 77.8%) | 0.2% (-0.2%, 0.6%) |
| | | Yes | Low | 1,994 (1,327, 3,433) | 39.2% (20.3%, 52%) | -0.8 (-1.2, -0.3) | 30.1 (26.3, 33.9) | 96.4% (94.8%, 98%) | 0% (0%, 0%) |
| | | | Medium | 688 (527, 946) | 38.3% (19.1%, 54.6%) | -1.7 (-2.2, -1.3) | 33.5 (28.8, 38.1) | 91.6% (89.2%, 94%) | 0% (0%, 0%) |
| | | | High | 448 (345, 587) | 36.1% (19.3%, 49.7%) | -3.9 (-4.4, -3.5) | 44.2 (38.7, 49.7) | 89% (86.3%, 91.7%) | 0% (0%, 0%) |

**Supplemental Table 27. Mean estimate, bias, mean squared error (MSE), coverage, and false negative rate for vaccine efficacy (VE) estimates against any *Shigella* diarrhea from realistically sized trials for all 24 simulation scenarios when using symptom-based reporting and a crude recurrent outcome regression model.** Hybrid immunity was either not possible (no) or possible such that the first post-vaccination infection conferred additional immunity (yes). Bias was calculated as the average VE estimate for a set of simulations minus the true VE (60%). MSE was calculated as the average of the square of the estimate minus the true parameter. Coverage represents the percent of 95% confidence intervals (CIs) that contained the true VE (60%), and the false negative rate indicates the percent of 95% CIs that incorrectly indicated a non-significant effect.

| VE against infection | Strength of infection-acquired immunity | Hybrid immunity | Force of infection | Mean N (range) | Mean VE estimate (range) | Bias (95% CI) | MSE (95% CI) | Coverage (95% CI) | False negatives (95% CI) |
|---|---|---|---|---|---|---|---|---|---|
| 0% | Similar | No | Low | 2,151 (1,419, 3,744) | 34.1% (18.1%, 48.9%) | -5.9 (-6.4, -5.4) | 69 (61.5, 76.6) | 80.2% (76.7%, 83.7%) | 0.2% (-0.2%, 0.6%) |
| 0% | Similar | No | Medium | 780 (613, 1,056) | 25.8% (7.5%, 43.6%) | -14.2 (-14.7, -13.6) | 238.9 (221.6, 256.3) | 30.6% (26.6%, 34.6%) | 5.6% (3.6%, 7.6%) |
| 0% | Similar | No | High | 528 (408, 674) | 20.1% (-2.5%, 35.5%) | -19.9 (-20.4, -19.3) | 436.6 (412.2, 461.1) | 6.6% (4.4%, 8.8%) | 17.6% (14.3%, 20.9%) |
| 0% | Similar | Yes | Low | 2,141 (1,395, 3,582) | 38.9% (24.2%, 54.1%) | -1.1 (-1.6, -0.6) | 31.8 (28, 35.7) | 95% (93.1%, 96.9%) | 0% (0%, 0%) |
| 0% | Similar | Yes | Medium | 823 (622, 1,097) | 36.8% (20.9%, 52.5%) | -3.2 (-3.7, -2.7) | 41.4 (36.4, 46.4) | 88.6% (85.8%, 91.4%) | 0% (0%, 0%) |
| 0% | Similar | Yes | High | 587 (421, 741) | 33% (10.4%, 47.7%) | -7 (-7.5, -6.5) | 80.3 (71.1, 89.4) | 75.8% (72%, 79.6%) | 0.4% (-0.2%, 1%) |
| 0% | Inferior | No | Low | 2,021 (1,371, 3,295) | 38.3% (20.3%, 57.5%) | -1.7 (-2.2, -1.2) | 33.3 (29.1, 37.4) | 94.6% (92.6%, 96.6%) | 0% (0%, 0%) |
| 0% | Inferior | No | Medium | 692 (559, 935) | 35.3% (14.5%, 52.3%) | -4.7 (-5.2, -4.2) | 55.7 (48.7, 62.7) | 84.6% (81.4%, 87.8%) | 0.2% (-0.2%, 0.6%) |
| 0% | Inferior | No | High | 452 (370, 551) | 32.2% (7.1%, 46.2%) | -7.8 (-8.3, -7.3) | 90.8 (81.8, 99.9) | 69.4% (65.4%, 73.4%) | 0.4% (-0.2%, 1%) |
| 0% | Inferior | Yes | Low | 2,028 (1,419, 3,295) | 39.5% (22.8%, 58.5%) | -0.5 (-1, 0) | 29.9 (26, 33.7) | 95.6% (93.8%, 97.4%) | 0% (0%, 0%) |
| 0% | Inferior | Yes | Medium | 697 (559, 1,003) | 38.2% (22.8%, 52.6%) | -1.8 (-2.3, -1.3) | 33.2 (29.2, 37.2) | 93.4% (91.2%, 95.6%) | 0% (0%, 0%) |
| 0% | Inferior | Yes | High | 459 (371, 563) | 36.2% (15.6%, 54.1%) | -3.8 (-4.3, -3.3) | 42.8 (37.5, 48.1) | 89.4% (86.7%, 92.1%) | 0% (0%, 0%) |
| 20% | Similar | No | Low | 2,132 (1,349, 3,582) | 34.3% (13.2%, 53.6%) | -5.7 (-6.2, -5.1) | 68.9 (61.3, 76.5) | 83.6% (80.4%, 86.8%) | 0.4% (-0.2%, 1%) |
| 20% | Similar | No | Medium | 784 (591, 1,069) | 26.1% (10.6%, 43.1%) | -13.9 (-14.4, -13.4) | 225.9 (211.5, 240.3) | 28.6% (24.6%, 32.6%) | 3.2% (1.7%, 4.7%) |
| 20% | Similar | No | High | 526 (411, 668) | 20% (-2.1%, 36.4%) | -20 (-20.6, -19.5) | 441.5 (418, 465) | 6.6% (4.4%, 8.8%) | 17.8% (14.4%, 21.2%) |

| | | | | | | | | | |
|---|---|---|---|---|---|---|---|---|---|
| | | Yes | Low | 2,138 (1,210, 3,295) | 38.5% (18.8%, 53.2%) | -1.5 (-2, -1) | 34.7 (30.2, 39.2) | 93.6% (91.5%, 95.7%) | 0% (0%, 0%) |
| | | | Medium | 819 (642, 1,113) | 34.9% (19.8%, 49.1%) | -5.1 (-5.5, -4.6) | 54 (47.9, 60.1) | 85.8% (82.7%, 88.9%) | 0% (0%, 0%) |
| | | | High | 573 (451, 747) | 31.3% (14.9%, 45.8%) | -8.7 (-9.2, -8.2) | 106.4 (96.3, 116.4) | 64.2% (60%, 68.4%) | 0.2% (-0.2%, 0.6%) |
| | Inferior | No | Low | 2,032 (1,327, 3,296) | 38.3% (20.9%, 53.7%) | -1.7 (-2.2, -1.2) | 34.1 (29.7, 38.4) | 94.6% (92.6%, 96.6%) | 0% (0%, 0%) |
| | | | Medium | 694 (530, 924) | 35.4% (15.7%, 51.8%) | -4.6 (-5.1, -4.1) | 57.1 (49.9, 64.3) | 85% (81.9%, 88.1%) | 0.2% (-0.2%, 0.6%) |
| | | | High | 446 (353, 570) | 32.7% (13.1%, 45.6%) | -7.3 (-7.8, -6.8) | 82.7 (74.4, 91) | 74% (70.2%, 77.8%) | 0.2% (-0.2%, 0.6%) |
| | | Yes | Low | 2,033 (1,327, 3,581) | 39.2% (20.7%, 52.6%) | -0.8 (-1.3, -0.3) | 30.6 (26.7, 34.5) | 95.6% (93.8%, 97.4%) | 0% (0%, 0%) |
| | | | Medium | 699 (523, 946) | 38.3% (19.5%, 55.3%) | -1.7 (-2.2, -1.2) | 33.1 (28.6, 37.7) | 92% (89.6%, 94.4%) | 0% (0%, 0%) |
| | | | High | 455 (353, 591) | 36% (18.7%, 48.7%) | -4 (-4.4, -3.5) | 44.1 (38.6, 49.7) | 88.6% (85.8%, 91.4%) | 0% (0%, 0%) |
